# Risk Factors for Pediatric Critical COVID-19: A Systematic Review and Meta-Analysis

**DOI:** 10.1101/2024.01.17.24301452

**Authors:** Camila Aparicio, Zachary I. Willis, Mari M. Nakamura, Joshua Wolf, Cordell Little, Gabriela M. Maron, Paul K. Sue, Brenda I. Anosike, Christine Miller, Laura L. Bio, Prachi Singh, Scott H. James, Carlos R. Oliveira, PIDS Pediatric COVID-19 Therapies Task Force

## Abstract

**Background:** Risk stratification is a cornerstone of the Pediatric Infectious Diseases Society COVID-19 treatment guidance. This systematic review and meta-analysis aimed to define the clinical characteristics and comorbidities associated with critical COVID-19 in children and adolescents.

**Methods:** Two independent reviewers screened the literature (Medline and EMBASE) for studies published through August 2023 that reported outcome data on patients aged _≤_21 years with COVID-19. Critical disease was defined as an invasive mechanical ventilation requirement, intensive care unit admission, or death. Random effects models were used to estimate pooled odds ratios (OR) with 95% confidence intervals (CI), and heterogeneity was explored through subgroup analyses.

**Results:** Among 10,178 articles, 136 studies met the inclusion criteria for review. Data from 70 studies, which collectively examined 172,165 children and adolescents with COVID-19, were pooled for meta-analysis. In previously healthy children, the absolute risk of critical disease from COVID-19 was 4% (95% CI, 1%–10%). Compared with no comorbidities, the pooled OR for critical disease was 3.95 (95% CI, 2.78–5.63) for presence of one comorbidity and 9.51 (95% CI, 5.62–16.06) for _≥_2 comorbidities. Key risk factors included cardiovascular and neurological disorders, chronic pulmonary conditions (excluding asthma), diabetes, obesity, and immunocompromise, all with statistically significant ORs >2.00.

**Conclusions:** While the absolute risk for critical COVID-19 in children and adolescents without underlying health conditions is relatively low, the presence of one or more comorbidities was associated with markedly increased risk. These findings support the importance of risk stratification in tailoring pediatric COVID-19 management.

**Summary:** This systematic review with meta-analysis integrated data from 136 studies (172,165 patients) and identified diabetes; obesity; immunocompromise; and cardiovascular, neurological, and pulmonary disease as predictors of severe pediatric COVID-19. The presence of multiple comorbidities increases the risk of critical outcomes.

## INTRODUCTION

While most pediatric COVID-19 cases are mild, a subset of children and adolescents experience severe outcomes [1]. Novel therapeutic and prophylactic agents have the potential to improve outcomes in these vulnerable children [2]. The Pediatric Infectious Diseases Society (PIDS) has established a COVID-19 Therapies Task Force that provides recommendations for preventing and managing SARS-CoV-2 in children and adolescents [3–5]. The latest guidance emphasizes the importance of risk stratification in clinical decision-making. Although previous reviews have identified comorbidities that put children at risk for worse outcomes [6–8], these were limited to studies from the early stages of the pandemic and focused on a narrow range of risk factors. As the pandemic has evolved, so has our understanding of how SARS-CoV-2 affects children. To develop effective risk stratification tools and therapeutic strategies, it is crucial to update the evidence base on the factors that predispose children and adolescents to severe disease. This systematic review and meta-analysis aimed to synthesize the best available evidence on the clinical characteristics and underlying health conditions associated with severe or critical pediatric COVID-19 outcomes.

## METHODS

A systematic review of the published literature was conducted following established PRISMA guidelines and included searching Medline and EMBASE for eligible studies published from inception through August 25, 2023. The search terms were selected in consultation with a clinical information specialist (Supplementary Methods). Using the Covidence platform, three investigators (CRO, CA, CL) independently screened the titles and abstracts of all retrieved citations to identify potentially eligible studies.

We selected studies that included patients aged _≤_21 years with confirmed COVID-19 and provided enough data to estimate the odds ratio (OR) of critical disease for a given risk factor. Studies defining critical disease as the need for invasive mechanical ventilation, admission to an intensive care unit, or death were considered. We excluded articles that were case reports or case series with fewer than thirty patients, those in languages other than English or Spanish, editorials, commentaries, opinions, or reviews. Studies that only provided data on post-acute sequelae of COVID-19, such as multisystem inflammatory syndrome in children and post-acute sequelae of SARS-CoV-2 infection, were also excluded, as were studies that used composite acute and post-acute outcomes that could not be disentangled.

The primary search strategy was supplemented by cross-checking reference lists from previous guidelines, systematic reviews, and meta-analyses. Further, articles recommended by subject matter experts on the PIDS Task Force were also considered for inclusion. Suggested references that did not meet the inclusion criteria but provided additional context, such as studies that only included children with a specific risk factor, were included in the narrative review but not synthesized as part of the meta-analysis. If there was an overlap in patient population between studies (i.e., used the same datasets or patient registries), the larger study was used in the meta-analysis.

Full-text reviews were conducted using standardized data extraction forms by two independent reviewers (CA, CL), and disagreements were resolved by discussion with a third (CRO). Quality assessment of each study was conducted using the Newcastle-Ottawa Scale [9]. Studies were weighted based on their variance, and random-effects models were used to compute pooled odds ratios (OR) with 95% confidence intervals (95%CI). Freeman–Tukey transformations were used to generate pooled prevalence estimates. Heterogeneity between studies was evaluated using the I^2^ statistic, and its source was explored with subgroup analyses when present. Two-sided P-values <.05 were defined as statistically significant. All analyses were conducted using STATA version 17.0. This study was exempt from ethics review as it used only previously published data. The study protocol was registered on PROSPERO (CRD 42023431457) and follows the PRISMA reporting guidelines.

## RESULTS

The initial search produced 10,178 articles. Following a screening of titles and abstracts, 655 articles were selected for full-text review. Out of these, 102 articles met the inclusion criteria. Through backward citation tracing and recommendations from subject matter experts, 34 additional articles were identified, yielding a total of 136 articles for the systematic review [6–8, 10–142]. The results of the search and study selection process are outlined in Figure 1. After excluding the single-arm or non-comparable studies, 70 studies were selected for the meta-analysis, which included data on 172,165 children from more than 20 countries. Supplemental Tables 1 and 2 summarize details of the studies included in the evidence base. Of the 70 studies included in the meta-analysis, 29 (41%) were judged to be of high quality using the Newcastle-Ottawa Scale (Figure 2). Uncertainty about the comparability of cohorts was the most commonly identified potential source of bias, given that analyses in only 23 (33%) studies controlled for other underlying comorbidities. Figure 3 provides an overview of the assessed risk factors, including sample sizes, pooled OR, and the I^2^ statistic. Further synthesis for each set of risk factors is provided in subsequent sections.

**Figure 1.**
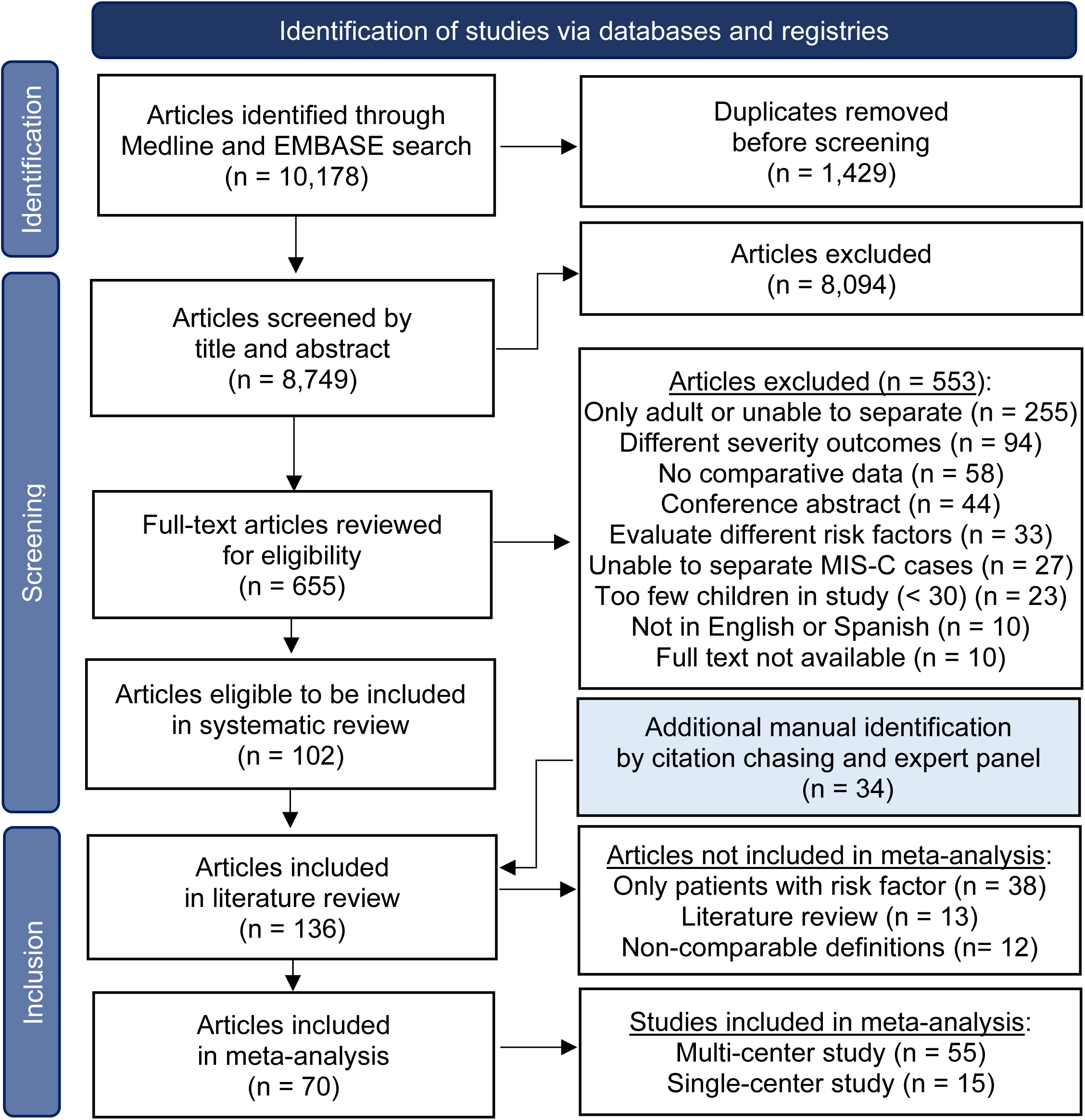
PRISMA Flow diagram of the literature search.

**Figure 2.**
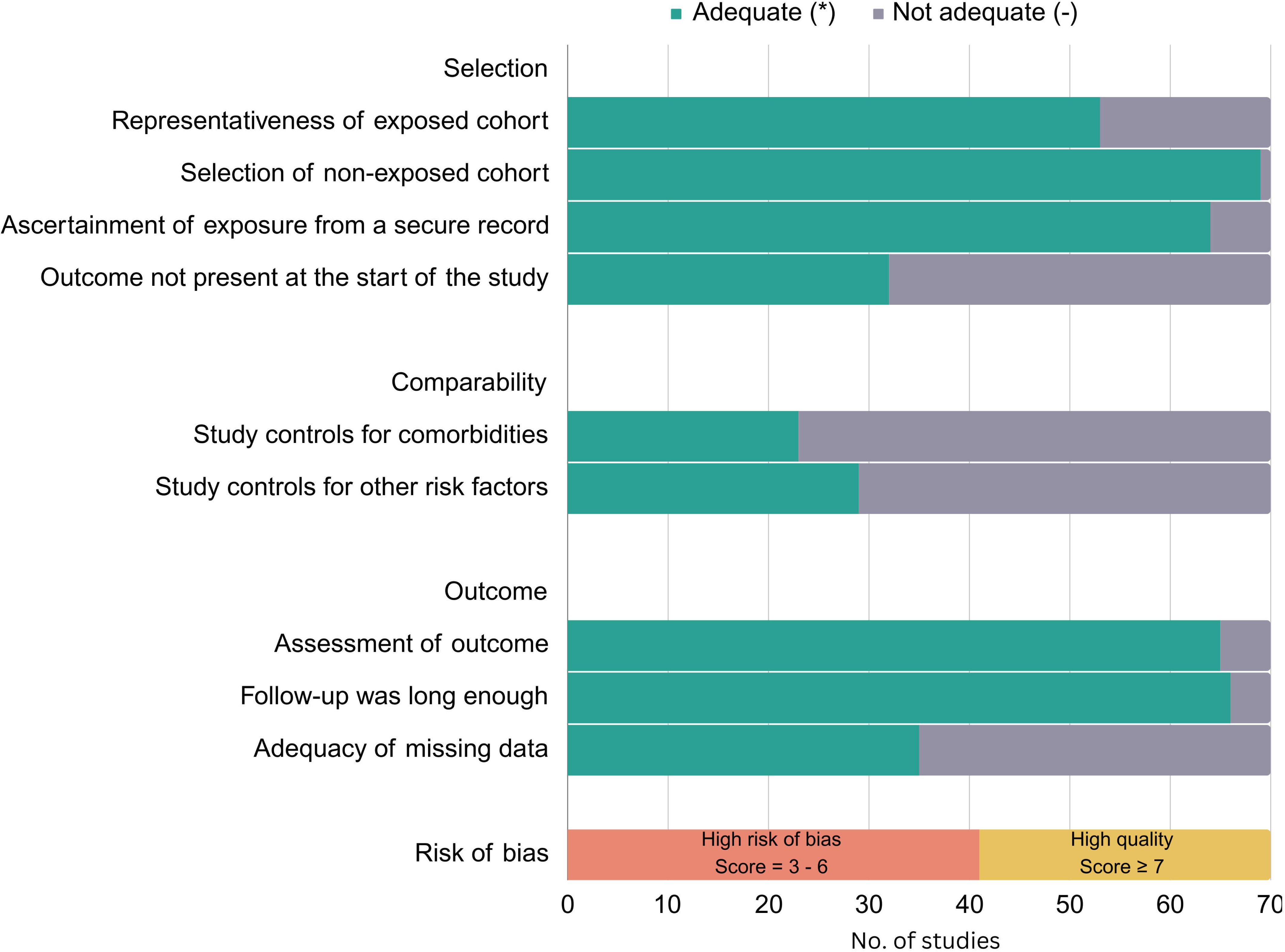
Risk of bias assessment for studies included in meta-analysis using the Newcastle-Ottawa Scale. Legend: Representativeness of exposed cohort: * indicates truly or somewhat representative of exposed cohort. Selection of non-exposed cohort: * indicates non-exposed cases drawn from same community as the exposed cohort. Ascertainment of exposure from secure record: * indicates taken from secure record. Outcome not present at the start of the study: * indicates yes. Study controls for comorbidities (most important factor): * indicates yes. Study controls for other risk factors: * indicates yes. Assessment of outcome: * indicated using record linkage. Follow-up was long enough: * indicates all included patients were followed up until discharge from hospital or for > 1 month. Adequacy of missing data: * Missing data unlikely to introduce bias (< 20). Orange bar: High risk of bias = 3 – 6 *; Yellow bar: High quality _≥_ 7 *

**Figure 3.**
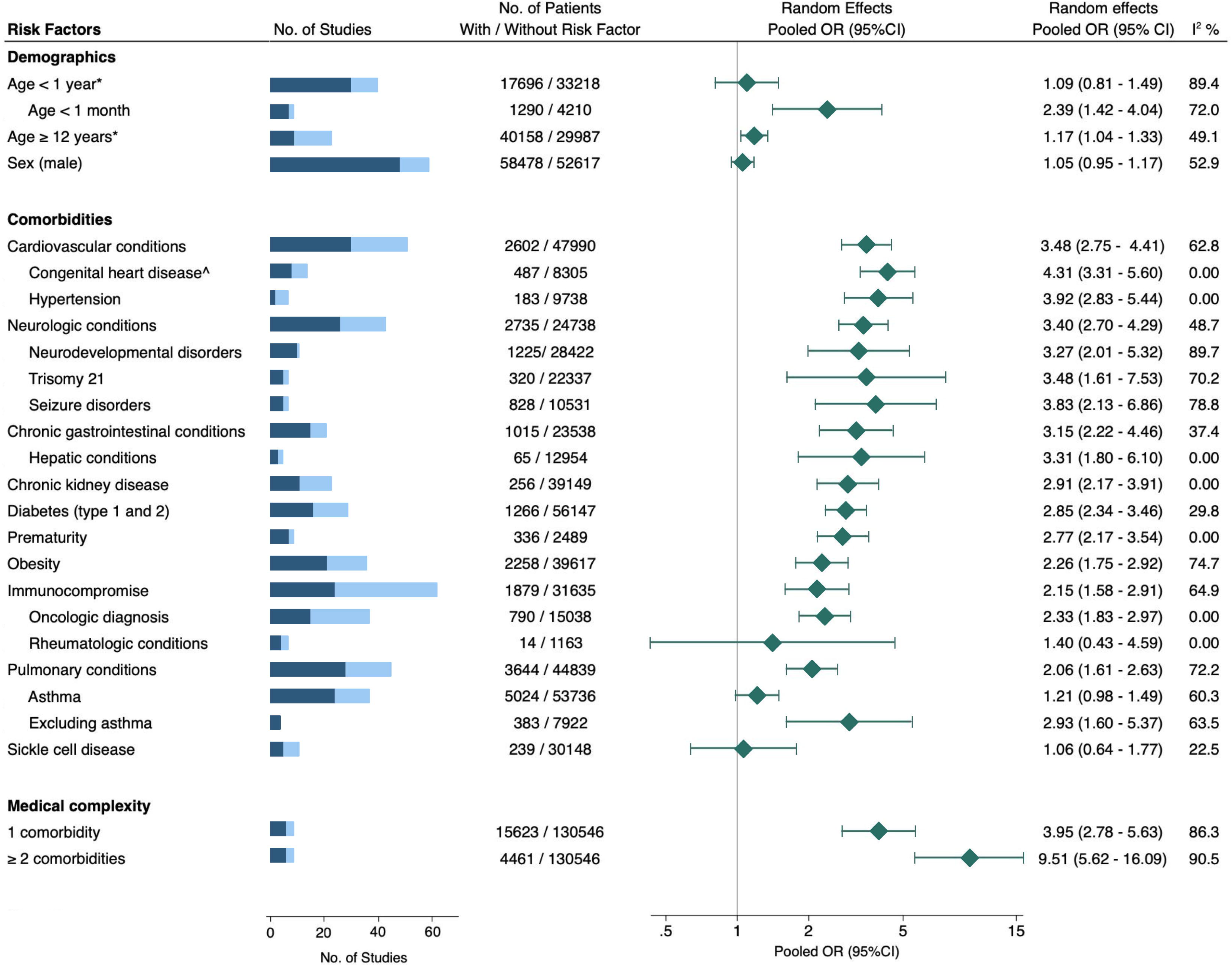
Association between risk factors and critical COVID-19 in children. Blue bar = total number of studies evaluated, with darker shade indicating the subset of studies included in the meta-analysis. Pooled OR (95%CI) = pooled odds ratio with 95% confidence interval, estimated using extracted data from published studies. * = Reference age 1-11 years. ^ = Fixed effects model used.

### Demographics

Of the 52 studies that considered age as a risk factor, 30 studies provided data on 17,696 children aged <1 year (10% critical; 95% CI, 6%–14%), 7 studies included information on 1,290 infants aged <1 month (20% critical; 95% CI, 8%–35%), and 9 studies had data on 40,158 children aged 12–21 years (19% critical; 95% CI, 8%–33%). While multiple studies suggested that age <1 year was a risk factor, we found this cutoff to be less consistently associated with increased risk and potentially influenced by younger infants (age <1 month) and prematurity, which were both stronger predictors of severity (Supplemental Figures 1–3). Increased odds of critical disease were also observed in adolescents relative to children aged 1–11 years, though the effect size was modest (OR 1.17; 95% CI, 1.04–1.33, Supplemental Figure 4). Evidence from 48 studies indicated that sex was not associated with an increased risk for progression to critical disease, with similar rates of critical outcomes in males and females (Supplemental Figure 5).

### Cardiovascular and pulmonary conditions

Cardiovascular conditions considered across 51 studies included congenital heart disease, hypertension, heart failure, cardiomyopathies, valvular disease, septal defects, myocarditis, arrhythmias, and pulmonary hypertension. The rate of critical disease among 2,372 children with cardiovascular disease in these studies was 30% (95% CI, 23%–37%), and the pooled OR was 3.60 (95% CI, 2.81–4.61). On subgroup analysis, there was an increased rate of critical disease in children with congenital heart disease (OR 4.31; 95% CI, 3.31–5.60) and hypertension (OR 3.92; 95% CI, 2.83–5.44) (Supplementary Figures 6-8).

A total of 45 studies were evaluated to assess the effect of pulmonary conditions. As a group, pulmonary conditions were a risk factor for severity, with a pooled critical disease rate of 24% (95% CI, 17%–33%) and an OR of 2.15 (95% CI, 1.66–2.75) relative to children without pulmonary conditions (Supplementary Figure 9). In a sub-analysis of 24 studies that investigated whether asthma was a predictor, the pooled effect size was non-significant. In contrast, in the studies that excluded asthma from the definition of pulmonary comorbidities, the effect size was robust and statistically significant (Supplemental Figures 10-11).

Few studies delved into other pulmonary conditions aside from asthma. One study, by Martin et al. found a statistically significant association between tracheostomy and severe disease, even after adjusting for other comorbidities (aOR 1.49; 95% CI, 1.08–2.04) [21]. In a multicenter study of 945 children with COVID-19, none of the 14 children with cystic fibrosis had severe disease, and five out of nine children with bronchopulmonary dysplasia required inpatient care [37].

Sleep apnea as a comorbidity was independently assessed by Graff et al. in their single-center cohort study and was associated with hospitalization but not respiratory support once hospitalized [99].

### Neurologic and psychiatric conditions

Pooling 26 studies involving 2,735 children with neurological conditions, the incidence of critical COVID-19 was 26% (95% CI, 16%–36%). These children had a threefold increased odds of critical illness compared to the general pediatric population (OR 3.40; 95% CI, 2.70–4.29; Supplemental Figure 12). Seizure disorder, in particular, showed a significant correlation with critical outcomes (Supplemental Figure 13), a relationship that persisted even in adjusted models (Supplemental Table 3). While a statistically significant correlation with critical disease was also seen with neurodevelopmental disorders, notable between-study heterogeneity was present (Supplemental Figure 14). This heterogeneity was primarily due to the largest study in this subgroup, which included 1,690 children with neurodevelopmental disorders and reported more hospitalizations but a lower-than-expected progression to critical illness (aRR 0.83; 95% CI, 0.70–0.98) in this cohort [107].

In a multicenter study that included 8,416 children with COVID-19 and mental health disorders, >90% had a mild or asymptomatic illness [14]. Parallel studies indicated that while conditions such as attention-deficit hyperactivity disorder, anxiety, and depression may be associated with an elevated risk of hospitalization, they do not necessarily correlate with increased likelihood of critical COVID-19 outcomes [12, 14, 107].

### Diabetes and Obesity

We synthesized findings from 36 studies to examine the association of diabetes (type 1 and/or 2) and obesity (defined as a body mass index or weight above the 95th percentile for age and sex) with severe COVID-19 in children. Data from 15 studies involving 1,112 children with diabetes showed a 26% rate of critical outcomes (95% CI: 15%–39%). Meanwhile, analyses of 18 studies with 2,258 children with obesity indicated a 32% incidence of critical disease (95% CI: 24%–40%). A pooled analysis revealed two-fold higher odds of critical disease in children with either condition (Supplemental Figures 15, 16). Subgroup analysis by study quality showed that diabetes and obesity are each independent predictors of severity, with six high-quality studies reporting robust effect sizes even after controlling for confounders (Supplemental Table 3). Notably, one study of 21,591 hospitalized COVID-19 patients found a quadrupled mortality risk for children with co-existing diabetes and obesity [143].

### Immunocompromise

Data on 1,879 children with COVID-19 who were classified as immunocompromised were extracted from 24 studies. Analysis of these data suggested that immunocompromise was a risk factor for critical COVID-19 (OR 2.15; 95% CI, 1.58–2.91; Supplemental Figure 17). These studies included a mixed group of immunocompromising conditions. However, restricting analyses to the 15 studies that only considered active oncologic diagnoses yielded a similar magnitude of risk (OR 2.33; 95% CI, 1.83–2.97). In contrast, studies of children with rheumatologic diseases did not show a significant correlation with critical COVID-19, though these studies were limited by small sample sizes (Supplemental Figures 18 and 19). Further analysis that considered data from an additional 7 case series, encompassing 1,278 children with rheumatologic conditions, revealed a pooled hospitalization rate of 6% (95% CI: 3%–10%) [62, 64, 65, 81, 95, 133, 134, 144–147]. The largest study in this subset, which included 607 children, did not find an independent correlation between hospitalization and the use of disease-modifying anti-rheumatic drugs or glucocorticoids [64].

### Chronic gastrointestinal conditions

Analysis of data from 15 studies, comprising 1,015 children with various chronic gastrointestinal (GI) diseases, showed an odds ratio (OR) of 3.15 for critical disease (95% CI: 2.22–4.46). The quality of these studies was moderate to low, and three studies that controlled for other comorbidities did not find GI diseases as a group to be independently associated with critical COVID-19 (Supplemental Table 3). The conditions included in the GI group included cirrhosis, pancreatic disease, esophageal disorders, inflammatory bowel disease, and dependency on parenteral or enteral feeding tubes. Of these, only feeding tube dependency and chronic liver disease were correlated with adverse outcomes (Supplemental Figures 20 and 21) [11, 110, 131, 141]. In contrast, no correlation with severity was noted for inflammatory bowel disease or esophageal disorders [66, 107].

### Chronic kidney disease

A total of 23 studies reported outcomes of children with chronic kidney disease (CKD) and COVID-19. Twelve studies were excluded from the subsequent meta-analysis because they included overlapping patients from larger studies or could not differentiate pre-existing kidney disease from kidney injury acquired during hospitalization. Of the remaining 11 studies, most (82%) did not find a statistically significant increased risk, but the sample sizes were generally small (Supplemental Figure 22). A data-linkage study by Martinez-Valdez et al. contributed the most weight to the meta-analysis, showing a 26% mortality rate among 149 hospitalized children with CKD [112], but carried a risk of bias because comorbidities were self-reported and not operationally defined. In contrast, two other studies that adjusted for comorbidities did not corroborate an association between CKD and severe COVID-19. In a sensitivity analysis that considered data from CKD registries that were not eligible for the meta-analysis, the pooled rate of critical COVID-19 in a sample of 566 children with CKD was 11% (95% CI, 5%–17%), with fatalities predominantly occurring in low-income countries and among those with multiple comorbidities [148–150].

### Sickle Cell Disease

Analysis from five studies of 239 children with sickle cell disease indicated a rate of critical disease of 15% (95% CI: 2%–35%), higher than otherwise reported baseline rates, but similar to comparison general pediatric study cohorts, with a pooled OR of 1.06 (CI 0.64-1.77).

Supplemental Figure 23). Although the studies included in the meta-analysis had small sample sizes, these estimates are consistent with data from a large case series involving 590 children with sickle cell disease, which also noted a 15% ICU admission rate [151].

### Medical Complexity

Six studies appraised medical complexity, defined as _≥_2 pre-existing conditions, as a risk factor for severity. A synthesis of these data, involving 15,623 pediatric COVID-19 patients, revealed that, on average, the absolute risk of critical COVID-19 outcomes in children without comorbidity was 4% (95% CI, 1%–10%). The presence of any one comorbidity was associated with 3.95 times higher odds of critical disease (Figure 4). For children who had _≥_2 chronic comorbidities, the odds of critical disease were even higher (OR 9.51; 95% CI, 5.62–16.06).

**Figure 4.**
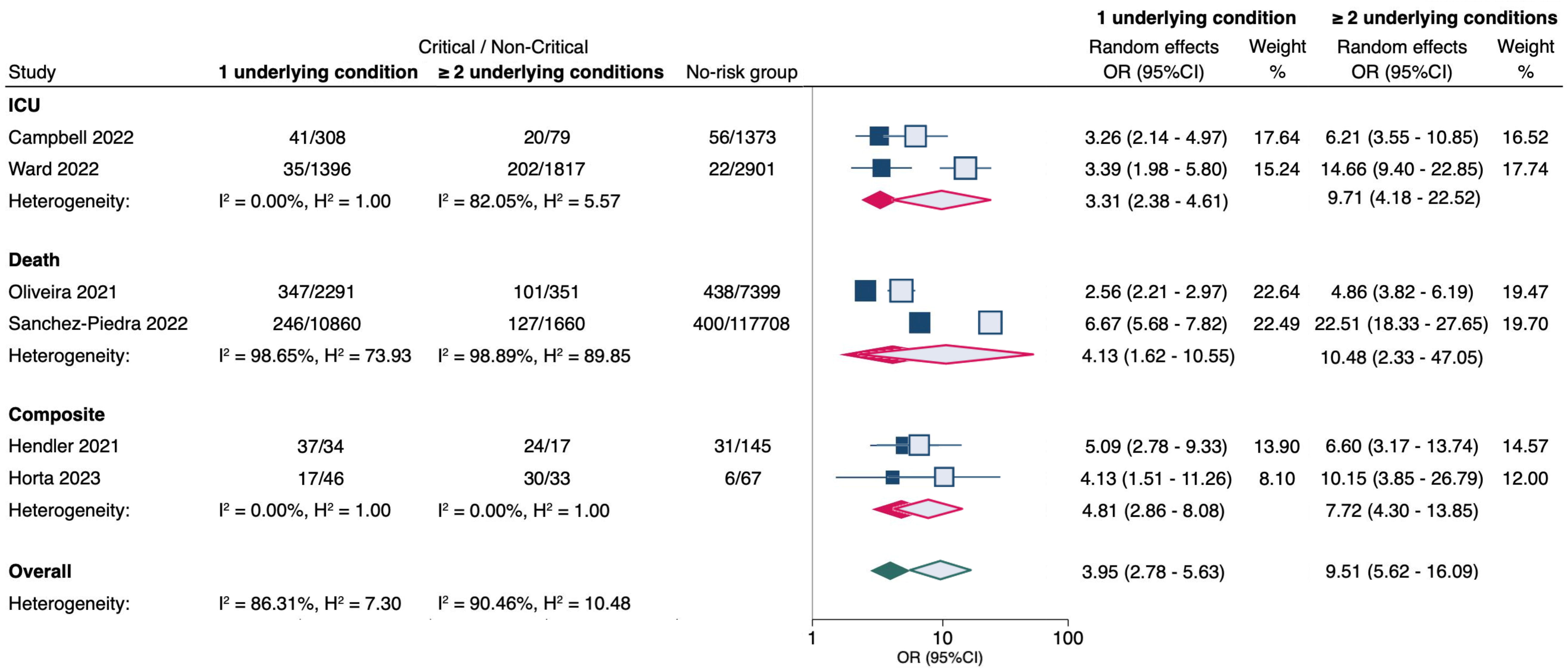
Meta-analysis of the association between the number of underlying comorbidities and critical COVID-19 in children. No risk group = patients with no underlying conditions. OR (95% CI) = odds ratio with 95% confidence interval, estimated using extracted data from published studies. Filled markers = 1 underlying condition; Unfilled markers = _≥_ 2 underlying conditions. ICU = admission to intensive care unit; Composite = combined critical outcomes (ICU, IMV, CVS, or death).

## DISCUSSION

In this systematic review and meta-analysis, we integrated data from 136 published manuscripts that collectively examined 172,165 children and adolescents with COVID-19. Based on our pooled data, we concluded that certain comorbidities are definite risk factors for critical COVID-19 outcomes. Specifically, cardiovascular and neurological disorders, chronic pulmonary conditions other than asthma, diabetes, obesity, and immunocompromise were all consistently associated with severe disease, exhibiting large effect sizes (OR >2.00) even when controlling for the presence of other comorbidities. While individual studies suggested potential associations between disease severity and conditions such as sickle cell disease, neuro-disabilities, and chronic kidney, rheumatologic, or gastrointestinal diseases, the available evidence was not robust enough to draw definitive conclusions. We found a dose-response relationship when considering the total number of comorbidities, wherein multiple pre-existing conditions correlated with increased odds of critical disease compared with a single condition.

We were able to corroborate certain findings of earlier evidence syntheses, but also contribute new insights and granularity into the conditions associated with COVID-19 severity in children. For example, in the most recent meta-analysis by Harwood et al., which included 57 studies and 21,549 patients, investigators reported that obesity was the only condition associated with increased odds of ICU admission and death independent of other comorbidities [6]. In contrast, our meta-analysis, which incorporated a substantially larger patient cohort and twice as many studies, further identified cardiovascular diseases, chronic pulmonary disorders, neurological conditions, and diabetes as independent predictors of severity. Some divergences from previous literature also emerged in these analyses. Where previous reviews indicated increased risks for both adolescents and children <1 year of age,[6] our data suggested that age was only a modest risk factor when no other risk factors were present, with the exception of age <1 month, for which evidence leaned towards increased severity.

The probability of critical illness from COVID-19 can be influenced by factors beyond those of pre-existing medical conditions. For instance, vaccination and immunity from prior infections can significantly mitigate risk in immunocompetent hosts [152–154]. Furthermore, the level of risk associated with specific medical conditions can vary depending on how well they are managed. For instance, there is evidence to suggest that diabetic children who maintain good glycemic control are at no greater risk than the general population [40, 78, 155, 156]. Similarly, children with congenital heart disease who have undergone surgical repair or no longer have a hemodynamically significant abnormality may be at lower risk [35, 157].

This study has some limitations to note. First, our findings are subject to the quality and biases of the included studies. We only analyzed observational studies, many of which were retrospective in nature. An inherent limitation of synthesizing data from non-randomized studies is the possibility that uncontrolled confounding could lead to biased pooled estimates. We attempted to mitigate this by focusing on high-quality studies that accounted for important confounders and only drawing inferences of definite risk when there was a sufficiently large magnitude of the effect (OR >2.00), as described in the GRADE conceptual framework [158]. Second, there was considerable heterogeneity across studies, particularly in how comorbidities were defined, introducing the possibility of misclassification bias. Lastly, the continuously evolving nature of the pandemic, with the introduction of novel variants, shifting disease- and vaccine-induced immunity, and the changing landscape of COVID-19 therapies, may affect the relevance of our conclusions over time.

The current management of COVID-19 in the pediatric population is a multifaceted issue, requiring a balanced assessment of the potential risks and benefits of various therapeutic agents, as well as a comprehensive evaluation of the myriad underlying risk factors that may predispose children to more severe disease. Although our study is subject to certain limitations, it contributes evidence on a number of risk factors that are clearly associated with a more severe trajectory of disease. Further research is needed to clarify the extent to which pre-existing immunity and the management of these comorbidities modify these risks.

## Data Availability

All data produced in the present work are contained in the manuscript.

## Contributors

The authors thank Mark J. Abzug, Monica Ardura, Juri Boguniewicz, Kathleen Chiotos, Kevin Downes, Stepen Grapentine, Christine Hanks, Molly Mcdonough, Adam L. Hersch, Sarah Heston, Diego R. Hijano, Charles W. Huskins, Candace L. Johnson, Sarah Hones, Elizabeth C. Lloyd, Christine MacBrayne, Theodore H. Morton, Rosemary M. Olivero, Rachel C. Orscheln, Hayden T. Schwenk, Vijaya L. Soma, Surabhi B. Vora, and Alpana Waghmare for their invaluable work.

