## Supplementary for "Risk Factors for Pediatric Critical COVID-19: A Systematic Review and Meta-Analysis"

**Risk Factors for Critical COVID-19 in Children and Adolescents:**

**A Systematic Review and Meta-Analysis**

**Supplementary material**

**APPENDIX**

[**Supplementary Figure 1**. Age < 1 year	51](#_Toc154081218)

[**Supplementary Figure 2**. Age < 1 month	52](#_Toc154081219)

### **Section 1: PIDS Pediatric COVID-19 Therapies Task Force Members**

| **Name** | **Institute** | **Location** |
| --- | --- | --- |
| Abzug, Mark J. | Department of Pediatrics, University of Colorado School of Medicine and Children’s Hospital Colorado | Aurora, CO |
| Anozike, Brenda I. | Department of Pediatrics, The Children’s Hospital at Montefiore and Albert Einstein College of Medicine | Bronx, NY |
| Aparicio, Camila | Department of Pediatrics, Yale University School of Medicine | New Haven, CT |
| Ardura, Monica I. | Department of Pediatrics, ID Host Defense Program, Nationwide Children's Hospital & The Ohio State University | Columbus, OH |
| Bio, Laura L. | Department of Pharmacy, Lucile Packard Children’s Hospital | Stanford, CA |
| Boguniewicz, Juri | Department of Pediatrics, University of Colorado School of Medicine and Children’s Hospital Colorado | Aurora, CO |
| Chiotos, Kathleen | Departments of Anesthesiology, Critical Care Medicine, and Pediatrics, Perelman School of Medicine at the University of Pennsylvania  Divisions of Critical Care Medicine and Infectious Diseases, Children’s Hospital of Philadelphia | Philadelphia, PA |
| Downes, Kevin | Department of Pediatrics, Perelman School of Medicine of the University of Pennsylvania Division of Infectious Diseases, Children’s Hospital of Philadelphia | Philadelphia, PA |
| Grapentine, Steven P. | Department of Pharmacy, University of California San Francisco Benioff Children's Hospital | San Francisco, CA |
| Hayes M, Molly | Center for Healthcare Quality & Analytics, The Children’s Hospital of Philadelphia | Philadelphia, PA |
| Hersh, Adam L. | Department of Pediatrics, Division of Infectious Diseases, University of Utah | Salt Lake City, UT |
| Heston, Sarah M. | Department of Pediatrics, Duke University School of Medicine | Durham, NC |
| Hijano, Diego R. | Department of Infectious Diseases, St. Jude Children's Research Hospital | Memphis, TN |
| Huskins, W. Charles | Department of Pediatric and Adolescent Medicine, Mayo Clinic | Rochester, MN |
| James, Scott H. | Department of Pediatrics, Division of Pediatric Infectious Diseases, University of Alabama at Birmingham | Birmingham, AL |
| Jones, Sarah | Department of Pharmacy, Boston Children’s Hospital | Boston, MA |
| Little, Cordell Z. | Department of Pediatrics, Yale University School of Medicine | New Haven, CT |
| Lockowitz, Christine R. | Department of Pharmacy, St. Louis Children's Hospital | St. Louis, MO |
| Lloyd, Elizabeth C. | Department of Pediatrics, University of Michigan | Ann Arbor, MI |
| MacBrayne, Christine | Department of Pharmacy, Children's Hospital Colorado | Aurora, CO |
| Maron, Gabriela M. | Department of Infectious Diseases, St. Jude Children's Research Hospital | Memphis, TN |
| Miller, Christine | Department of Pediatrics, Yale University School of Medicine | New Haven, CT |
| Morton, Theodore H. | Department of Pharmacy, St Jude’s Children’s Research Hospital | Memphis, TN |
| Nakamura, Mari M. | Antimicrobial Stewardship Program and Division of Infectious Diseases, Boston Children's Hospital | Boston, MA |
| Oliveira, Carlos R. | Department of Pediatrics, Yale University School of Medicine | New Haven, CT |
| Olivero, Rosemary M. | Department of Pediatrics and Human Development, Michigan State College of Human Medicine and Helen DeVos Children's Hospital of Corewell Health | Grand Rapids, MI |
| Orscheln, Rachel C. | Department of Pediatrics, Washington University | St. Louis, MO |
| Schwenk, Hayden T. | Department of Pediatrics, Stanford School of Medicine | Stanford, CA |
| Singh, Prachi | Department of Pediatrics, University of California San Francisco | San Francisco, CA |
| Soma, Vijaya L. | Department of Pediatrics, NYU Grossman School of Medicine | New York, NY |
| Sue, Paul K. | Department of Pediatrics, Columbia University | New York, NY |
| Vora, Surabhi B. | Department of Pediatrics, University of Washington School of Medicine, and Division of Infectious Diseases, Seattle Children’s Hospital | Seattle, WA |
| Willis, Zachary I. | Department of Pediatrics, University of North Carolina School of Medicine | Chapel Hill, NC |
| Wolf, Joshua | Department of Infectious Diseases, St. Jude Children's Research Hospital and Department of Pediatrics, University of Tennessee Health Science Center | Memphis, TN |

### **Section 2: Search Strategy and Terms**

**Risk factor**: "risk factors"[MeSH Terms] OR ("risk"[All Fields] AND "factors"[All Fields]) OR "risk factors"[All Fields] OR ("risk"[All Fields] AND "factor"[All Fields]) OR "risk factor"[All Fields]

**Prognosis**: "prognosis"[MeSH Terms] OR "prognosis"[All Fields] OR "prognoses"[All Fields]

**Predictor**: "predictor"[All Fields] OR "predictors"[All Fields]

**pediatric**: "paediatrics"[All Fields] OR "pediatrics"[MeSH Terms] OR "pediatrics"[All Fields] OR "paediatric"[All Fields] OR "pediatric"[All Fields]

**Children**: "child"[MeSH Terms] OR "child"[All Fields] OR "children"[All Fields] OR "child's"[All Fields] OR "children's"[All Fields] OR "childrens"[All Fields] OR "childs"[All Fields]

**COVID-19**: ("COVID-19" OR "COVID-19"[MeSH Terms] OR "COVID-19 Vaccines" OR "COVID-19 Vaccines"[MeSH Terms] OR "COVID-19 serotherapy" OR "COVID-19 serotherapy"[Supplementary Concept] OR "COVID-19 Nucleic Acid Testing" OR "covid-19 nucleic acid testing"[MeSH Terms] OR "COVID-19 Serological Testing" OR "covid-19 serological testing"[MeSH Terms] OR "COVID-19 Testing" OR "covid-19 testing"[MeSH Terms] OR "SARS-CoV-2" OR "sars-cov-2"[MeSH Terms] OR "Severe Acute Respiratory Syndrome Coronavirus 2" OR "NCOV" OR "2019 NCOV" OR (("coronavirus"[MeSH Terms] OR "coronavirus" OR "COV") AND 2019/11/01[PDAT] : 3000/12/31[PDAT]))

**SARS-CoV-2**: "sars-cov-2"[MeSH Terms] OR "sars-cov-2"[All Fields] OR "sars cov 2"[All Fields]

### **Section 3: Evidence Tables**

#### **Supplementary Table 1**. Characteristics of studies included in meta-analysis.

| **Study**  (Author year) | **Study design and setting** | **Subjects** | **Severity definition** | **Results: Risk factor**  (ES [95%CI, p – value]) | Risk factors reviewed |
| --- | --- | --- | --- | --- | --- |
| **Abdolsalehi 2023 [1]** | Single-center retrospective cross-sectional study conducted in Iran (Mar/20 - Aug/20) | < 18 years  n = 189 | ICU | Age < 1 year (ref. 8 - 18 years): OR 4.4 (1.6 - 11.5, p 0.002)  Underlying comorbidities: OR 3.2 (1.4 - 7.0, p 0.003)  Not associated: gender, BMI, underlying comorbidity + high BMI  *Adjusted for age, gender, and underlying disorders* | Sex |
| **Alharbi 2021 [2]** | Single-center retrospective cohort study conducted in Saudi Arabia (Apr/20 - Jul/20) | < 15 years  n = 65 | ICU | Not associated: sex, age, BMI, any comorbidity, cardiac disease, neurologic disorder, asthma, pulmonary disease, endocrine disease, gastrointestinal disease, hematological disease, metabolic disease, immunodeficiency | Age  Sex  Cardiovascular conditions  Neurologic conditions  GI and liver conditions |
| **Antúnez-Montes 2021 [3]** | Multi-center multinational cohort study (Jul/20 - Aug/20) | < 18 years  n = 96 | ICU | Combined COVID-19 and MIS-C cases (n = 409)  **COVID-19**: 314  **MIS-C**: 95  Preexisting medical conditions: 44.2% (n = 23) vs. 16.8% (n = 60), p < 0.001  Congenital syndromes: 9.6% (n = 5) vs. 0.3% (n = 1), p < 0.001  Immunologic diseases: 7.7% (n = 4) vs. 0.8% (n = 3)  Known immunodeficiency: 11.5% (n = 6) vs. 3.3% (n = 3)  Not associated: age, sex, congenital heart disease, neurologic disorders, immunosuppressive therapy, chemotherapy in the past, socioeconomic status | Data extracted from Harwood et al. individual patients’ data.  Sex  Neurologic conditions  Immunocompromise |
| **Araujo da Silva 2021 [4]** | Multi-center retrospective cohort study conducted in Brazil (Mar/20 - Nov/20) | < 18 years  n = 50 | ICU | Do not report individual OR. Combined with estimates of Harwood et al. individual patient meta-analysis. | Data extracted from Harwood et al. individual patients’ data.  Sex  Cardiovascular conditions  Neurologic conditions  Pulmonary conditions  Immunocompromise  GI and liver conditions |
| **Armann 2020 [5]** | Multi-center cohort study conducted in Germany (Mar/20 - May/20) | < 18 years  n = 102 | ICU | Do not report individual OR. Combined with estimates of Harwood et al. individual patient meta-analysis. | Data extracted from Harwood et al. individual patients’ data.  Age  Sex  Cardiovascular conditions  Neurologic conditions  Pulmonary conditions  Immunocompromise  Chronic kidney disease |
| **Banull 2022 [6]** | Single-center retrospective cohort study conducted in USA (Mar/20 - Feb/21) | < 25 years  n = 390 | ICU | Multivariate analysis  Hypothyroidism: aOR 11.077 (3.012 - 40.740)  Diabetes: aOR 8.227 (2.789 - 24.271)  Obesity: aOR 2.762 (1.295 - 5.890)  Not associated: adrenal insufficiency, overweight  *Adjusted for age, socioeconomic status, weight, hypothyroidism, and comorbidities* | Sex  Diabetes  Obesity |
| **Bayesheva 2021 [7]** | Multi-center retrospective cohort study conducted in Kazakhstan (Mar/20 - Jun/20) | < 19 years  n = 549 | Severe/Critical | **ICU**: 0.9% (n = 6)  **Death**: 0% (n = 0)  Age 5 - 12 years: 50% (n = 2/4 severe, p < 0.001  Any comorbidity: 50% (n = 2/4 severe, p < 0.001)  Cardiovascular disease: 25% (n = 1/4 severe, p < 0.001)  Hypertension: 25% (n = 1/4 severe, p < 0.001)  Not associated: sex, neonates or infants, young children, location of residence, diabetes, obesity, kidney disease, liver disease, chronic neurological disease, anemia, congenital malformation | Data extracted from Harwood et al. individual patients’ data.  Age  Sex  Cardiovascular conditions  Neurologic conditions  GI and liver conditions  Chronic kidney disease |
| **Bellino 2020 [8]** | Multi-center retrospective cohort study conducted in Italy (Feb/20 - May/20) | < 18 years  n = 3,836 | **Critical**: severe pneumonia, acute respiratory distress syndrome, septic shock, and/or multiple organ dysfunction requiring hospitalization in intensive care. | ICU - Age groups: p < 0.001  0 - 1 year: 2.6% (n = 5/528)  2 - 6 years: 9.5% (n = 8/659)  7 - 12 years: 1% (n = 1/1109)  13 - 17 years: 2.9% (n = 4/1540)  Death - Age groups: p < 0.03  0 - 1 year: 0.4% (n = 2/528)  2 - 6 years: 0.3% (n = 2/659)  7 - 12 years: 0% (n = 0/1109)  13 - 17 years: 0% (n = 0/1540) | Age  Sex  Cardiovascular conditions  Neurologic conditions  Pulmonary conditions  Immunocompromise |
| **Bhavsar 2021 [9]** | Multi-center retrospective cohort study conducted in USA (Mar/20 - May/20) | ≤ 21 years  n = 67 | **ICU** | Not associated: sex, ethnicity, age > 1 year vs. age < 1 year, smoking, BMI, asthma, chronic lung disease, oncologic diagnoses, cardiac disease, diabetes, neurologic disorders, rheumatologic disease, stem cell transplant | Age  Sex  Neurologic conditions  Pulmonary conditions  Immunocompromise |
| **Bolaños-Almeida 2021 [10]** | Multi-center retrospective cohort study conducted in Colombia (Jan/20 - Jun/20) | < 18 years  n = 597 | ICU or death | Age (ref. asymptomatic or mild)  Moderate: 3.1 ± 0.5 years (p < 0.001)  Severe (ICU): 4.5 ± 1.1 years (p < 0.001)  Death: 6.5 ± 1.9 years (p < 0.009)  Age (ref. recovered or outpatient)  Moderate (hospitalized): 6.2 ± 0.44 years (p < 0.001)  Severe (ICU): 4.88 ± 1.01 years (p < 0.001)  Death: 2.88 ± 1.9 years (p < 0.009)  No statistical difference between moderate, severe or death | Data extracted from Harwood et al. individual patients’ data.  Age  Sex  Neurologic conditions |
| **Bundle 2021 [11]** | Multi-center multinational retrospective cross-sectional study (Aug/20 - Oct/21) | < 18 years  n = 9,611 | ICU, ventilation, ECMO, or death | ICU  Any comorbidity: aOR 8.74 (6.22 – 12.27)  Age 1 – 4 years + any comorbidity: aOR 10.25 (5.70 – 18.45)  Age 5 – 11 years + any comorbidity: aOR 18.56 (10.12 – 34.06)  Age 12 – 17 years + any comorbidity: aOR 10.25 (7.55 – 13.90)  Sex (male): 0.09% vs. 0.07%, p < 0.05  Not associated: age < 1 year  *Adjusted for age group, country, sex, and infection period* | Age  Sex |
| **Cairoli 2020 [12]** | Single-center cohort study conducted in Argentina (Apr/20 - Jun/20) | < 21 years  n = 578 | - | - | Age  Sex  Cardiovascular conditions  Neurologic conditions  Pulmonary conditions  Immunocompromise  GI and liver conditions  Chronic kidney disease |
| **Campbell 2022 [13]** | Multi-center retrospective cohort study conducted in USA (Apr/20 - Apr/21) | 12 - 17 years  n = 1,877 | ICU between days 0 – 28 | Multivariate analysis  Immunocompromising condition: aOR 2.2 (1.0 - 4.8)  Obesity: aOR 4.7 (2.8 - 7.6)  Pulmonary disease (excl mild asthma): aOR 2.5 (1.4 - 4.5)  Heart disease: aOR 2.3 (1.0 - 5.3, p < 0.05)  Not associated: sex (female), sickle cell disease, CKD, diabetes, neurologic disease  *Adjusted for race/ethnicity and comorbidities* | Sex  Medical complexity  Cardiovascular conditions  Neurologic conditions  Pulmonary conditions  Immunocompromise  Sickle cell disease  Chronic kidney disease  Diabetes  Obesity |
| **Chao 2020 [14]** | Single-center retrospective cross-sectional study conducted in USA (Mar/20 - Apr/20) | ≤ 21 years  n = 46 | ICU | Not associated: age, sex, weight, BMI, obesity, asthma, immunosuppressed, seizure disorder, malignancy, heart disease | Age  Sex  Cardiovascular conditions  Neurologic conditions  Pulmonary conditions  Immunocompromise  Obesity |
| **Choudhary 2022 [15]** | Multi-center cross-sectional study conducted in USA (Jul/21 - Aug/21) | < 21 years  n = 759 | Receiving HFNC, BiPAP, CPAP, or IV | < 1 year (n = 176)  ≥ 2 underlying conditions: PR 1.82 (1.04 - 2.18)  RSV coinfection: PR 3.64 (2.5 - 5.3)  Bacterial coinfection: PR 1.88 (1.21 - 2.94)  1 - 4 years (n = 122)  RSV coinfection: PR 1.96 (1.31 - 2.93)  5 - 11 years (n = 143)  Obesity: PR 2.2 (1.25 - 3.87)  GI/hepatic disease: PR 2.32 (1.26 - 4.27)  Feeding tube dependent: PR 2.77 (1.54 - 4.96)  Bacterial coinfection: PR 2.34 (1.36 - 4.02)  12 - 17 years (n = 272)  1 underlying condition: PR 2.95 (1.29 - 6.75)  ≥ 2 underlying conditions: PR 3.19 (1.41 - 7.2)  Obesity: PR 2.48 (1.74 - 7.2)  Feeding tube dependent: PR 1.55 (1.08 - 2.23)  Neurologic or developmental condition: PR 1.4 (1.05 - 1.88)  Not associated: preterm, cardiovascular disease, congenital heart disease, immunologic condition or malignancy, respiratory condition, asthma, other congenital anomaly, wheelchair dependent, other viral coinfection | Age |
| **de Ceano-Vivas 2020 [16]** | Single-center retrospective cohort study conducted in Spain (Mar/20 - Apr/20) | < 18 years  n = 33 | ICU | **ICU**: 15% (n = 5)  - 3/5 due to COVID-19  - 1/5 hypertensive crisis  - 1/5 diabetic ketoacidosis  **Death**: 3% (n = 1)  - 5-month-old infant with dilated cardiomyopathy and Hurler’s disease | Data extracted from Harwood et al. individual patients’ data.  Age  Sex  Neurologic conditions |
| **Desai 2020 [17]** | Multi-center retrospective cohort study conducted in USA (Jan/20 - May/20) | < 18 years  n = 293 | Mechanical ventilation or death | Inpatient  Disease of the respiratory system: 86% vs 53% (p 0.001)  Glucocorticoids: 36% vs. 12% (p 0.002)  Not associated: age, sex, congenital heart disease, other forms of heart disease, cystic fibrosis, asthma, BPD, neoplasms, overweight/obesity, diabetes insipidus, diabetes mellitus, endocrine/nutritional/metabolic diseases, neuromuscular scoliosis, congenital malformations/deformations/abnormalities, disease of the nervous system, disease of musculoskeletal system and connective system | Age  Sex  Cardiovascular conditions  Neurologic conditions  Pulmonary conditions  Immunocompromise |
| **Du 2020 [18]** | Single-center retrospective cohort study conducted in China (Jan/20 - Feb/20) | < 16 years  n = 182 | ICU + respiratory failure requiring mechanical ventilation, shock, or complications with other organ failures | Allergic conditions vs. No allergic conditions  Not associated: allergic conditions | Age  Sex |
| **Farrar 2022 [19]** | Multi-center prospective cohort study conducted in Canada (Apr/20 - May/21) | < 18 years  n = 330 | ICU, ventilatory support or CVS, select organ system complication, or death. | Multivariate analysis  < 1 year + prematurity: aRR 3.47 (1.69 - 7.09)  < 1 month: aRR 2.55 (1.15 - 5.64)  Bronchopulmonary dysplasia: aRR 2.39 (1.37 - 4.18)  Uncontrolled asthma: aRR 2.24 (1.54 - 3.27)  Epilepsy: aRR 2.08 (1.44 - 2.99)  Genetic/chromosomal disorders: aRR 2.04 (1.33 - 3.14)  Any technology dependence aRR 2.01 (1.37 - 2.95)  BMI Z-score ≥ 3: aRR 1.9 (1.1 - 3.28)  Any neurologic/neurodevelopmental condition: aRR 1.84 (1.32 - 2.57)  Parenteral nutrition: aRR 1.69 (1.01 - 2.83)  Chronic encephalopathy: aRR 1.67 (1.05 - 2.64)  Any pulmonary condition: aRR 1.63 (1.12 - 2.39)  Not associated: sex, any concomitant infection, timing of hospitalization, chronic comorbid conditions (non-complex, complex), congenital heart disease, obesity, cerebral palsy, BMI Z-score 2 - < 3, any neurologic condition, diabetes, metabolic disease, chronic renal disease, GI/liver disease, malignancy, immunocompromising medication, immunosuppression, asthma with controller medication, any hematologic disease, sickle cell disease  *Adjusted for age sex, concomitant infections, timing of hospitalization, and chronic conditions (non, non-complex, complex)* | Age  Sex  Prematurity  Cardiovascular conditions  Neurologic conditions  Pulmonary conditions  Sickle cell disease  GI and liver conditions  Obesity |
| **Farzan 2021 [20]** | Multi-center retrospective cohort study conducted in USA (Feb/20 - Feb/22) | 2 - 18 years  n = 38 | ICU | Asthma vs. Non-asthma  Not associated: asthma | Pulmonary conditions |
| **Fattahi 2022 [21]** | Multi-center retrospective cohort study conducted in Iran (Feb/20 - Feb/22) | < 18 years  n = 645 | ICU or death | Multivariate analysis  **ICU**  Sex (male): aOR 2.7 (1.2 - 6.5)  Comorbidities: aOR 2.8 (1.05 - 7.3)  Not associated: age  **Death**  Comorbidities: aOR 3.8 (1.2-12.1)  Not associated: sex, age  *Adjusted for age, sex, and comorbidities* | Age  Sex |
| **Fernandes 2021 [22]** | Multi-center retrospective cohort study conducted in USA (Mar/20 - May/20) | < 23 years  n = 143 | ICU admission for ≥ 48 hours | Multivariate analysis  Age (per 1 year decrease): aOR 1.09 (1.02 - 1.16, p 0.01)  Obesity: aOR 3.39 (1.26 - 9.1, p 0.02)  Not associated: BMI, sex, medical complexity  *Adjusted for hospital site, age, obesity, WBC count, ethnicity, oxygen saturation, medical complexity (comorbidities), and rx bilateral infiltrates* | Sex  Obesity |
| **Fisler 2020 [23]** | Single-center retrospective cohort study conducted in USA (Feb/20 - Apr/20) | < 21 years  n = 77 | ICU | Age ≥ 12 years 66.7% (n = 20/30 ICU admissions, p 0.015)  Not associated: any comorbidity, BMI | Age  Sex  Prematurity  Cardiovascular conditions  Pulmonary conditions  Immunocompromise  Chronic kidney disease  Diabetes  Obesity |
| **Funk 2021 [24]** | Multi-center multinational prospective cohort study (Mar/20 - Jun/21) | < 18 years  n = 3,221 | Cardiac, infectious, neurologic, or respiratory complications, and death, or performance of chest drainage, CVS, positive pressure ventilation (inv/non inv), and hemodialysis. | Age 5 - 9 years (ref. < 1 year): aOR 1.6 (1.09 - 2.34, p 0.02)  Age 10 - 17 years (ref. < 1 year): aOR 2.39 (1.38 - 4.14, p 0.002)  Chronic condition (self-reported): aOR 2.34 (1.59 - 3.44, p < 0.001)  Previous pneumonia: aOR 3.15 (1.83 - 5.42, p < 0.001)  Not associated: sex, age 1 - 2 years, age 2 - 4 years, asthma  *Adjusted for country and time period of enrollment* | Age  Sex |
| **Giacomet 2020 [25]** | Multi-center retrospective cohort study conducted in Italy (Feb/20 - May/20) | < 18 years  n = 119 | ICU | Not associated: age, sex, comorbidities, chronic cardiac conditions, GI disorder, obesity, CKD, chronic neurologic disease, immunologic conditions | Sex  Cardiovascular conditions  Neurologic conditions  Immunocompromise  GI and liver conditions  Chronic kidney disease  Obesity |
| **Gotzinger 2020 [26]** | Multi-center multinational prospective cohort study (Apr/20) | < 19 years  n = 582 | ICU | Multivariate analysis  Age < 1 month: aOR 5.06 (1.72 - 14.87)  Sex (Male): aOR 2.12 (1.06 - 4.21, p 0.033)  Comorbidities: aOR 3.27 (1.67 - 6.42, p 0.0015)  Not associated: chronic pulmonary diseases, congenital heart disease, neurological disorders, malignancy, viral coinfection, chromosomal abnormality, CKD, malignancy, immunosuppressive therapy, known immunodeficiency, chemotherapy in past 6 months  *Adjusted for age, sex, pre-existing medical conditions, signs and symptoms at presentation, radiological findings, and viral co-infection* | Age  Sex  Cardiovascular conditions  Neurologic conditions  Pulmonary conditions  Immunocompromise  GI and liver conditions  Chronic kidney disease |
| **Graff 2021 [27]** | Single-center retrospective cohort study conducted in USA (Mar/20 - Jul/20) | < 21 years  n = 66 | ICU | Obstructive sleep apnea: OR 4.7 (1.0 - 20.8, p.04)  Not associated: age, sex, pulmonary comorbidity, asthma, GI comorbidity, endocrine comorbidities, diabetes/prediabetes, prematurity, number or comorbidities, BMI, overweight/obesity | Age  Sex  Prematurity  Neurologic conditions  Pulmonary conditions  Immunocompromise  GI and liver conditions  Diabetes  Obesity |
| **Gujski 2021 [28]** | Multi-center retrospective cohort study conducted in Poland (Mar/20 - Dec/20) | < 20 years + adults  n = 5879 pediatric | Death | - | Sex |
| **Gupta 2021 [29]** | Multi-center retrospective cohort study conducted in India (Mar/20 - May/20) | < 21 years + adults  n = 4526 pediatric | Death | - | Sex |
| **Hendler 2021 [30]** | Multi-center retrospective cohort study conducted in Brazil (Mar/20 - Dec/20) | < 19 years  n = 288 | ICU, IMV, or death | Multivariate analysis  Age < 1 month (ref. 2 - 5 years): aOR 9.52 (3.01- 30.08)  1 comorbidity: aOR 5.27 (2.69 - 10.34)  ≥ 2 comorbidities: aOR 9.93 (4.45 - 22.17)  Not associated: age 1 month - 1 year, age 6 - 10 years, age > 11 years  *Adjusted for categorized age (<1 month, 1 month-1 year, 2-5 years, 6-10 years, and 11-18 years), race/ethnicity (White, non-White, and non-Black, or Black), and presence of one underlying condition, two or more underlying conditions vs. no underlying conditions*. | Age  Medical complexity  Cardiovascular conditions  Neurologic conditions  Pulmonary conditions  Immunocompromise |
| **Hillesheim 2020 [31]** | Multi-center prospective cross-sectional study conducted in Brazil (Mar/20 - Sep/20) | < 20 years  n = 6,989 | Death | Age < 1 year: lethality 14.2% (n = 191)  Age 1 - 4 years: lethality 6.7% (n = 129)  Age 5 - 9 years: lethality 6.3% (n = 68)  Age 10 - 14 years: lethality 8.9% (n - 85)  Age 15 - 19 years: lethality 11% (n = 188)  Sex (Female): lethality 9.7% vs. 9.2%  Indigenous: lethality 23%  Rural area: 18.1%` | Age  Sex |
| **Horta 2023 [32]** | Multi-center retrospective cohort study conducted in Brazil (Feb/20 - Nov/21) | < 19 years  n = 199 | ICU, IMV, or death | Any comorbidity: OR 6.59 (2.59 - 20.04, p ≤ 0.001)  1 comorbidity: OR 4.08 (1.4 - 13.64, p 0.005)  ≥ 2 comorbidities: OR 9.96 (3.62 - 32.27, p ≤ 0.001)  Asthma: OR 2.32 (1.16 - 4.66, p 0.01)  Neurological diseases: OR 4.7 (2.14 - 10.47, p ≤ 0.001)  Gastrointestinal diseases: OR 9.24 (3.34 - 28.47, p ≤ 0.001)  Congenital or chromosomic abnormalities: OR 7.73 (2.74 - 24.13, p ≤ 0.001)  Heart failure: OR 3.52 (1.2 - 10.48, p 0.01)  Not associated: age, sex | Sex  Medical complexity  Cardiovascular conditions  Neurologic conditions  Pulmonary conditions  GI and liver conditions |
| **Kainth 2020 [33]** | Single-center retrospective cohort study conducted in USA (Jan/20 - Apr/20) | < 22 years  n = 65 | ICU, IMV with or without use CVS, and renal replacement therapy (RRT), or death | Healthy infants vs. Healthy children vs. Immunocompromised children vs. Chronically ill children  Not associated: immunocompromised children, chronically ill children | Age  Cardiovascular conditions  Neurologic conditions  Pulmonary conditions  Immunocompromise |
| **Kalyanaraman 2020 [34]** | Multi-center cohort study conducted in USA (Mar/20 - Apr/20) | < 19 years  n = 32 | Death | Do not report individual OR. Combined with estimates of Harwood et al. individual patient meta-analysis. | Data extracted from Harwood et al. individual patients’ data.  Immunocompromise |
| **Kompaniyets 2021 [35]** | Multi-center cross-sectional study conducted in USA (Mar/20 - Jan/21) | < 19 years  n = 43,465 | ICU, IMV, or death | Multivariate analysis  Type 1 diabetes: aRR 2.38 (2.06-2.76)  Cardiac and circulatory congenital anomalies: aRR 1.72 (1.48 - 1.99)  Epilepsy and/or convulsions: aRR 1.71 (1.41 - 2.08)  Obesity: aRR 1.42 (1.22 - 1.66)  Essential hypertension: aRR 1.39 (1.19 - 1.63)  Sleep/wake disorders (including sleep apnea): aRR 1.26 (1.09 - 1.45)  Other specified status: aRR 1.25 (1.07 - 1.47)  Non complex chronic diseases: aRR 1.95 (1.69 - 2.26)  Complex Chronic Diseases: aRR 2.86 (2.47 - 3.32)"  Not associated: type 2 diabetes, tobacco-related disorders, asthma, esophageal disorders, anxiety and fear-related disorders, headache including migraine, depressive disorders, other congenital anomalies, neurodevelopmental disorders, trauma and stressor-related disorders, other upper respiratory disease  Age < 1 year  Cardiac and congenital anomalies: aRR 1.89 (1.48 - 2.41)  Prematurity: aRR 1.83 (1.47 - 2.29)  Age 2 - 5 years  Epilepsy and/or convulsions: aRR 1.95 (1.4 - 2.74)  Cardiac and circulatory congenital anomalies: aRR 1.5 (1.05 - 2.16)  Age 6 - 11 years  Epilepsy and/or convulsions: aRR 1.54 (1.05 - 2.26)  Sleep/wake disorders: aRR 1.82 (1.35 - 2.45)  12 - 18 years  Type 1 diabetes: aRR 2.47 (2.12 - 2.87)  Epilepsy and/or convulsions: aRR 1.89 (1.53 - 2.34)  Obesity: aRR 1.57 (1.32 - 1.85)  Essential hypertension: aRR 1.23 (1.01 - 1.51)  Asthma: aRR 1.17 (1.02 - 1.33)  *Adjusted for frequent (ie, prevalence >0.7%) underlying medical conditions, age group, sex, race/ethnicity, payer type, hospital urbanicity, hospital US Census region, admission month, and admission month squared.* | Age  Sex  Prematurity  Cardiovascular conditions  Neurologic conditions  Pulmonary conditions  GI and liver conditions  Diabetes  Obesity |
| **Kufa 2022 [36]** | Multi-center retrospective cohort study conducted in South Africa (Jan/20 - Sep/20) | < 18 years  n = 2,007 | Death | Multivariate analysis  Age < 1 year (ref. 1 - 4 years): aOR 4.11 (1.08–15.54)  Age 10–14 years (ref. 1 - 4 years): aOR 4.20 (1.07–16.44)  Age 15–17 years (ref. 1 - 4 years): aOR 4.86 (1.28–15.81)  Sex (Male): aOR 2.18 (1.08–4.40)  Admission at a public hospital: aOR 5.07 (2.01–12.76)  ≥1 non-communicable underlying medical condition: aOR 12.09 (4.19–34.89)  Not associated: ethnicity, province, month, HIV infection, past tuberculosis  *Adjusted for age, birth sex, admission to public or private sectors, and presence of underlying conditions* | Age  Sex |
| **Leung 2023 [37]** | Multi-center retrospective cohort study conducted in Brazil (Mar/20 - Nov/21) | < 18 years  n = 14,686 | Severe illness and death | Trisomy 21 vs. No trisomy 21  Multivariate analysis  Trisomy 21: aOR 2.06 (1.39–3.01)  Cardiovascular diseases: aOR 3.04 (2.38–3.87)  Hematologic disease: aOR 1.54 (1.03 - 2.26)  Liver diseases: aOR 4.15 (2.24–7.53)  Neurologic diseases: aOR 3.23 (2.62–3.96)  Pulmonary disease: aOR 2.04 (1.44 - 2.84)  Immunocompromise: aOR 2.76 (2.14 - 3.53)  Renal diseases: aOR 1.90 (1.29–3.01)  Obesity: aOR 2.30 (1.59–3.25)  Protector  Asthma: aOR 0.49 (0.34 - 0.62)  Vaccination against influenza: aOR 0.52 (0.37 - 0.71)  Not associated: age, sex  *Adjusted for demographic factors, underlying conditions other than Down syndrome, and intervention* | Neurologic conditions |
| **Madani 2021 [38]** | Multi-center retrospective cross-sectional study conducted in Iran (Feb/20 - Nov/20) | < 19 years  n = 6,610 | Death | Multivariate analysis  Cardiovascular disease: aOR 3.27 (2.09 - 5.11, p < 0.001)  Diabetes: aOR 2.5 (1.38 - 4.55, p 0.003)  Chronic pulmonary disease: aOR 3.21 (1.59 - 6.47, p 0.001)  Liver dysfunction: aOR 3.42 (1.75 - 6.67, p < 0.001)  Kidney dysfunction: aOR 3.42 (1.75 - 6.67, p < 0.001)  Protector  Age 1 - 5 years (ref. < 1 year): aOR 0.62 (0.44 - 0.87, p 0.006)  Age 6 - 12 years (ref. < 1 year): aOR 0.67 (0.47 - 0.96, p 0.029)  Not associated: age 13 - 17 years, sex, malignancies, immunodeficiency disorders  *Adjusted for age, sex, and all comorbidities.* | Cardiovascular conditions  Pulmonary conditions  Immunocompromise  GI and liver conditions  Chronic kidney disease  Diabetes |
| **Mania 2021 [39]** | Single-center retrospective cohort study conducted in Poland (Mar/20 - Apr/21) | IQR 15 - 152 months  n = 332 | ICU | Any comorbidity: 80% (n = 4/5 admitted to ICU, p < 0.00001)  Complex comorbidities: 60% (n = 3/5 admitted to ICU, p < 0.00001)  Genetic syndromes: 20% (n = 2/5 admitted to ICU, p < 0.00001)  Neurological disorders: 20% (n = 2/5 admitted to ICU, p < 0.00001)  Not associated: age, sex, asthma, cardiovascular disease, immune deficiency, obesity, diabetes mellitus, hypertension, coinfections | Sex  Cardiovascular conditions  Neurologic conditions  Pulmonary conditions  Immunocompromise  Diabetes  Obesity |
| **Martinez-Valdez 2022 [40]** | Multi-center retrospective cross-sectional study conducted in Mexico (Mar/20 - Sep/21) | < 18 years  n = 9,446 | Death | Age < 1 year (ref. 10 - 17 years): OR 16.67 (14.13 - 19.66, p 0.020)  Age 1 - 5 years (ref. 10 - 17 years): OR 3.34 (2.82 - 3.96, p 0.00)  Not associated: age 6 - 9 years  < 1 year  Immunosuppression: OR 7.02 (3.1 - 15.5)  Other comorbidity: OR 8.2 (4.95 - 13.6)  CKD: OR 4.8 (1.31 - 17.5)  Cardiovascular disease: OR 13.85 (7.8 - 24.3)  High blood pressure: OR 4.81 (3.1 - 7.3)  Diabetes: OR 4.41 (2.6 - 7.3)  1 - 2 comorbidities: OR 41.3 (22.3 - 76.4)  3 - 4 comorbidities: OR 41.3 (22.3 - 76.4)  Not associated: sex, COPD  1 - 5 years  Immunosuppression: OR 28.43 (15.8 - 50.9)  CKD: OR 12.86 (3.8 - 42.9)  Cardiovascular disease: OR 32.4 (18.8 - 55.8)  High blood pressure: OR 11.57 (4.9 - 27.1)  Diabetes: OR 7.47 (2.2 - 24.2)  1 - 2 comorbidities: OR 74.39 (51.71 - 107)  3 - 4 comorbidities: OR 354.47 (168.5 - 745.6)  Not associated: sex, obesity, COPD, asthma  6 - 9 years  Immunosuppression: OR 77.61 (42.5 - 141.6)  CKD: OR 22.13 (5.2 - 93.9)  Cardiovascular disease: OR 10.72 (2.5 - 44.5)  High blood pressure: OR 14.42 (3.4 - 60.3)  Obesity: OR 5.39 (2.4 - 11.7)  1 - 2 comorbidities: OR 40.48 (23.3 - 70.0)  3 - 4 comorbidities: OR 628.92 (261.4 - 1512.7)  Not associated: sex, diabetes, COPD, asthma  10 - 17 years  Immunosuppression: OR 72.23 (50.5 - 103.2)  CKD: OR 63.87 (42.2 - 96.5)  Cardiovascular disease: OR 19.26 (11.5 - 32.2)  High blood pressure: OR 10.10 (5.5 - 18.5)  Obesity: OR 5.18 (3.8 - 6.9)  Diabetes: OR 16.75 (10.5 - 26.5)  COPD: OR 8.32 (2.0 - 33.9)  1 - 2 comorbidities: OR 51.28 (37.2 - 70.5)  3 - 4 comorbidities: OR 848.1 (543 - 1324.6)  Not associated: sex, asthma | Age  Sex  Medical complexity  Cardiovascular conditions  Pulmonary conditions  Immunocompromise  Chronic kidney disease  Diabetes  Obesity |
| **Moreira 2021 [41]** | Multi-center retrospective cohort study conducted in USA (mar/20 - Jul/20) | < 20 years  n = 20,096 | Death | Multivariate analysis  Underlying medical condition: OR 8.8 (3.7 - 21.1, p < 0.001)  Not associated: age 0 - 9 years (ref. 10 - 19 years), sex  *Adjusted for age, sex, race/ethnicity, and comorbidity* | Sex |
| **Murillo-Zamora 2022-1 [42]** | Multi-center retrospective cohort study conducted in Mexico (Mar/20 - Jun/21) | < 16 years  n = 1,423 | Death | Multivariate analysis  Immunosuppression: HR 2.7 (1.38 - 5.3, p < 0.001)  Not associated: sex, age, obesity, diabetes mellitus, asthma, cardiovascular disease, CKD  *Adjusted for sex, age, and immunosuppression* | Sex  Cardiovascular conditions  Pulmonary conditions  Immunocompromise  Chronic kidney disease  Diabetes  Obesity |
| **Murillo-Zamora 2022-2 [43]** | Multi-center retrospective cohort study conducted in Mexico (Nov/21 - Jan/22) | < 10 years  n = 1,601 | Death | Multivariate analysis  Not associated: sex, age (ref. < 1 year), cardiovascular disease, obesity, asthma, type.1 diabetes, CKD, immunosuppression  *Adjusted for sex, age group, date of symptoms onset, days from symptoms onset to healthcare-seeking, pneumonia diagnosis at hospital admission, cardiovascular disease* | Age  Cardiovascular conditions  Pulmonary conditions  Immunocompromise  Chronic kidney disease  Diabetes  Obesity |
| **Nguyen 2022 [44]** | Single-center retrospective cohort study conducted in Vietnam (Jul/21 - Dec/21) | < 17 years  n = 555 | ICU  (severe/critical) | Age (median [IQR] months): severe/critical 38.7 (5/9 - 113.9) vs. mild/moderate 21 (2.9 - 80.7), p 0.03  Underlying disease: 60.4% (n = 58/96 severe/critical, p < 0.001) vs. 29.2%  Neurologic disease: 15.6% (n = 15/96 severe/critical, p 0.002) vs. 6.3%  Congenital heart disease: 7.3% (n = 7/96 severe/critical, p 0.04) vs. 2.4%  GI/hepatic disease: 8.3% (n = 8/96 severe/critical, p 0.004) vs. 2.4%  Obesity: 20.8% (n = 20/96 severe/critical, p < 0.001) vs. 3.1%  Not associated: sex, respiratory disease, kidney disease, hematologic disease  Multivariate analysis  Underlying disease: OR 3.1 (1.37 - 6.99, p 0.006)  Not associated: age  *Adjusted for age, underlying disease, symptoms, and labs* | Sex  Cardiovascular conditions  Neurologic conditions  Pulmonary conditions  GI and liver conditions  Chronic kidney disease  Obesity |
| **Oliveira 2021 [45]** | Multi-center retrospective cohort study conducted in Brazil (Feb/20 - Jan/21) | < 20 years  n = 11,613 | Death | Pulmonary: HR 1.8 (1.27 - 2.55, p 0.001)  Neurology: HR 2.66 (2.2 - 3.21, p < 0.001)  Oncology: HR 3.22 (2.68 - 3.87, p < 0.001)  Cardiology: HR 3.36 (2.68 - 4.2, p < 0.001)  Hematology: HR 1.66 (1.17 - 2.35, p 0.004)  Renal: HR 3.4 (2.48 - 4.66, p < 0.001)  Diabetes: HR 2.16 (1.53 - 3.06, p < 0.001)  Obesity: HR 1.93 (1.29 - 2.9, p < 0.001)  Syndrome: HR 3.11 (2.33 - 4.15, p < 0.001)  Protector  Asthma: HR 0.60 (0.42 - 0.85, p < 0.001)  Not associated: sex  Multivariate analysis  < 2 years (ref. 2 - 11 years): aHR 2.36 (1.94 - 2.88, p < 0.0001)  12 - 19 years (ref. 2 - 11 years): aHR 2.23 (1.84 - 2.71, p < 0.0001)  1 comorbidity: aHR 1.96 (2.52 - 3.47, p < 0.0001)  2 comorbidities: aHR 4.96 (3.8 - 6.48, p < 0.0001)  ≥ 3 comorbidities: aHR 7.28 (4.56 - 11.6, p < 0.001)  *Adjusted for age, macroregion, ethnicity, number of comorbidities* | Age  Sex  Medical complexity  Cardiovascular conditions  Neurologic conditions  Pulmonary conditions  Chronic kidney disease  Diabetes  Obesity |
| **Oliveira 2022 [46]** | Multi-center retrospective cohort study conducted in Brazil (Feb/20 - Jan/21) | < 20 years  n = 21,591 | ICU, mechanical ventilation, or disease-related death | Diabetic vs. Non-diabetic  Death – Multivariate analysis  Diabetes aHR 1.53 (1.17 - 2.03, p 0.002)  Kidney disorders: aHR 3.39 (1.42 – 8.09, p 0.006)  Obesity aHR 3.77 (1.83 – 7.76, p < 0.0001)  Not associated: age, sex, cardiovascular disorders, amount of comorbidities  *Adjusted for age, gender, region, ethnicity, respiratory symptoms, cardiovascular diseases, kidney disorders and obesity* | Chronic kidney disease  Diabetes  Obesity |
| **Oliveira 2023-1 [47]** | Multi-center retrospective cohort study conducted in Brazil (Feb/20 - Jan/21) | < 20 years  n = 21,591 | Death | Hematologic (malignant, non-malignant, non-specified) vs. Non-hematologic disease  Hematological disease: HR 2.4 (1.98 - 2.91)  Multivariate analysis  > 10 years of age (ref. 0 - 10 years): aHR 2.19 (1.46 – 3.19)  Male: aHR 1.52 (1.02 – 2.27)  Malignant hematological disorder: aHR 2.22 (1.47 – 3.36)  Abdominal pain at admission: aHR 2.75 (1.76 - 4.27)  Not associated: non-malignant hematological disorders, non-specified hematological disorders  *Adjusted by competing risk* | Immunocompromise  Sickle cell disease |
| **Oliveira 2023-2 [48]** | Multi-center retrospective cohort study conducted in Brazil (Feb/20 - Mar/22) | 2 - 19 years  n = 30,405 | Death | Asthmatics (without other comorbidities, with other comorbidities) vs. Non-asthma (without other comorbidities, with other comorbidities)  Multivariate analysis  Ref. No asthma + No other comorbidities  Asthma + Other comorbidities: aHR 2 (1.51 - 2.65, p < 0.001)  No asthma + Other comorbidities: aHR 3.17 (2.89 - 3.49, p < 0.001)  Age 12 - 20 years (ref. < 12 years): aHR 1.86 (1.68 - 2.05, p < 0.001)  Protector  Asthma + No other comorbidity: aHR 0.39 (0.29 - 0.53, p < 0.001)  Not associated: sex  *Adjusted for comorbidity group, sex, age, ethnicity, wave, region of admission, and signs and symptoms at admission (fever, dyspnea, respiratory distress, gastrointestinal, and oxygen saturation)* | Pulmonary conditions |
| **Parri 2020 [49]** | Multi-center retrospective cohort study conducted in Italy (Mar/20) | < 19 years  n = 130 | ICU, IMV, multiorgan failure or shock, encephalopathy, myocardial injury or heat failure, coagulation dysfunction, or acute kidney injury | < 6 months (compared to older children): 6/35 (17.1%) vs 3/86 (3.5%) p 0.034 OR 5.6 (1.3 to 29.1) | Sex |
| **Qian 2021 [50]** | Multi-center retrospective cohort study conducted in China (Jan/20 - Mar/20) | 1 month - 16 years  n = 127 | Severe/Critical | Do not report individual OR. Combined with estimates of Harwood et al. individual patient meta-analysis. | Data extracted from Harwood et al. individual patients’ data.  Cardiovascular conditions  Neurologic conditions  Immunocompromise |
| **Richardson 2020 [51]** | Multi-center cohort study conducted in USA (Mar/20 - Apr/20) | < 19 years + adults  n = 110 | Death | Do not report individual OR. Combined with estimates of Harwood et al. individual patient meta-analysis. | Data extracted from Harwood et al. individual patients’ data.  Age  Sex  Cardiovascular conditions  Pulmonary conditions |
| **Rivas-Ruiz 2020 [52]** | Multi-center retrospective cohort study conducted in Mexico (Mar/20 - May/20) | < 18  n = 1,443 | Death | Multivariate analysis  Not associated: age, immunosuppression, high cardiovascular risk, sex, obesity  *Adjusted for age, pneumonia, immunosuppression, high cardiovascular risk, and intubation* | Pulmonary conditions  Immunocompromise  Chronic kidney disease  Diabetes  Obesity |
| **Saatci 2021 [53]** | Multi-center retrospective cohort study conducted in UK (Jan/20 - Nov/20) | < 19 years  n = 343 | ICU | Multiple imputation analysis  T1 DM: aOR 10.35 (1.23 - 87.3)  Congenital heart disease: aOR 9.47 (3.79 - 26.54)  Cerebral palsy: aOR 25.16 (4.53 - 139.83)  Epilepsy: aOR 6.81 (1.27 - 36.46)  Sickle cell disease: aOR 18.31 (2.22-150.95)  Not associated: age, sex, asthma  *Adjusted for age, sex, deprivation level, household size and comorbidities* | Age  Sex  Cardiovascular condition  Neurologic conditions |
| **Sanchez-Piedra 2022 [54]** | Multi-center retrospective cohort study conducted in Mexico (Mar/20 - Jun/21) | < 20 years  n = 131,001 | Death | Multivariate analysis  Diabetes: aOR 3.898 (2.596−5.851, p 0.00)  Immunosuppression: aOR 5.410 (4.088−7.158, p 0.00)  Obesity: aOR 1.876 (1.397−2.521, p 0.00)  Hypertension: aOR 1.906 (1.239−2.932, p 0.003)  Cardiovascular disease: aOR 2.288 (1.482−3.531, p 0.00)  CKD: aOR 13.250 (9.066−19.350, p 0.00)  Not associated: sex  *Adjusted for sex, age, and comorbidities* | Age  Medical complexity  Cardiovascular conditions  Pulmonary conditions  Immunocompromise  Chronic kidney disease  Diabetes  Obesity |
| **Sedighi 2022 [55]** | Multi-center retrospective cohort study conducted in Iran (Mar/20 - May/20) | < 16 years  n = 166 | ICU | I**CU**  Age < 1 year: 59% (n = 17), p 0.048  Age 1 - 5 years: 35% (n = 19)  Age 5 - 10 years: 35% (n = 9)  Age 10 - 15 years: 28% (n = 16)  Any comorbidity: 57% (n = 39) vs. 23% (n = 22), p < 0.001  **Death**  Any comorbidity 32% (n = 22) vs. 8.2% (n = 8), p < 0.001  Not associated: age | Age |
| **Sena 2021 [56]** | Multi-center retrospective cross-sectional study conducted in Brazil (Feb/20 - Jun/20) | < 20 years  n = 682 | Death | Multivariate analysis  Age < 30 days (ref. 10 - 19 years): aOR 5.17 (1.81–14.77, p 0.002)  Age 1 - 11 months (ref. 10 - 19 years): aOR 3.28 (1.21–8.91, p 0.020)  Comorbidities: aOR 14.13 (6.35–31.44, p < 0.001)  Not associated: sex, age 1 - 9 years  *Adjusted for age and comorbidities* | Age  Sex |
| **Simoes E Silva 2022 [57]** | Multi-center retrospective cohort study conducted in Brazil (Feb/20 - May/21) | < 20 years  n = 21,591 | ICU | Obesity vs. Non-obesity  Multivariate analysis  Obesity: aHR 1.42 (1.11–1.79, p 0.004)  *Adjusted for age, gender, region, ethnicity, respiratory symptoms, and comorbidities* | Obesity |
| **Sorg 2022 [58]** | Multi-center cross-sectional study conducted in Germany (Jun/20 - May/21) | < 18 years  n = 5,780 | Hospitalization for treatment, admission to ICU, death due to COVID-19, or PIMS-TS | **Hospitalization** (n = 1059): 7.13 (5.97 - 8.86) per 10,000 children  **ICU** (n = 328/1059): 2.21 (1.85 - 2.74) per 10,000 children  **Death** (n = 14): 0.09 per 10,000 children  **ICU**  Age < 5 years (n = 60/391): 1.4 (1.17 - 1.74) per 10,000 children (p < 0.0001)  Age 5 - 11 years (n = 88/197): 1.55 (1.29 - 1.92) per 10,000 children (p < 0.0001)  Age 12 - 17 years (n = 180/471): 3.7 (3.1 - 4.59) per 10,000 children (p < 0.0001)  Comorbidity: p < 0.0001  Not associated: no comorbidity  **Death**  Age < 5 years (n = 8): 0.19 (0.16 - 0.23) per 10,000 children  Age 5 - 11 years (n = 4): 0.07 (0.06 - 0.09) per 10,000 children  Age 12 - 17 years (n = 2): 0.04 (0.03 - 0.05) per 10,000 children | Age |
| **Sousa 2021 [59]** | Multi-center retrospective cross-sectional study conducted in Brazil (Jan/20 - Dec/20) | < 20 years  n = 5,857 | Death | Multivariate analysis  1 comorbidity: aOR 3.71 (2.89 - 4.76)  ≥ 2 comorbidities: aOR 9.67 (6 .89 - 13.57)  Cardiovascular disease: aOR 4.98 (3.31 - 7.49)  Renal disease: aOR 4.79 (2.67 - 8.6)  Obesity: aOR 2.53 (1.26 - 5.09)  DM: aOR 3.31 (1.97 - 5.55)  Immunosuppression: aOR 3.41 (2.39 - 4.86)  Neurologic disease: aOR 2.69 (1.83 - 3.96)  Hematologic disease: aOR 2.35 (1.37 - 4.05)  Lung disease: OR 2 (1.1 - 3.63)  Protector  Asthma: OR 0.42 (0.24 - 0.67)  Not associated: hepatic disease  *Adjusted for municipalities and hospitals* | Age  Medical complexity  Cardiovascular conditions  Neurologic conditions  Pulmonary conditions  GI and liver conditions  Diabetes  Obesity |
| **Storch-de-Gracia 2020 [60]** | Single-center retrospective cohort study conducted in Spain (Mar/20 - Apr/20) | < 18 years  n = 39 | **Complicated**: need for hemodynamic support for stabilization (infusion of fluids or vasopressors) or respiratory support (high flow nasal cannula, non-invasive ventilation, invasive ventilation) or encephalopathy. | **ICU**: 35.9% (n = 14)  - 14/15 complicated cases were admitted to ICU  Age < 1 year: 13.3% (n = 2)  Age 1 - < 6 years: 6.6% (n = 1)  Age 6 - 10 years: 40% (n = 6)  Age > 10 years: 40% (n = 6)  Sex (male): 60% (n = 9)  Not associated: comorbidities | Age  Sex |
| **Swann 2020 [61]** | Multi-center cohort study conducted in England, Wales, and Scotland (Jan/20 - Jul/20) | < 19 years  n = 580 | ICU | **ICU** (Excluding MIS-C)  Age < 1 month: 21.8% (n = 17/78 ICU adm) vs. 6.8% (n = 34/502 ward adm), p < 0.001  Any comorbidity: 67.9% (n = 53/78 ICU adm) vs. 40.8% (n = 205/502 ward adm), p < 0.001  Prematurity: 19.2% (n = 15/78 ICU adm) vs. 6% (n = 30/502 ward adm), p 0.001  Respiratory comorbidities: 12.8% (n = 10/78 ICU adm) vs. 4% (n = 20/502 ward adm), p 0.004  Cardiac comorbidities: 15.4% (n = 12/78 ICU adm) vs. 4.8% (n = 24/502) ward adm), p 0.002  Neurological comorbidities: 21.8% (n = 17/78 ICU adm) vs. 9.2% (n = 46/502 ward adm), p 0.003  Neurodisability: 11.5% (n = 9/78 ICU adm) vs. 7.8% (n = 39/502 ward adm), p 0.014  Cardiac comorbidities: 15.4% (n = 12/78 ICU adm) vs. 4.8% (n = 24/502 ward adm), p 0.002  GI comorbidities: 9% (n = 7/78 ICU adm) vs. 2.2% (11/502 ward adm), p 0.007  Not associated: age > 1 month, sex, obesity, immunosuppressant use prior to presentation, asthma, hematology/oncology/immunology, malnutrition, genetic comorbidities, diabetes, renal comorbidities, metabolic comorbidities, other endocrine comorbidities, rheumatology comorbidities  Multivariate analysis (COVID-19 + MIS-C)  Age < 1 month (ref. 15 - 19 years): aOR 3.21 (1.36 - 7.66, p 0.008)  Age 10 - 14 years (ref. 15 - 19 years): aOR 3.23 (1.55 - 6.99, p 0.002)  Not associated: sex, age 1 month - 1 year, age 1 - 4 years, age 5 - 9 years, comorbidities  *Adjusted for sex, age group, ethnicity, and any comorbidity* | Age  Sex  Prematurity  Cardiovascular conditions  Neurologic conditions  Pulmonary conditions  Immunocompromise  GI and liver conditions  Chronic kidney disease  Diabetes  Obesity |
| **Ungar 2023 [62]** | Multi-center retrospective cohort study conducted in USA (Mar/20 - Sep/21) | < 21 years  n = 475 | ICU | Multivariate analysis  Protector  Age < 1 year (ref. 6 - 11 years): aOR 0.13 (0.04 - 0.38, p < 0.001)  Not associated: sex, age 1 - 5 years, age 12 - 17 years, age 18 - 20 years  *Adjusted for age, and hospital (NYULH/HMH)*  Multivariate analysis  At least 1 comorbidity: OR 4.65 (2.95 - 7.33, p < 0.001) - not included in multivariate analysis.  Malignancy: aOR 4.25 (1.55 - 11.62, p 0.005)  Congenital heart disease: OR 3.04 (1.05 - 8.86, p 0.03) - not included in multivariate analysis.  Hypertension: OR 6.1 (1.5 - 24.79, p 0.004) - not included in multivariate analysis  Intellectual disability: aOR 5.45 (2.36 - 12.61, p < 0.001)  Prematurity: aOR 4.95 (1.7 - 14.43, p 0.003)  Tracheostomy: OR 17.59 (3.84 - 80.53, p < 0.001) - not included in multivariate analysis.  Not associated: asthma, obesity, cystic fibrosis, CKD, diabetes mellitus, down syndrome, rheumatologic disease, sickle cell disease, bone marrow transplant, solid organ transplant, neurologic disorder  *Adjusted for asthma, obesity, malignancy, intellectual disability, prematurity, and neurologic disorder.*  < 1 year - Univariate analysis  Not associated: at least 1 comorbidity, malignancy, congenital heart disease, CKD, prematurity, bone marrow transplant, neurologic disorder | Age  Sex  Prematurity  Cardiovascular conditions  Neurologic conditions  Pulmonary conditions  Immunocompromise  Sickle Cell Disease  Chronic kidney disease  Diabetes  Obesity |
| **van der Zalm 2021 [63]** | Single-center retrospective cohort study conducted in South Africa (Apr/20 - Jul/20) | < 14 years  n = 62 | ICU | **ICU**  Age 0 - 3 months: 18.8% (n = 3/16)  Age > 3 - 12 months: 28.6% (n = 2/7)  Age > 1 - 5 years: 16.7% (n = 3/18)  Age > 5 years: 30% (n = 3/10)  **IV**  Age 0 - 3 months: 33.3% (n = 1/3 ICU adm)  Age > 1 - 5 years: 66.7% (n = 2/3 ICU adm)  Age > 5 years: 33.3% (n = 1/3 ICU adm)  **CVS**  Age 0 - 3 months: 66.7% (n = 2/3 ICU adm)  Age > 5 years: 66.7% (n = 2/3 ICU adm)  **Death**  Age 0 - 3 months: 6.3% (n = 1/16) | Data extracted from Harwood et al. individual patients’ data.  Age |
| **Vasconcelos 2023 [64]** | Multi-center retrospective cohort study conducted in Brazil (Feb/20 - May/21) | < 20 years  n = 21,591 | ICU, IMV, and death | Kidney disease (n = 290) vs. Non-kidney disease (n = 21,301) cohort  **ICU**: KD 45.1% (n = 120) vs. non-KD 24.1% (n = 5,123), p < 0.0001  **IMV**: KD 23.4% (n = 63) vs. non-KD 9.7% (n = 2067), p < 0.001  **Death**: KD 20.3% (n = 59) vs. non-KD 7.5% (n = 1602), p < 0.001  Death  Kidney disease: HR 2.85 (2.21 - 3.68, p < 0.0001)  Death for the first 10, 20, 30 and 50 days of hospitalization:  - KD: 11.2%, 15.9%, 18.9% and 21.3%  - Non-KD: 4.5%, 6.3%, 7.3%, and 7.6%  Nephrotic syndrome: 28% (n = 7/25, p 0.017)  Kidney transplant recipients: 12% (n = 3/25, p 0.017)  Congenital anomalies of kidney and urinary tract: 0% (n = 0/12, p 0.017)  CKD: 30.9% (n = 25/81, p 0.017)  Non-specified KD: 16.3% (n = 24/147, p 0.17)  Invasive ventilation  Nephrotic syndrome: 22.7% (n = 5/25, p 0.01)  Kidney transplant recipients: 12.5% (n = 3/25, p 0.001)  Congenital anomalies of kidney and urinary tract: 8.3% (n = 1/12, p 0.001)  CKD: 39% (n = 30/81, p 0.001)  Non-specified KD: 17.1% (n = 24/147, p 0.01) | Chronic kidney disease |
| **Verma 2021 [65]** | Multi-center retrospective cohort study conducted in USA (Mar/20) | < 22 years  n = 82 | ICU | Any comorbidity: 70% (n = 16/23 ICU adm) vs. 37% (n = 22.59 ward adm), p 0.08  Obesity: 63% (n = 10/23 ICU adm) vs. 28% (n = 28/59 ward adm), p 0.02  Not associated: asthma, chronic lung disease, congenital heart disease, diabetes mellitus (type1 or 2), malignancy and/or immunosuppression | Sex  Cardiovascular conditions  Pulmonary conditions  Immunocompromise  Diabetes  Obesity |
| **Wanga 2021 [66]** | Multi-center retrospective cross-sectional study conducted in USA (Jul/21 - Aug/21) | < 18 years  n = 713 | ICU, IMV, CVS, or death | ICU  Age < 1 year: 19.35% (n = 34/176), p < 0.001  Age 1 - 4 years: 25.4% (n = 31/122), p < 0.001  Age 5 - 11 years: 25.9% (n = 37/143), p < 0.001  Age 12 - 17 years: 39.7% (n = 108/272), p < 0.001  Underlying condition: 34.7% (n = 167/481) vs. 18.5% (n = 43/232), p < 0.001  Obesity: 41.1% (n = 95/231) vs. 23.9% (n = 115/482), p < 0.001  Not associated: viral coinfection  Invasive ventilation  Age < 1 year: 14.6% (n = 12/176), p < 0.001  Age 1 - 4 years: 10.8% (n = 7/122), p < 0.001  Age 5 - 11 years: 18% (n = 11/143), p < 0.001  Age 12 - 17 years: 14.7% (n = 26/272), p < 0.001 | Obesity |
| **Ward 2022 [67]** | Multi-center retrospective cohort study conducted in UK (Feb/20 - Jan/21) | < 18 years  n = 6,338 | ICU | Multivariate analysis  Any comorbidity: aOR 6.75 (5.64 - 7.86)  Non life limiting comorbidity: aOR 3.02 (2.07 - 3.97)  Life limiting comorbidity: aOR 14.36 (11.92 -16.8)  One body system comorbidity: aOR 2.03 (1.06 - 3.0)  More than one body system: aOR 10.91 (9.06 - 12.76)  Cancer/hem comorbidity: aOR 9.7 (6.9 - 12.5)  Cardiovascular comorbidity: aOR 15.43 (12.64 - 18.22)  Neurological comorbidity: aOR 12.31 (9.78 - 14.84)  Respiratory comorbidity: aOR 11.16 (8.34 - 13.98)  Neuro + Cardio comorbidity: aOR 19.34 (14.84 - 23.83)  Neuro + Resp comorbidity: aOR 18.26 (13.32 - 23.2)  Resp + Cardio comorbidity: aOR 20.65 (15.47 - 25.84)  Asthma: aOR 4.5 (1.67 - 7.33)  Diabetes mellitus: aOR 10.96 (2.69 - 19.22)  Epilepsy: aOR 13.67 (8.77 - 18.58)  Trisomy 21: aOR 18.61 (8.26 - 28.96)  *Adjusted for age, sex, IMD category, and ethnicity* | Age  Sex  Medical complexity  Cardiovascular conditions  Neurologic conditions  Pulmonary conditions  Immunocompromise  Sickle Cell Disease  GI and liver conditions  Chronic kidney disease  Diabetes |
| **Wong-Chew 2022 [68]** | Multi-center retrospective cohort study conducted in Mexico (Feb/20 - Sep/20) | < 18 years  n = 18,465 | Death | Multivariate analysis - Mexico  Diabetes: HR 8.0 (4.6 - 13.9, p < 0.0001) - not included in multivariate analysis  Hypertension: aHR 3.4 (1.8 - 6.6, p < 0.0001)  Obesity: aHR 1.7 (1.1 - 2.8, p 0.02)  CKD: HR 6.8 (3.4 - 13.8, p < 0.0001) - not included in multivariate analysis.  Not associated: sex, chronic lung disease, immunosuppression  *Adjusted for age, pneumonia, comorbidities, intubation, sick-contact, and origin.*  Univariate analysis - Ciudad de Mexico  Sex (male): HR 1.9 (1.03 - 3.9, p 0.04)  Diabetes: HR 15.6 (4.8 - 50.6, p < 0.0001)  Chronic lung disease: HR 43 (5.9 - 314.2, p < 0.0001)  Immunosuppression: HR 8.1 (3.4 - 19.4, p < 0.0001)  Hypertension: HR 17.1 (5.3 - 55.5, p < 0.0001)  Obesity: HR 4.0 (1.7 - 9.6, p 0.002)  CKD: HR 22.8 (8.1 - 64.1, p < 0.0001)  Protector  Influenza vaccine in the last year: HR 0.2 (0.05 - 0.81, p 0.02) | Age  Sex |
| **Woodruff 2022 [69]** | Multi-center retrospective cohort study conducted in USA (Mar/20 - May/21) | < 18 years  n = 2,095 | ICU, IMV, or in-hospital death | Multivariate analysis  ≥ 1 underlying medical condition: aRR 1.9 (1.5 - 2.6, p < 0.0001)  Not associated: age, sex  *Adjusted for age, sex, ethnicity, residential type, and underlying medical conditions.*  Age < 2 years - Multivariate analysis  Chronic lung disease (excl asthma): aRR 2.2 (1.1 - 4.3, p 0.02)  Neurologic disorder: aRR 2.1 (1.5 - 2.9, p < 0.0001)  Cardiovascular disease: aRR 1.6 (1.1 - 2.3, p 0.007)  Airway abnormality: aRR 1.6 (1.1 - 2.3, p 0.02)  Prematurity: aRR 1.6 (1.3 - 2.1, p < 0.001)  Protector  Feeding tube dependent: aRR 0.4 (0.2 - 0.8, p 0.02)  Age 2 - 17 years - Multivariate analysis  Diabetes (type 1 or 2): aRR 2.4 (2.0 - 2.8, p < 0.0001)  Feeding tube dependent: aRR 2.2 (1.7 - 2.9, p < 0.0001)  Chronic lung disease (excl asthma): aRR 1.2 (0.9 - 1.6, p 0.004)  Obesity: aRR 1.3 (1.1 - 1.5, p 0.002)  Developmental delay: aRR 1.3 (1.1 - 1.6, p 0.004)  Protector  Blood disorder: aRR 0.6 (0.4 - 0.8, p 0.002)  Not associated: chronic metabolic disease, asthma, airway abnormality, immunocompromised condition, cardiovascular disease, neurologic disorder  *Adjusted for age group, sex, race and ethnicity group, and housing type* | Age  Sex  Prematurity  Cardiovascular conditions  Neurologic conditions  Pulmonary conditions  Immunocompromise  GI and liver conditions  Diabetes  Obesity |
| **Yayla 2021 [70]** | Single-center retrospective cohort study conducted in Turkey (Mar/20 - May/20) | < 18 years  n = 77 | Severe/Critical | 1/77 patient was diagnosed with fulminant myocarditis, was admitted to the ICU, required invasive ventilation, was transferred for ECMO, and died. | Data extracted from Harwood et al. individual patients’ data.  Neurologic conditions |

#### **Supplementary Table 2**. Characteristics of studies included in the narrative review.

| **Study**  (Author year) | **Study design and setting** | **Subjects** | **Severity definition** | **Results: Risk factor**  (ES [95%CI, p – value]) | **Risk factors reviewed** |
| --- | --- | --- | --- | --- | --- |
| **Antoon 2021 [71]** | Multi-center retrospective cohort study conducted in USA (Apr/20 - Sep/20) | < 18 years  n = 19,976 | ICU ± mechanical ventilation, shock, or death | Mixed COVID-19 + MIS-C data  **ICU** = 461  **Composite** = 380  Multivariate analysis  Black race/ethnicity: aOR 1.52 (1.20 – 1.93)  Other race/ethnicity: aOR 1.62 (1.24 – 2.12)  Age 5 – 11 years: aOR 2.66 (2.14 – 3.32)  Age 12 – 17 years: aOR 2.09 (1.71 – 2.55)  Cardiovascular conditions: aOR 3.10 (2.56 – 2.74)  Neuromuscular conditions: aOR 1.56 (1.23 – 1.96)  Obesity/Type2 diabetes: aOR 2.16 (1.64 – 2.83)  Pulmonary conditions: aOR 2.66 (1.83 – 3.86)  Not associated: Hispanic ethnicity, sex, payor type, asthma, immunocompromise  *Adjusted for age, sex, race and ethnicity, payor type, and complex chronic conditions* | Sex  Cardiovascular conditions  Pulmonary conditions |
| **Armann 2021 [72]** | Multi-center prospective cohort study conducted in Germany and Austria (Mar/20 - Apr/21) | < 18 years  n = 1,501 | ICU | Mixed COVID-19 + MIS-C data  **ICU** = 111  **IV** = 57  **CVS** = 4  **Death** = 8  Bivariate analysis  Cyanotic heart disease: RR 6.20 (2.92 – 13.16)  Pulmonary hypertension: RR 7.82 (4.24 – 14.43)  Epilepsy: RR 2.33 (1.09 – 4.94)  Psychomotor retardation: RR 4.80 (2.90 – 7.93)  Fatty liver disease: RR 7.98 (4.09 – 15.55)  S/p cardiac surgery: RR 7.82 (4.24 – 14.43)  Multivariate analysis  Age 1 – 5 years: aRR 2.27 (1.13 – 4.58)  Age 6 – 11 years: aRR 7.33 (3.92 – 13.70)  Primary immunodeficiency: aRR 2.68 (1.15 – 6.24)  Trisomy 21: aRR 4.24 (1.42 – 12.64)  Coinfection: aRR 4.16 (2.03 – 8.50)  Not associated: age ≥ 12 years, sex, respiratory disease, cardiovascular disease, gastrointestinal disease, liver disease, renal disease, neurologic/neuromuscular disease, hematologic disease, oncological disease, autoimmune disease, tracheostomy, immunosuppressive therapy, home oxygen therapy, history of prematurity, asthma/wheezing, acyanotic heart disease, hypertension, diabetes, acute leukemia, hemolytic anemia  *Adjusted for all considered risk factors and ethnic background (Caucasian, Arabic, African, other)* | Sex  Cardiovascular conditions  Neurologic conditions  Pulmonary conditions  Immunocompromise  GI and liver conditions |
| **Catalan 2022 [73]** | Multi-center retrospective cohort study conducted in Spain (Mar/20 - Mar/21) | < 20 years + adults  n - 27,913 | Death | Mixed adult + pediatric data  **Death** = 1  Cardiovascular disease: OR 4.4 (3.93 – 5.08, p < 0.0001)  Pulmonary disease: OR 1.44 (1.26 – 1.65, p < 0.0001)  Neurology disorder: OR 8.05 (6.79 – 9.55, p < 0.0001)  Multivariate analysis  Sex (Male): aOR 2.04 (1.88 – 2.20, p < 0.0001)  Age (years): aOR 1.12 (1.12 – 1.13, p < 0.0001)  Metabolic and endocrine disease: aOR 1.34 (1.14 – 1.56, p 0.0003)  Renal disease: aOR 1.99 (1.76 – 2.24, p < 0.0001)  Hepatic disease: aOR 1.54 (1.76 – 2.24, p 0.0271)  Obesity: aOR 1.72 (1.22 – 2.43, p 0.0022)  Cancer: aOR 2.33 (2.06 – 2.63, p < 0.001)  Substance abuse: aOR 1.88 (1.33 – 3.14, p 0.0152)  Psychosis: aOR 1.45 (1.09 – 1.94, p 00114)  Protector  Anxiety disorder: OR 0.50 (0.37 – 0.68), p < 0.001)  Affective disorders: aOR 0.80 (0.61 – 0.99, p 0.0407)  Not associated: personality disorders, eating disorders  *Adjusted for age, sex, physical comorbidities, and psychiatric disorders* | Sex |
| **Choi 2022 [74]** | Systematic review and meta-analysis (Jan/20 - Aug/21) | ≤ 21 years  n = 17 studies | ICU, IMV or death | Neonates (2 studies): RR 2.69 (1.83 – 3.97)  Prematurity (2 studies): RR 2.0 (1.63 – 2.46)  Obesity (8 studies): RR 1.43 (1.24 – 1.64)  Diabetes (4 studies): RR 2.26 (1.95 – 2.62)  Chronic lung disease (4 studies): RR 2.62 (1.71 – 4.0)  Cardiac disease (5 studies): RR 1.79 (1.56 – 2.07)  Neurologic disease (5 studies): RR 2.21 (1.56 – 3.14)  Seizure disorder (2 studies): RR 1.73 (1.43 – 2.09)  Protector  Age < 3 months (2 studies): RR 0.26 (0.11 – 0.66)  Not associated: age < 1 year (2 studies), genetic disease (3 studies), asthma (7 studies), neurodevelopmental disorder (2 studies), immunocompromise/ immunosuppression/hemato-oncological disease (4 studies), hemato-oncological disease (3 studies), immunosuppression (2 studies) | Sex  Prematurity  Cardiovascular conditions  Neurologic conditions  Pulmonary conditions  Immunocompromise  Diabetes  Obesity |
| **Choi 2023 [75]** | Multi-center retrospective cohort study conducted in Korea (Aug/21 - Mar/22) | < 18 years  n = 612 | **Severe**: LRTI requiring oxygen via nasal prongs or a mask, high-flow nasal cannula at ≤ 2 L/kg/min, or end- organ damage  **Critical**: ICU admission, mechanical ventilation, or ECMO, sepsis/ septic shock, multiple organ failure, or death. | **ICU** = 21  **IMV** = 14  **Death** = 2  Severe/Critical  Age: 9.9 years (IQR, 3.3–13.0) vs. moderate, 2.2 years (IQR, 1.0–7.4) vs. asymptomatic/mild, 1.7 years (IQR 0.5–7.3) years; P < 0.001  Multivariate analysis - Moderate/Severe/Critical in Delta period  Neurologic disease: aOR 39.43 (6.9 – 268.3)  Obesity: aOR 8.18 (2.8 – 27.36)  Age 12 – 18 years: aOR 3.92 (1.46 – 10.85)  Not associated: complex chronic disease  Multivariate analysis - Moderate/Severe/Critical in Omicron period  Neurologic disease: aOR 9.8 (4.5 – 22.57)  Not associated: respiratory diseases, malignancy, obesity, complex chronic disease, age < 1 year | Age |
| **Forrest 2022 [76]** | Multi-center retrospective cohort study conducted in USA (Mar/20 - Dec/21) | < 18 years  n = 82,798 | Moderate/Severe  **Moderate**: Moderately severe COVID-19 related health conditions  **Severe**: Unstable COVID-19 health conditions | Mixed COVID-19 + MIS-C data  **ICU** = 813  **IMV** = 412  **CVS** = 205  **Death** = 27  **Composite** = 1,766  Multivariate analysis  Age < 1 year: aOR 4.5 (4.1 – 4.9)  Age 1 – 4 years: aOR 2.1 (1.9 – 2.3)  Age 12 – 17 years: aOR 1.6 (1.4 – 1.7)  Asthma with history of previous hospitalization: aOR 2.9 (2.6 – 3.3)  Cancer not actively treated: aOR 2.0 (1.4 – 2.8)  Cancer actively treated: aOR 11.8 (8.3 - 16.8  Cardiac disorder: aOR 2.7 (2.4 - 3)  Diabetes: aOR 6.3 (5-8.1)  Immunologic disorder: aOR 2.6 (2.2 - 3.2)  Mental health disorder: aOR 1.4 (1.2 - 1.5)  Obesity: aOR 1.6 (1.4 - 1.7)  Prematurity: aOR 1.3 (1.1 - 1.5)  Renal disorder: 1.8 (1.5 - 2.2)  Sickle cell disease: 2.9 (1.9 -4.5)  Not associated: sex, asthma with no previous hospitalization, trisomy 21  *Adjusted for demographic, institution, chronic conditions* | Age  Sex  Prematurity  Cardiovascular conditions  Immunocompromise  Sickle cell disease  Obesity |
| **Greenan-Barrett 2023 [77]** | Systematic review and meta-analysis (Jan/20 - Oct/21) | < 25 years  n = 30 studies | ICU, IMV or death | Immunosuppressed vs. Not-immunosuppressed  **ICU**: 12% vs. 2%  **IMV**: 8% vs. 1%  **Death**: 6.5% vs. 0.2% | Immunocompromise |
| **Harwood 2022 [78]** | Systematic review and meta-analysis (Jan/20 - May/21) | ≤ 21 years  n = 83 studies | ICU, IMV, CVS, or death | Age < 1 year: ICU OR 1.63 (1.40−1.90) / Death OR 2.08 (1.57−2.86)  Age > 14 years: Death OR 2.15 (1.61−2.88)  Any comorbidity: ICU OR 2.56 (1.77 – 3.71) / Death OR 4.16 (1.97 – 8.80)  Cardiovascular: ICU OR 2.88 (2.33 – 3.57) / Death OR 12.06 (2.37 – 61.34)  Respiratory: ICU OR 1.98 (1.23−3.19)  GI/Liver: ICU OR 3.08 (1.92−4.96) / Death OR 3.66 (1.71 – 7.84)  Neurological: ICU OR 2.60 (2.16 − 3.13) / Death OR 4.34 (2.0 – 9.43)  Malignancy: Death OR 10.76 (3.3 − 35.4)  Hematological: Death OR 2.92 (1.20 − 7.13)  Immunosuppression: Death OR 4.93 (1.57−15.5)  Trisomy 21: ICU OR 3.08 (1.02 − 9.32) / Death OR 5.12 (1.08 – 24.40)  CKD: ICU OR 2.63 (1.58 − 4.39) / Death OR 3.72 (2.44 – 5.67)  Endocrine including diabetes: ICU OR 3.06 (1.26 − 7.41) / Death OR 13.17 (2.57 – 67.45)  Metabolic including obesity: ICU OR 2.48 (1.66 − 3.70) / Death OR 11.65 (2.43 – 55.85)  Not associated: sex, 5 – 9 years, asthma | Age  Sex  Medical complexity  Cardiovascular conditions  Neurologic conditions  Pulmonary conditions  Immunocompromise  GI and liver conditions  Chronic kidney disease  Obesity |
| **Ho 2023 [79]** | Multi-center retrospective cohort study conducted in USA (Jan/20 - Jan/22) | < 19 years  n = 165,437 | ICU, IMV, or death | **ICU** = 2,954  **Death** = 31  **Composite** = 2,985  Multivariate analysis - 0 - 4 years  Sex (Female): aOR 0.82 (0.75 – 0.9)  Protector:  Age 1 year (compared with 3-year-old): aOR 0.58 (0.54 – 0.63)  Multivariate analysis - 5 - 11 years  Protector  Sex (Female): aOR 0.83 (0.74 – 0.94)  Age 10 years (compared with 6-year-old): aOR 0.78 (0.69 – 0.88)  At least 1 COVID-19 vaccine dose: aOR 0.48 (0.33 – 0.71)  Multivariate analysis - 12 - 18 years  Protector  At least 1 COVID-19 vaccine dose: aOR 0.59 (0.51 – 0.68)  *Adjusted for race, ethnicity, and insurance status* | Sex |
| **Jank 2023 [80]** | Multi-center cross-sectional study conducted in Germany (Jun/21 - Oct/21) | < 18 years  n = 2,169 | Need for hospitalization for COVID-19 related treatment, ICU, or death | **ICU** = 126  **Death** = 9  March 21 - June 21  Age < 5 years: 5.05 (3.11 - 6.99) per 10,000 children  Age 5 - 11 years: 5.71 (4.2 - 7.22) per 10,000 children  Age 12 - 17 years: 8.86 (6.91 - 10.82) per 10,000 children  July 21 - December 21  Age < 5 years: 7.33 (5.76 - 8.89) per 10,000 children  Age 5 - 11 years: 2.12 (1.71 - 2.54) per 10,000 children  Age 12 - 17 years: 3.97 (3.3 - 4.64) per 10,000 children  July 21 - December 21  Age < 5 years: 0.85 (0.62 - 1.08) per 10,000 children  Age 5 - 11 years: 0.31 (0.23 - 0.38) per 10,000 children  Age 12 - 17 years: 0.45 (0.34 - 0.56) per 10,000 children | Age |
| **Jone 2022 [81]** | Literature review | - | Mechanical ventilation or ECMO | "Because of the heterogeneity of congenital heart disease, however, it has been difficult to determine risk profiles for specific subtypes. In addition, the risk profile of pediatric versus adult patients with congenital heart disease is likely to be variable as a result of the diversity of risk factors such as age and associated morbidities, clouding our understanding of congenital heart disease–associated risk for severe COVID-19"  "More evidence is needed in children with solid-organ trans- plantation, but caution should be taken with this population given immunosuppression and the unknown risk of SARS-CoV-2 infection on the graft. The association of SARS-CoV-2 infection and future acute graft rejection in pediatric heart transplant recipients is currently unknown." | Cardiovascular conditions |
| **Leon-Abarca 2020 [82]** | Multi-center retrospective cohort study conducted in Mexico | < 18 years  n = 21,161 | ICU or death | Not associated: immunodeficiencies, asthma, obesity, cardiovascular disease, chronic kidney disease, hypertension, diabetes  *Adjusted for age and sex* | Immunocompromise  Obesity |
| **Lu 2021 [83]** | Single-center cross-sectional study conducted in China (Jan/20 - Feb/20) | < 16 years  n = 121 | **Critical** (group III): ARDS or respiratory failure, multiple organ dysfunction | **Death** = 1  **Composite** = 8  "Seven indices (T, Th, Tc, IL-6, IL-10, RBC, Hb) are helpful for early diagnosis of COVID-19 severity in children and may provide a laboratory basis for clinicians to offer timely treatment to children with critical COVID-19." | Immunocompromise |
| **Martin 2022 [84]** | Multi-center retrospective cohort study conducted in USA (Mar/20 - Sep/21) | < 19 years  n = 167,272 | IMV, CVS, or death | **COVID-19**: n = 8241  **MIS-C**: n = 707  **IMV** = 514  **CVS** = 426  **Death** = 95  **Composite** = 818  Multivariate analysis  Sex (male): aOR 1.32 (1.19 – 1.46, p < 0.001)  Obesity: aOR 1.19 (1.0 – 1.41, p 0.04)  Any complex chronic condition: aOR 1.16 (1.12 – 1.21, p < 0.001)  Cardiovascular condition: aOR 1.57 (1.27 – 1.92, p < 0.001)  Malignancy: aOR1.70 (1.18 – 2.45, p 0.005)  Protector:  Renal condition: aOR 0.59 (0.43 – 0.80, p 0.001)  Respiratory condition: aOR 1.46 (1.01 – 2.12, p 0.04)  Tech dependence: aOR 1.49 (1.08 – 2.04, p 0.01)  Not associated: asthma, congenital/genetic condition, gastrointestinal condition, heme/immune condition, metabolic condition, neonatal, neuromuscular condition, transplant  *Adjusted* for demographic and comorbidity variables | Sex  Cardiovascular conditions  Neurologic conditions  Pulmonary conditions  Obesity |
| **Navarro-Olivos 2021 [85]** | Multi-center retrospective cross-sectional study conducted in Mexico (Mar/20 - Dec/20) | < 18 years  n = 48,505 | Death | **Death** = 425  Multivariate analysis  Age 0 - 2 years: aOR 10.14 (8.06 - 12.74, p < 0.05)  Age 3 - 5 years: aOR 2.12 (1.51 - 2.98, p < 0.05)  Immunosuppression: aOR 1.03 (1.01 - 1.18, p 0.002)  Not associated: sex, age 6 - 11 years, diabetes, COPD, asthma, hypertension, cardiovascular disease, CKD, obesity, smoking  *Adjusted for sex and age, and comorbidities for the other comorbidities* | Age  Cardiovascular conditions  Neurologic conditions  Immunocompromise  Chronic kidney disease  Diabetes  Obesity |
| **Ouldali 2021 [86]** | Multi-center prospective cohort study conducted in France (Feb/20 - Jun/20) | < 18 years  n = 397 | Ventilatory support (HFNC, cPAP, bPAP, invasive ventilation), hemodynamic support, or death | **ICU** = 81  **IV** = 17  **CVS** = 21  **Death** = 6  Multivariate analysis  Age > 10 years: aOR 3.4 (1.1 - 10.3, p 0.34)  Not associated: sex, underlying comorbidities, asthma, other chronic respiratory diseases, immunosuppression or malignancy, diabetes, cardiac disease, obesity, neurologic disease  *Adjusted for age, comorbidities, hypoxemia, and procalcitonin.* | Age  Neurologic conditions  Immunocompromise |
| **Preston 2021 [87]** | Multi-center retrospective cohort study conducted in USA (Mar/20 - Oct/20) | < 19 years  n = 2,430 | ICU, IMV, or death | **ICU** = 747  **IMV** = 172  Presence of chronic condition: aOR 3.27 (2.44 - 4.37)  Sex (Male): aOR 1.52 (1.26 - 1.83)  Age 2 - 5 years vs 12 - 18 years: aOR 1.53 (1.11 - 2.13)  Age 6 - 11 years vs 12 - 18 years: aOR 1.53 (1.04 - 2.23)  Not associated: age 0 - 1 year, ethnicity, payment type  *Adjusted for within-facility correlation, age, sex, race/ethnicity, insurance type, and presence of chronic conditions* | Age |
| **Schober 2021 [88]** | Multi-center cross-sectional multinational study (Feb/20 - May/21) | < 18 years  n = 403 | Oxygen by non-invasive ventilation or HFNC or mechanical ventilation or vasopressors or death | **ICU** = 115  **Death** = 6  Multivariate analysis  Obesity: aOR 3.42 (1.76 - 6.66)  Pulmonary disease: aOR 1.73 (1.0 - 3.01)  Chromosomal disorder: aOR 4.47 (1.25 - 16.01)  1 comorbidity: aOR 1.9 (1.13 - 3.2)  ≥ 2 comorbidities: aOR 2.12 (1.19 - 3.79)  Not associated: non-asthma pulmonary disease, asthma, prematurity, immunosuppression, anemia/hemoglobinopathy, neurological disorder, cardiac disorder, metabolic disorder, renal disorder, hypertension, age 1 - 4 years, age 5 - 11 years, age ≥ 12 years  *Adjusted for age* | Neurologic conditions  Pulmonary conditions  Obesity |
| **Shi 2021 [89]** | Systematic review and meta-analysis (Jan/20 - Jul/21) | < 19 years  n = 56 studies | ICU, respiratory support, progression to severe or critical disease or death | Death  Age < 4 years (1 study): OR 4.02 (1.87 - 8.65, very low quality)  Age < 10 years (7 studies): OR 1.76 (1.07 - 2.9, I2 16%, very low quality)  Underlying conditions (5 studies): OR 8.68 (5.27 - 14.3, I2 0%, very low quality)  ICU  Age < 1 month (3 studies): OR 2.29 (1.48 - 3.56, I2 0%, moderate quality)  Underlying conditions (10 studies): OR 2.41 (1.77 - 3.27, I2 25.6%, low quality)  Congenital heart disease (4 studies): OR 2.9 (1.26 - 6.67, I2 0%, low quality)  Chronic pulmonary disease: OR 3.45 (1.47 - 8.07, I2 0%, low quality)  Sex (male): OR 1.2 (1.01 - 1.43, I2 0%, very low quality)  Obesity: OR 1.66 (1.1 - 2.5, I2 20.4%, very low quality) | Age  Sex  Cardiovascular conditions  Neurologic conditions  Pulmonary conditions  Diabetes  Obesity |
| **Solorzano-Santos 2022 [90]** | Multi-center retrospective cohort study conducted in Mexico (Apr/20 - Jan/22) | < 19 years  n = 261,099 | Death | **ICU** = 1,156  **Death** = 1,028  Inpatient + Outpatient  Diabetes: HR 2.0 (1.4 – 2.8)  Immunosuppression: HR 2.905 (2.311-3.652)  Hypertension: HR 2.134 (1.514-3.009)  Obesity: HR 1.92 (1.5 – 2.4)  Cardiovascular disease: HR 3.0 (2.323 - 3.878)  CKD: HR 3.479 (2.51 - 4.824)  Protector  Asthma: HR 0.627 (0.44-0.7)  Inpatient  Immunosuppression: HR 5.74 (4.51 - 7.3)  Obesity: HR 2.43 (1.9 - 3.1)  Cardiovascular disease: HR 4.96 (3.7 - 6.6)  CKD: HR 4.03 (2.7 - 5.9)  Protector  Diabetes: HR 0.33 (0.227 - 0.479)  Hypertension: HR 0.54 (0.36 - 0.79)  Asthma: HR 0.65 (0.54 - 0.78) | Age  Cardiovascular conditions  Pulmonary conditions  Immunocompromise  Chronic kidney disease  Diabetes  Obesity |
| **Tsankov 2021 [91]** | Systematic review and meta-analysis (Jan/20 - Oct/20) | < 21 years  n = 42 studies | **Severe**: Supplemental help to normal breathing and/or ICU | Severe  Comorbidities (37 studies): RR 1.79 (1.27 - 2.51, I2 94%)  Obesity (13 studies): RR 2.87 (1.16 - 7.07, I2 36%)  Not associated: cancer (5 studies)  Death  Comorbidities (15 studies): RR 2.81 (1.31 - 6.02, I2 82%) | Obesity |
| **Zachariah 2020 [92]** | Single-center retrospective cohort study conducted in USA (Mar/20 - Apr/20) | ≤ 21 years  n = 50 | Requirement of mechanical ventilation during hospitalization. | Obesity: 67% (n = 6/11 obese, p 0.03)  Protector  Age < 1 year: 0% (n = 0/14 < 1 year, p 0.05)  Not associated: sex, race/ethnicity, any comorbidity, asthma, immunosuppression, neurologic comorbidity, sickle cell disease, cardiac disease, diabetes, genetic syndromes, chronic respiratory disease | Sex  Cardiovascular conditions  Neurologic conditions  Pulmonary conditions  Immunocompromise  Sickle cell disease  Diabetes  Obesity |

| **Study**  (Author year) | **Study design**  **and setting** | **Subjects** | **Severity definition** | **Results: Risk factor**  (ES [95%CI, p-value]) | **Risk factor** |
| --- | --- | --- | --- | --- | --- |
| **Abu Shanap 2022 [93]** | Single-center observational study conducted in Jordan (Sep/20 - Nov/21) | < 18 years  n = 263 | ICU | **ICU**: 4.6% (n = 12)  **Death**: 1.9% (n = 5)  Multivariate analysis  Age ≥ 12 years: OR 3.4 (1.10 – 10.9, p 0.035)  ≥ 1 comorbidities: aOR 3.0 (1.2 – 8.3, p 0.025)  Obesity: aOR 5.0 (1.2 – 21.0, p 0.034)  Absolute lymphocyte count ≤ 300 cel/mm3: aOR 3.0 (1.2 – 1.4, p 0.022)  Absolute neutrophil count ≤ 500 cel/mm3: aOR 4.3 (1.4 – 14.0, p 0.014)  Not associated: sex, type of cancer, type of therapy, steroid in last 14 days, intense therapy, active therapy, time since last chemo/IS  *Adjusted for type of cancer, type of therapy, steroids in last 14 days, active/ongoing therapy, time since last chemo/IS, radiation, and ANC* | Immunocompromise |
| **Alqanathish 2023 [94]** | Single-center retrospective cohort study conducted in Saudi Arabia (Mar/20 - Mar/22) | < 19 years  n = 126 | Hospitalization | **Hospitalization**: 15% (n = 12)  **MIS-C**: 8.73% (n = 11)  **Death**: 2.38% (n = 3)  Age < 6 months: 57.9% (n = 11, p 0.000)  Age 6 - < 12 months: 15.8% (n = 3, p 0.000)  Age 12 - < 19 years: 26.3% (n = 5, p 0.000)  Sex (male): aOR 5.97 (1.44 - 24.71, p 0.014)  t-DMARDs: aOR 17.53 (1.85 - 165.8, p 0.012)  Glucocorticoids: aOR 6.69 (1.46 - 30.64, p 0.014)  Protector  Vaccinated: aOR 0.09 (0.02 - 0.54, p 0.008)  Not associated: c-DMARDs, other comorbidities | Rheumatic diseases |
| **Arlet 2020 [95]** | Multi-center case-series conducted in France (Mar/20 - Dec/21) | < 18 years + adults  n = 17 | ICU | **ICU**: 11.7% (n = 2) | Sickle Cell Disease |
| **Averbuch 2023 [96]** | Multi-center multinational prospective cohort study (Mar/20 - Ddec/21) | < 18 years  n = 89 | ICU or death within 2 months of diagnosis | **ICU**: 10.1% (n = 9)  **Death**: 7.8% (n = 7)  **Composite**: 12.3% (n = 11)  Non-malignant disease: HR 4.7 (1.25 – 17.76, p 0.02)  Chronic graft versus host disease (GVHD): HR 9.95 (2.37 – 41.73, p 0.002)  Use of janus kinase inhibitors: HR 3.42 (1.0 – 11.73, p 0.05)  Mycophenolic acid: HR 9.8 (2.94 – 32.62, p 0.0002)  Coinfection: HR 5.15 (1.50 – 17.62, p 0.009)  Not associated: use of corticosteroids | Hematopoietic cell transplantation (HCT) recipients |
| **Behr 2022 [97]** | Single-center retrospective cohort study conducted in USA (Mar/20 - Dec/20) | ≤ 20 years  n = 42 | Mechanical ventilation or death | Hematologic malignancies (n = 12)  Mechanical ventilation: 17% (n = 2)  Death: 17% (n = 2)  Composite: 33% (n = 4)  Not associated: solid tumors (n = 30) | Cancer |
| **Belsky 2021 [98]** | Systematic review and meta-analysis (Jan/20 - Jul/20) | < 18 years + adults  n = 100 studies | Pneumonia ARDS or ICU | **Composite**: pooled prevalence 20.24% (16.07 – 24.94)  Death  Immunocompromise: pooled prevalence 10.9% (6.9 – 16.2) | Cancer, hematopoietic cell, and solid organ transplant |
| **Bhatt 2022 [99]** | Multi-center retrospective multination cohort study (Mar/20 - May/21) | ≤ 21 years  n = 167 | Mechanical ventilation | **Mechanical ventilation**: 4%  **45-day survival in non-US recipients:** 85% (71 – 95)  **45-day survival in US recipients:** 98% (93 – 99) | Hematopoietic stem cell transplant recipients |
| **Brenner 2021 [100]** | Multi-center retrospective multinational cohort study (Mar/20 - Oct/20) | < 18 years  n = 209 | Mechanical ventilation | **Mechanical ventilation**: 1% (n = 2) | Inflammatory bowel disease |
| **Cardona-Hernandez 2021 [101]** | Multi-center multinational case-series (Jan/20 - Aug/20) | ≤ 23 years  n = 11 | ICU | **ICU due to COVID-19**: 0% (n = 0) | Type 1 diabetes |
| **Chivers 2023 [102]** | Multi-center cohort study conducted in UK (Mar/20 - Jun/21) | < 16 years + adults  n = 127 | Episode requiring hospital admission, including emergency department assessment and ‘hospice at home’ management | **IMV**: 8.6% (n = 11)  - 5/11 due to COVID-19  **CVS (inotropes):** 6.3% (n = 8)  - 2/8 due to COVID-19  **Death**: 3.9% (n = 5)  - 4/5 ≤ 12 months  - 1/5 due to COVID-19  - All children who died had significant CHD-related comorbidity which was stated as the primary cause of death on the death certificate, including sepsis, multiorgan failure and acute renal failure, and prolonged hospital admission before death (range 12–90 days) | CHD, arrhythmic disease, inherited cardiac disease and/or previous cardiac transplantation |
| **Demeterco-Berggren 2022 [103]** | Multi-center cross-sectional study conducted in USA (Apr/20 - Mar/21) | < 18 years  n = 415 | ICU | **ICU**: 8% (n = 35)  **Death**: 0% (n = 0) | Type 1 diabetes |
| **Dominguez-Rojas 2022 [104]** | Multi-center multinational prospective cohort study (Apr/21 - Nov/21) | < 14 years  n = 226 | Death | **IMV**: 2.7% (6)  **Death**: 13.2% (n = 30)  Multivariate analysis  Coinfection: aOR 3.09 (1.09 – 876)  New-onset oncologic diagnosis: aOR 11.91 (1.34 – 105.98)  Relapse of oncologic diagnosis: aOR 28.32 (3.23 – 248.09)  Not associated: underlying condition (not oncological), solid tumor (vs. hematological) | New or established oncologic diagnosis |
| **El‐Qushayri 2022 [105]** | Systematic review and meta-analysis (Jan/20 - Oct/21) | < 18 years  n = 21 studies | ICU or death | **ICU** (10 studies): pooled prevalence 14% (9 – 21%)  **ICU in HIC**: pooled prevalence 75% (1 – 32%)  **ICU in UMIC**: pooled prevalence 13% (7 – 23%)  **ICU in LMIC**: pooled prevalence 18% (6 – 41%)  **Death** (20 studies): pooled prevalence 9% (6 – 12)  **Death in HIC**: 3% (2 – 5%)  **Death in UMIC**: 12% (8 – 18%)  **Death in LMIC**: 13% (8 – 20%) | Cancer |
| **Emes 2021 [106]** | Multi-center multinational cohort study (Apr/20 - Oct/20) | < 18 years  n = 328 | ICU or death | **ICU**: 33.2% (n = 69)  **Death**: 0.5% (n = 1) | Trisomy 21 |
| **Fonseca 2021 [107]** | Single-center retrospective cohort study conducted in Colombia (Apr/20 - Sep/20) | < 18 years  n = 33 | ICU, IMV, CVS, or death | **ICU**: 21.2% (n = 7)  **IMV**: 9.0% (n = 3)  **CVS**: 9.0% (n = 3)  **Death**: 15.1% (n = 5) | Confirmed oncologic or benign hematologic diagnosis |
| **Freitas 2022 [108]** | Single-center cross-sectional study conducted in Brazil (Mar/20 - Oct/21) | < 18 years  n = 74 | **Severe**: increased need for supplemental oxygen support and/or ventilator support requirements  **Critical**: respiratory failure requiring mechanical ventilation with high parameters, acute respiratory distress syndrome and/or shock, or systemic inflammatory response | **Severe**: 22.2% (n =2)  **Critical**: 0% (n = 0)  **Intubation/Mechanical ventilation**: 11.1% (n = 1)  **Hemodynamic/Vasoactive support**: 11.1% (n = 1)  **Death**: 0% (n = 0)  Multivariate analysis  COVID-19 infection < 6 months after liver transplant: HR 0.01 (0.001 - 0.67, p 0.03)  Not associated: age, weight at liver transplant, other comorbidities | Pediatric liver transplant recipients (PLTR) |
| **Goss 2021 [109]** | Multi-center cohort study conducted in USA (Apr/20 - Jul/20) | < 19 years  n = 26 | ICU, IMV, CVS, or death | 0 patients had severity outcomes | Solid organ transplant recipients  - 2 lung transplant  - 6 heart transplant  - 8 kidney transplant  - 10 liver transplant |
| **Haeusler 2021 [110]** | Multi-center multinational cohort study (Jan/20 - Feb/21) | < 19 years  n = 131 | ICU or death | 60% (n = 67) had leukemia/lymphoma, 37% (n = 48) had solid tumor and 5 had primary immunodeficiency and HSCT  **ICU**: 11% (n = 15)  **Death** 3% (n = 4)  Multivariate analysis  Any comorbidity: aOR 2.94 (1.81 - 5.21)  Coinfection: aOR 1.74 (1.03 - 3.03)  Neutropenia: aOR 1.82 (1.13 - 3.09)  Not associated: age ≤ 7 years, sex (male), leukemia vs. solid tumor, intensive treatment, radiotherapy, relapse/progressive, HSCT, CAR-T, active cancer treatment, chronic lung disease, bacterial coinfection, lymphopenia  *Adjusted for any comorbidity, lung disease, coinfection, neutropenia and lymphopenia.* | Cancer or hematopoietic stem cell transplant |
| **Hammad 2021 [111]** | Single-center prospective cohort study conducted in Egypt (May/20 - Nov/20) | < 19 years  n = 76 | ICU, septic shock, or mechanical ventilation | **Composite**: 15.7% (n = 12)  Absolute lymphocyte count ≤ 500 cel/mm3: 100% (n = 12) vs. 0% (ALC > 500 cel/mm3)  Not associated: sex, age, cancer diagnosis, intensive chemotherapy, days from last chemotherapy, neutropenia | Cancer |
| **Hoogenboom 2021 [112]** | Multi-center cohort study conducted in USA (Jan/20 - Jan/21) | Pediatric + Adults  n = 115 | ICU, IMV, or in-hospital death | Not associated: sickle cell disease, sickle cell trait  *Adjusted for age, sex, race, ethnicity, and comorbidities* | Sickle Cell Disease and Sickle Cell Trait |
| **Hoogenboom 2022 [113]** | Systematic literature review (Jan/20 - Oct/21) | Pediatric + Adults  n = 17 studies | Oxygen, ICU, mechanical ventilation, or death | While the literature suggests that most children with SCD who become infected with SARS-CoV-2 have mild disease and low risk of death, children with SCD, especially those with SCD-related comorbidities, are more likely to be hospitalized and require escalated care than children without SCD of the same age.  There was not sufficient data to review children with SCT and COVID-19. | Sickle Cell Disease |
| **Kahraman 2023 [114]** | Single-center retrospective cohort study conducted in Turkey (Jan/20 - Jun/21) | < 18 years + adults  n = 131 pediatric | ARDS or death | **ICU**: 1.5% (n = 2)  **IMV**: 0.8% (n = 1)  **Death**: 0.8% (n = 1)  Disorders of complex molecule degradation: OR 9.7 (p < 0.01) for moderate-severe-critical COVID-19 | Inherited Metabolic Disorders |
| **Kamdar 2021 [115]** | Single-center retrospective cohort study conducted in USA (Jan/20 - Sep/20) | < 18 years  n = 109 | ICU | **ICU**: 8.8% (n = 9)  **Mechanical ventilation**: 6.4% (n = 7)  **ECMO**: 0.9% (n = 1)  **Death**: 1.8% (n = 2) – both had an oncologic diagnosis + active cancer therapy | Hematology/ Oncology |
| **Kearsley-Fleet 2022 [116]** | Multi-center multinational cohort study (Mar/20 - Jul/21) | < 19 years + adults  n = 607 pediatric | Hospitalization | **Hospitalization**: 7% (n = 43)  **Death**: 0.5% (n = 3)  Multivariate analysis  Comparator JIA  SLE/MCTD/Vasculitis: aOR 4.3 (1.7 - 11.0, p < 0.05)  Auto-inflammatory syndromes: aOR 3.0 (1.1 - 8.6, p < 0.05)  Obesity: aOR 4.0 (1.3 - 12.0, p < 0.05)  Not associated: sex, age, other rheumatologic diseases, glucocorticoid use, csDMARD only, b/tsDMARD only, combination therapy  *Adjusted for disease activity (remission, low, moderate to high, or unknown) and dataset (EULAR COVID-19 Registry or CARRA (CARRA Registry and CARRA COVID-19 GPRD))* | Rheumatology diagnosis (JIA, SLE, MCTD, vasculitis or other RMD, autoinflammatory syndromes, other) |
| **Ljungman 2023 [117]** | Multi-center multinational retrospective cohort study (Feb/20 - Jul/22) | < 18 years + adults  n = 114 pediatric | Death | **6-week overall survival**: 96.4% (89.3 = 98.1)  31 – 100 days after HSCT: HR 5.21 (2.81 – 9.63, p 0.003)  CMV positive serostatus: HR 2.38 (1.35 – 4.2, p 0.003)  Ongoing immune suppressive therapy: HR 2.18 (1.34 – 3.56, p 0.002)  Other lung disease: HR 2.47 (1.47 – 4.14, p 0.0006) | Allogenic hematopoietic stem cell transplant recipients |
| **Madhusoodhan 2020 [118]** | Multi-center cross-sectional study conducted in USA (Jan/20 - Apr/20) | ≤ 21 years  n = 98 | ICU | **ICU**: 23.2% (n = 17/73 hospitalized)  **Mechanical ventilation**: 8.2% (6/73 hospitalized)  **Death**: 4.08% (n = 4) – none were solely attributed to COVID-19 (three had relapsed/refractory disease and one had newly diagnosed AML with hyperleukocytosis, none of whom were in remission)  Obesity: (p 0.008)  Not associated: age > 15 years, sex (male) | Oncology patients receiving anticancer therapy |
| **Mann 2022 [119]** | Multi-center cross-sectional study conducted in USA (Apr/20 - Oct/21) | ≤ 24 years  n = 651 | ICU | **ICU**: 7.8% (n = 50)  **DKA at COVID-19 diagnosis**: 12.3% (n = 80)  **Median HbA1c**: 8.4% (IQR 7.3 – 10.2)  ≥ 1 comorbidity: aOR 2.12 (1.28 – 3.54, p 0.003)  Number of comorbidities (continuous): aOR 1.47 (1.15 – 186, p 0.001)  Asthma: aOR 2.68 (1.07 – 6.41, p 0.03)  Not associated: obesity, celiac disease  *Adjusted for age and Hba1c* | Type 1 diabetes |
| **Meena 2021 [120]** | Systematic literature review (Jan/20 - Oct/20) | < 18 years  n = 33 studies | Severe/Critical | **ICU**: 10.3% (n = 21/204 children)  **Death**: 4.9% (n = 11/226 children)  **MIS**-C: 3.4% (n = 7/204 children)  **Severe**: early respiratory symptoms, such as fever and cough, ± GI symptoms. Progression at ∼1 week, and dyspnea + central cyanosis. Sat O2 92% + hypoxia manifestations.  **Critical**: progression to ARDS or respiratory failure ± shock, encephalopathy, myocardial injury or heart failure, coagulation dysfunction, and AKI. Organ dysfunction can be life-threatening. | Undergoing/had undergone cancer treatment |
| **Millen 2022 [121]** | Multi-center cohort study conducted in UK (Mar/20 - Feb/21) | < 16 years  n = 110 | Severe/Critical | Hematological malignancies: 3.2% (n = 2/62)  Non-hematological malignancies: 6.3% (n = 3/48)  Compared with the non-hematological malignancy (CNS tumors, sarcomas, neuroblastomas and other, rarer tumors) subgroup.  Hematological malignancies: OR 0.5 (0.08 - 3.11)  Not associated: hematological vs. non-hematological malignancies | Diagnosis of any subtype of hematological malignancy including (but not limited to) all subtypes of leukemia and lymphoma as well as myelodysplastic syndromes.  Children who received stem cell transplantation were excluded |
| **Moeller 2020 [122]** | Multi-center multinational cohort study (Mar/20 - May/20) | < 18 years  n = 185 | ICU | **ICU**: 12.4% (n = 23)  Asthma: 7.9% (n = 5/63)  Cystic fibrosis: 21.4% (n = 3/14)  BPD: 22.2% (n = 2/9)  Other conditions: 39.4% (n = 13/33) | Chronic respiratory diseases  - Asthma (n = 63)  - Cystic fibrosis (n = 14)  - BPD (n = 9)  - Other (n = 33) |
| **Morford 2021 [123]** | Systematic literature review (Jan/20 - Oct/21) | Pediatric + Adults  n = 7 studies | ICU or death | All primary immunodeficiency conditions.  **Death**: 6 cohort studies indicate effect associated with increase in mortality  **ICU**: 2 cohort studies reported unadjusted and adjusted effects indicating increases risk for ICU admission; 1 cohort study reported an adjusted effect suggesting no difference in the risk  Underlying Primary Immunodeficiency (PID)  **Death**: 2 cohort studies reported an increased risk in patients with PID vs. no-PID  **ICU**: 2 cohort studies reported an increased risk in adults and children with PID vs. no-PID  Common Variable Immunodeficiency (CVID)  **Death**: 1 cohort study reported fatality rated suggesting an increase in mortality  Inborn Errors of Immunity (IEI)  **Death**: 3 cohort studies reported rates associated with an increased risk  Insufficient data: x-linked agammaglobulinemia (XLA), combined immunodeficiency (CID), severe combined immunodeficiency (SCID), selective IgA deficiency (SIgAD), age | Primary Immunodeficiency |
| **Mucalo 2021 [124]** | Multi-center multinational cohort study (Mar/20 - Mar/21) | < 18 years + adults  n = 364 pediatric | Moderate/Severe/Critical | **ICU**: 5.8% (n = 21)  **Death**: 0.3% (n = 1)  Multivariate analysis  Prior acute care visits for pain (> 2 vs. 0): RR 3.09 (1.59 - 6.03, p 0.0009)  SCD + albuminuria, decreased renal function, and/or SCR nephropathy: RR 3.67 (1.93 - 6.96, p < 0.0001)  SCD + acute chest syndrome in the last 3 years and/or pulmonary hypertension: RR 1.76 (1.06 - 2.92, p 0.03)  Not associated: hydroxyurea use, SCD + neuro (overt stroke, silent stroke, known CNS vasculopathy and/or abnormal transcranial Doppler result)  *Adjusted for age, sex, genotype, and medical history* | Sickle Cell Disease |
| **Mukkada 2021 [125]** | Multi-center multinational cohort study (Apr/20 - Feb/21) | < 19 years  n = 1,500 | Severe/Critical | **Severe**: requirement for a higher level of care (HLC) for any reason or any oxygen support needed greater than regular nasal cannula or facemask, but less than intubation.  **Critical**: organ dysfunction, intubation, or death  Induction phase: 30.4% (n = 51/168, p 0.001)  Relapse or refractory phase: 29.8% (n = 17/57, p 0.0001)  Not associated: consolidation phase, reinduction or interim maintenance, maintenance continuation, immunotherapy or cell therapy, recipient of HSCT, time since transplant  Multivariate analysis  LMIC: aOR 5.8 (3.8 - 8.8, p < 0.001)  UMIC: aOR 1.6 (1.2 - 2.2, p 0.0024)  ALC ≤ 300 cell/mm3: aOR 2.5 (1.8 - 3.4, p < 0.0001)  ANC ≤ 500 cell/mm3: aOR 1.8 (1.3 - 2.4, p 0.0001)  ≥ 1 comorbidities: aOR 1.4 (1.0 - 1.9, p 0.074)  Not associated: HIC, acute lymphoblastic leukemia/lymphoma, solid tumors, CNS, tumors, age < 1 year, age 1 - 9 years, age 10 - 14 years, age 15 - 18 years, sex  *Adjusted for income group, cancer type, age, labs, comorbidities, and intensive treatment* | Current or past diagnosis of cancer or who have received a hematopoietic stem-cell transplantation |
| **Nicastro 2021 [126]** | Systematic literature review |  |  | "Subsequently, different studies showed that patients on DMARDs are not at greater risk of severe SARS-CoV2 disease than the general population."  "Even in children with rheumatologic disease treated with DMARDs, there is no increased incidence of SARS-CoV-2 disease or its complications."  "... scarce susceptibility replicates that of the general pediatric population and denies a role of immunosuppression as an additional risk in transplanted children. Even data from the European Liver Transplant Registry demonstrate a protective effect of the tacrolimus on the development of severe disease in adults."  "[Patients affected by chronic renal diseases] have a high susceptibility to infections because of malnutrition, uremia, comorbidities, and of course, the immunosuppressive treatment itself."  "Data regarding the clinical manifestations of COVID-19 in pediatric hemato-oncological patients are still relatively scarce [...] in conclusion, pediatric cancer patients have overall good COVID-19 outcomes, but they are still slightly worse than the general population."  "According to these conclusions, immunosuppressive medications should not be withdrawn, and scheduled therapies should not be delayed in such children. Also, relevant scientific societies recommend in favor of the maintenance of standard care for children needing immunosuppression for diverse conditions." | Immunocompromise |
| **Nicastro 2022 [127]** | Literature review (Jan/20 - May/21) | < 18 years + adults | ICU, mechanical ventilation, CVS | "Children with end stage liver disease may experience hepatic decompensation during SARS-CoV2 infection."  "Since the proportion of NAFLD patients in published obese cohorts remains unknown, the question as to whether NAFLD is an independent risk factor of COVID-19 severity warrants further studies in children. Nevertheless, regarded adult data, children with NAFLD especially those with obesity, should be considered a risk group for severe COVID-19. [...] Children with CLD, including obese patients with suspected or documented NAFLD, may be at higher risk of developing severe COVID-19."  "Children after LT may not be at higher risk of severe complications of COVID-19 infection and that they present with similar symptoms and similar mild disease course as the general pediatric population." | Liver diseases: end-stage liver disease, liver transplant (LT) recipients and pre-transplant candidates |
| **Parker 2022 [128]** | Single-center retrospective and prospective cohort study conducted in USA (Mar/21 - Aug/21) | ≤ 24 years  n = 87 | **Critical**: Requiring life-sustaining treatment (non-IV, IV, or CVS) for ARDS, septic shock, or MIS-C | **ICU**: 7% (n = 6)  The low number of patients with severe/critical COVID-19 infection limited the ability to analyze associations with disease severity. | Patients with cancer who were treated with chemotherapy, HSCT, or CAR-T |
| **Rouger-Gaudichon 2020 [129]** | Multi-center cohort study conducted in France (Jan/20 - May/20) | < 18 years + adults  n = 31 | ICU | **ICU:** 16.1% (n = 5)  **Death:** 3.2% (n = 1)  4/5 had been heavily treated for their cancer and/or recently underwent HSCT | Hematologic malignancy or solid tumors who had undergone anti-cancer treatment in the past 6 months or who underwent HSCT for any reason and with immunosuppressive therapy either ongoing or interrupted less than 6 months prior |
| **Rouger-Gaudichon 2021 [130]** | Literature review | n = 19 studies | Require a higher level of care | "Infants under one year of age: the risk of serious forms of COVID-19 in infants has been reported. The test for SARS-CoV-2, possibly repeated, is absolutely necessary here. Again, the recommendation is to follow the current guidelines i.e. to follow the Interfant 06 protocol."  "Some reports suggest that patients with Down syndrome have a greater risk of developing severe COVID-19. Of note, this group benefits from an induction with "only'' 3 drugs in the CAALL-F01 protocol, including dexamethasone."  "Patients with relapsed ALL may be at greater risk of severe COVID-19."  "Patients who are the most at risk of developing COVID-19 complication (e.g. patients with recent HSCT history, relapsed leukemia under intensive treatment and any patient with significant comorbidity)."  "A recent study suggests that patients with solid tumors may have an effective immune response to SARS-CoV-2."  "... the immune response of patients with hematological cancers  seems to be impaired, particularly those with B-cell malignancies, which on the one hand may explain their vulnerability..." | Acute lymphoblastic leukemias |
| **Sachdeva 2021 [131]** | Multi-center retrospective cohort study conducted in India (Apr/20 - Aug/20) | < 18 years + adults  n = 83 pediatric | Death | **IMV**: 13.8% (n = 13)  **Death**: 13.8% (n = 13)  - 9/13 due to COVID-19  **Death**: COVID-19 positive 27.1% (13/48 hospitalized) vs. COVID-19 negative 9.2% (109/1,187 hospitalized)  Socioeconomic status (lower and lower-middle vs. upper-middle): 22.2% (n = 10) vs. 6.1% (n = 3), p 0.04  Severity of illness (presence of cyanotic spells, refractory heart failure, persistent shock, or requirement for ventilatory support): 72.7% (n = 9) vs. 5.1% (n = 4), p < 0.001  Not associated: sex, type of CHD, pulmonary hypertension, surgery during index hospitalization | Children with heart disease (CHD, acquired heart diseases such as rheumatic heart disease, previously diagnosed myocardial diseases) + Grown ups with congenital heart disease |
| **Schlage 2022 [132]** | Systematic review and meta-analysis (Dec/19 - Oct/21) | < 18 years  n = 55 studies | Severe | **Asymptomatic**: 23.9% (n = 239/1003 pediatric cancer patients)  **Mild or moderate**: 41.7% (n = 418/1003 pediatric cancer patients)  **Severe**: 11.1% (n = 111/1003)  **Death**: 2.5% (n = 25/1003) | Cancer |
| **Sengler 2021 [133]** | Multi-center cross-sectional study conducted in Germany (Apr/20 - Feb/21) | < 21 years  n = 76 | Hospitalization | Predominantly adolescents  **Mild**: 96%  "The clinical course of SARS-CoV-2 infection in patients with rheumatic diseases was mild in most juvenile patients with RMD, regardless of whether immunosuppressive therapy was maintained or discontinued." | Rheumatic and musculoskeletal diseases |
| **Strah 2022 [134]** | Multi-center cohort study conducted in USA (Apr/20 - Mar/21) | < 18 years + adults  n = 160 | Death | **Death**: CHD 3.8% (n = 6) vs. no-CHD 0.8% (n = 79), p < 0.001  Not associated: cyanotic and acyanotic moderate or severe CHD | Moderate or severe congenital heart disease |
| **Yeh 2023 [135]** | Multi-center multinational cohort study (Mar/20 - Dec/20) | < 18 years  n = 339 | ICU | **ICU**: 12% (n = 40)  **Death**: 5% (n = 18)  Multivariate analysis  Moderate clinical cardiac status: aOR 4.8 (1.82 - 12.7, p < 0.001)  Poor clinical cardiac status: aOR 24.0 (7.99 - 71.8, p < 0.001)  Recent cardiac intervention (cardiac surgery or catheterization within 90 days): aOR 5.67 (2.39 - 13.4, p < 0.001)  Not associated: age < 1 year, genetic syndrome, anatomic cardiac complexity, overall risk score | Congenital Heart Disease |
| **Zareef 2023 [136]** | Single-center cross-sectional study conducted in Lebanon (Jan/22 - Jul/22) | < 18 + adults  n = 124 | ICU or death | 96.2% of infected patients did not require hospitalization nor oxygen support.  ICU: 1.5% (n = 2)  Death: 0.8% (n = 1)  - 17-month-old with trisomy 21 and severe Tetralogy of Fallot, who required previous BT-shunt insertion during the neonatal period. Had history of recurrent hospitalization due to frequent upper respiratory tract infections. | Congenital Heart Disease |

#### **Supplementary Table 3**. Studies that provided estimates of risk adjusted by other comorbidities.

|  | **Statistically significant** | | **Not statistically significant** | |
| --- | --- | --- | --- | --- |
| **Risk factor** | **Study** | **aES (95%CI)** | **Study** | **aES (95%CI)** |
| Cardiovascular conditions | Madani 2021 | aOR 3.27 (2.09-5.11) | Campbell 2022 | aOR 2.30 (1.00-5.30) |
|  | Kompaniyets 2021 | aRR 1.72 (1.48-1.99) |  |  |
| Congenital heart disease | Saatci 2021 | aOR 9.4 (3.79-26.54) | Farrar 2022 | aRR 1.53 (0.78-2.97) |
|  | Kompaniyets 2021 | aRR 1.39 (1.19-1.63) |  |  |
| Pulmonary conditions | Madani 2021 | aOR 3.21 (1.59-6.47) |  |  |
|  | Campbell 2022 | aOR 2.50 (1.40-4.50) |  |  |
|  | Farrar 2022 | aRR 1.63 (1.12-2.39) |  |  |
| Asthma | Farrar 2022 ^a^ | aRR 2.24 (1.54-3.27) | Kompaniyets 2021 | aRR 1.09 (0.98-1.21) |
|  | Oliveira 2023-2 ^b^ | aHR 2.00 (1.51-2.65) | Farrar 2022 ^d^ | aRR 0.33 (0.05-2.20) |
|  |  |  | Oliveira 2023-2 ^c^ | aHR 0.39 (0.29-0.53) |
| Neurologic conditions | Farrar 2022 | aRR 1.84 (1.32-2.57) | Campbell 2022 | aOR 1.80 (0.90-3.30) |
| Neurodevelopmental disorders | Saatci 2021 | aOR 25.16 (4.53-139.83) | Kompaniyets 2021 | aRR 0.83 (0.70-0.98) |
|  | Ungar 2023 | aOR 5.45 (2.36-12.61) |  |  |
|  | Farrar 2022 | aRR 1.67 (1.05-2.64) |  |  |
|  | Leung 2023 (trisomy 21) | aOR 2.06 (1.39-3.01) |  |  |
| Seizure disorder | Saatci 2021 | aOR 6.81 (1.27-36.46) |  |  |
|  | Farrar 2022 | aRR 2.08 (1.44-2.99) |  |  |
|  | Kompaniyets 2021 | aRR 1.71 (1.41-2.08) |  |  |
| Prematurity | Ungar 2023 | aOR 4.95 (1.70-14.43) |  |  |
|  | Farrar 2022 | aRR 3.47 (1.69-7.09) |  |  |
| Diabetes | Banull 2022 | aOR 8.23 (2.79-24.27) | Kompaniyets 2021 ^f^ | aRR 1.21 (0.98-1.49) |
|  | Madani 2021 | aOR 2.50 (1.38-4.55) |  |  |
|  | Oliveira 2022 | aHR 1.53 (1.17-2.03) |  |  |
|  | Kompaniyets 2021 ^e^ | aRR 2.38 (2.06-2.76) |  |  |
| Obesity | Campbell 2022 | aOR 4.70 (2.80-7.60) | Farrar 2022 | aRR 1.33 (0.86-2.06) |
|  | Banull 2022 | aOR 2.76 (1.30-5.89) |  |  |
|  | Kompaniyets 2021 | aRR 1.42 (1.22-1.66) |  |  |
| Immunocompromise |  |  | Campbell 2022 | aOR 2.20 (1.00-4.80) |
|  |  |  | Madani 2021 | aOR 1.06 (0.29-3.89) |
|  |  |  | Farrar 2022 | aRR 0.43 (0.16-1.16) |
| Oncologic diagnosis | Ungar 2023 | aOR 4.25 (1.55-11.62) | Madani 2021 | aOR 1.88 (0.88-3.98) |
|  |  |  | Farrar 2022 | aRR 0.65 (0.22-1.90) |
| Chronic GI conditions |  |  | Kompaniyets 2021 | aRR 1.04 (0.90-1.20) |
|  |  |  | Farrar 2022 | aRR 0.77 (0.30-1.98) |
| Liver conditions |  |  | Madani 2021 | aOR 2.90 (0.96-8.84) |
| Chronic kidney disease |  |  | Farrar 2022 | aRR 0.78 (0.26-2.32) |
|  |  |  | Campbell 2022 | aOR 0.40 (0.10-3.70) |
| Sickle cell disease |  |  | Farrar 2022 | aRR 0.20 (0.03-1.39) |

**Statistically significant**: confidence interval does not cross the null, p-value < 0.05. **aES (95% CI):** adjusted effect size with 95% confidence interval extracted from published studies. **aOR**: adjusted odds ratio. **aRR**: adjusted risk ratio. **aHR**: adjusted hazard ratio. ^a^Risk factor = asthma without controller medication. ^b^Risk factor = asthma with other comorbidities. ^c^Risk factor = asthma without other comorbidities. ^d^Risk factor = asthma with controller medication. ^e^Risk factor = type 1 diabetes mellitus. ^f^risk factor = type 2 diabetes mellitus.

### **Section 4: Meta-Analysis of the Association Between Risk Factors and Critical COVID-19 in Children**

#### **Supplementary Figure 1**. Age < 1 year

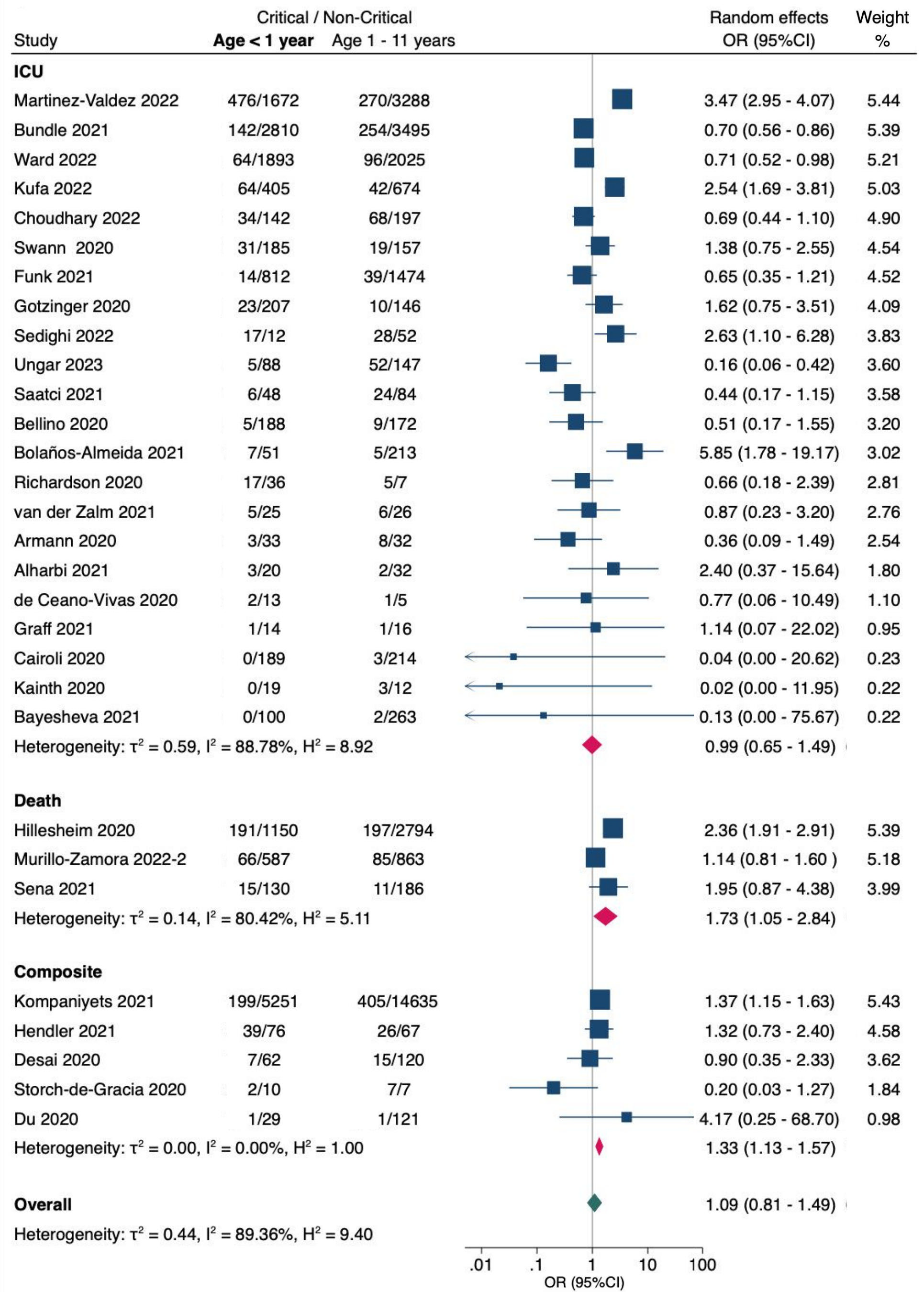

**OR (95% CI)** = odds ratio with 95% confidence interval, estimated using extracted data from published studies.

**ICU** = admission to intensive care unit; **Composite** = combined critical outcomes (ICU, IMV, CVS, or death).

#### **Supplementary Figure 2**. Age < 1 month

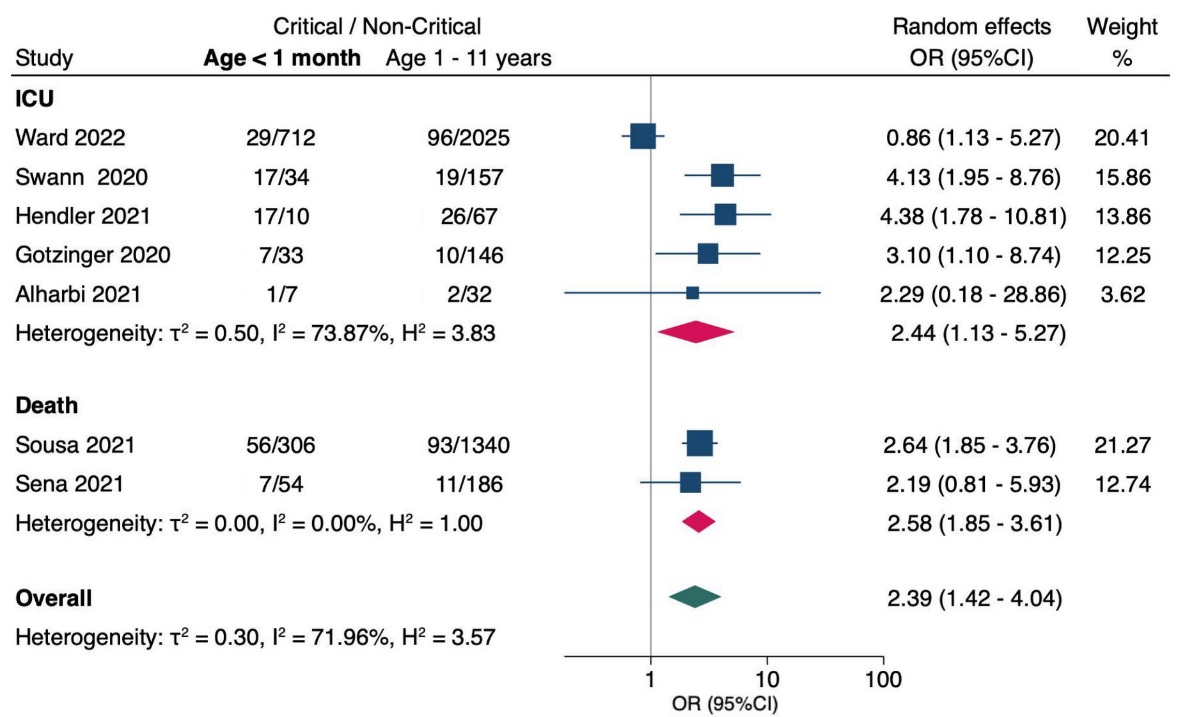

**OR (95% CI)** = odds ratio with 95% confidence interval, estimated using extracted data from published studies.

**ICU** = admission to intensive care unit.

#### **Supplementary Figure 3**. Prematurity

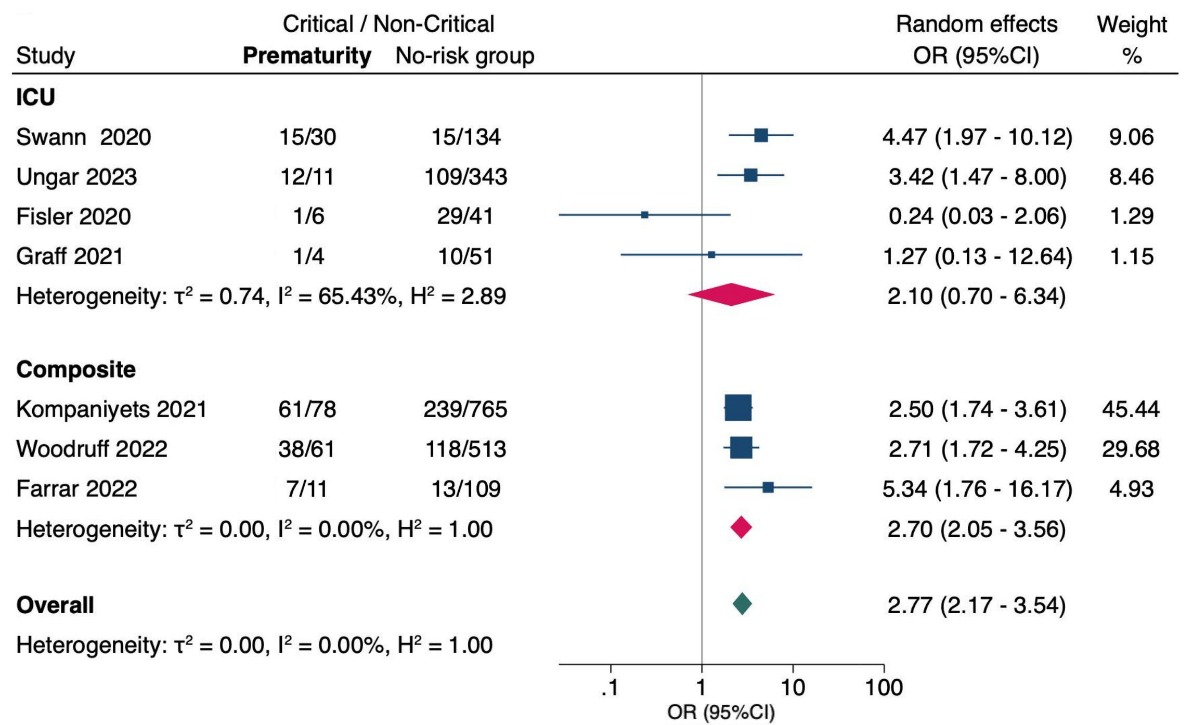

**OR (95% CI)** = odds ratio with 95% confidence interval, estimated using extracted data from published studies.

**ICU** = admission to intensive care unit; **Composite** = combined critical outcomes (ICU, IMV, CVS, or death).

#### **Supplementary Figure 4**. Age ≥ 12 years

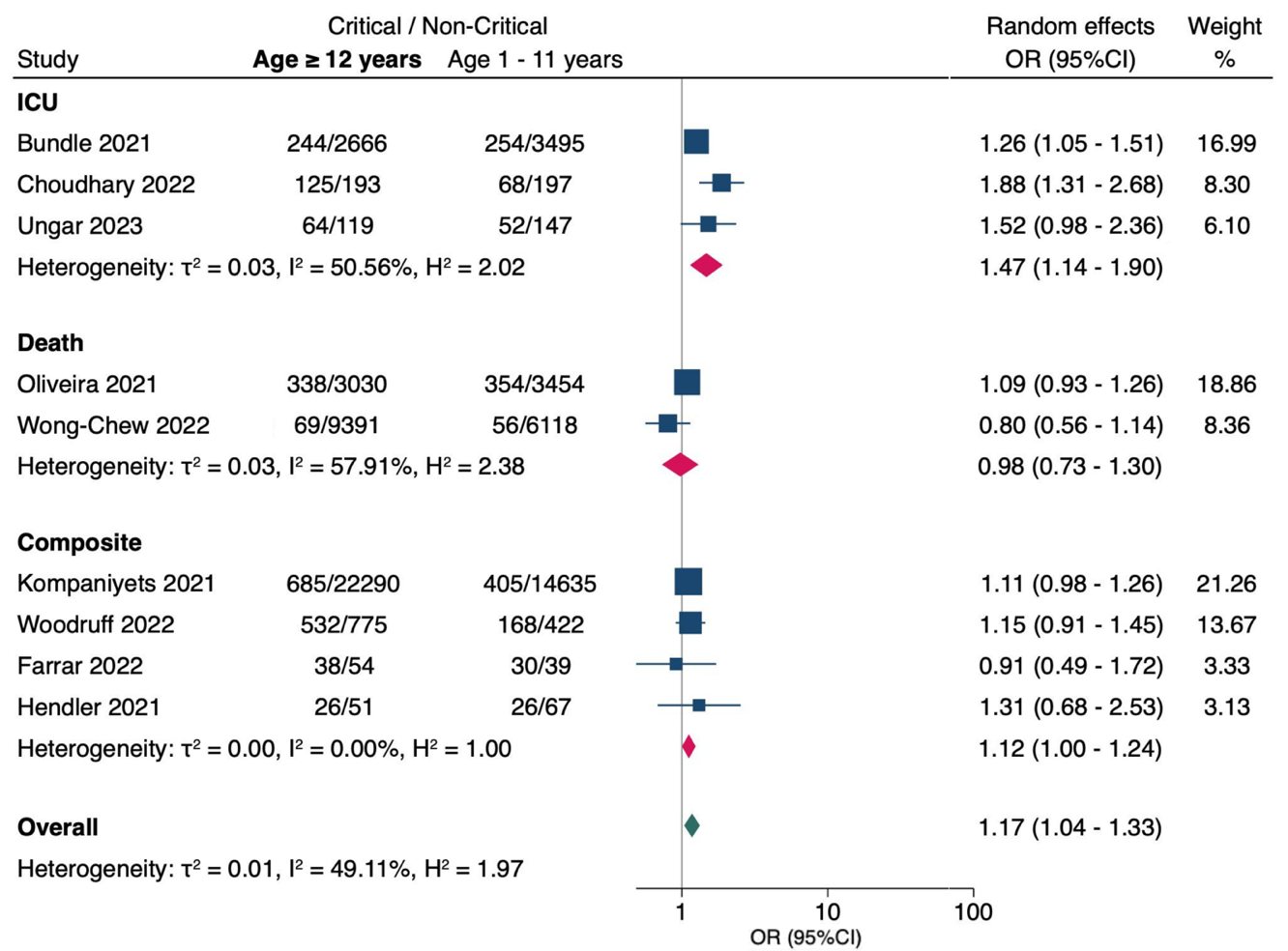

**OR (95% CI)** = odds ratio with 95% confidence interval, estimated using extracted data from published studies.

**ICU** = admission to intensive care unit; **Composite** = combined critical outcomes (ICU, IMV, CVS, or death).

**Supplementary Figure 5**. Sex (Reference. Female)

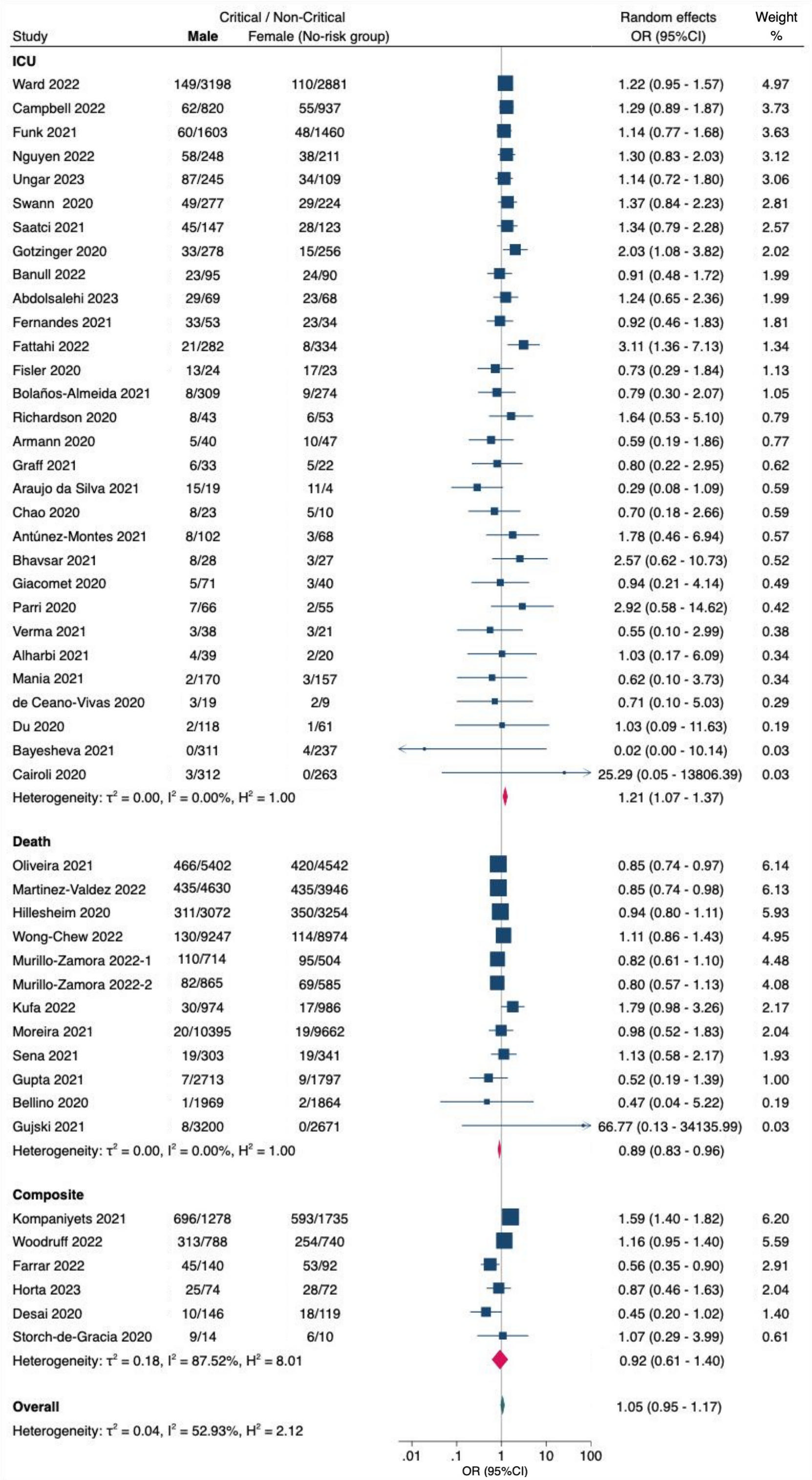

**OR (95% CI)** = odds ratio with 95% confidence interval, estimated using extracted data from published studies.

**ICU** = admission to intensive care unit; **Composite** = combined critical outcomes (ICU, IMV, CVS, or death).

#### **Supplementary Figure 6**. Cardiovascular conditions

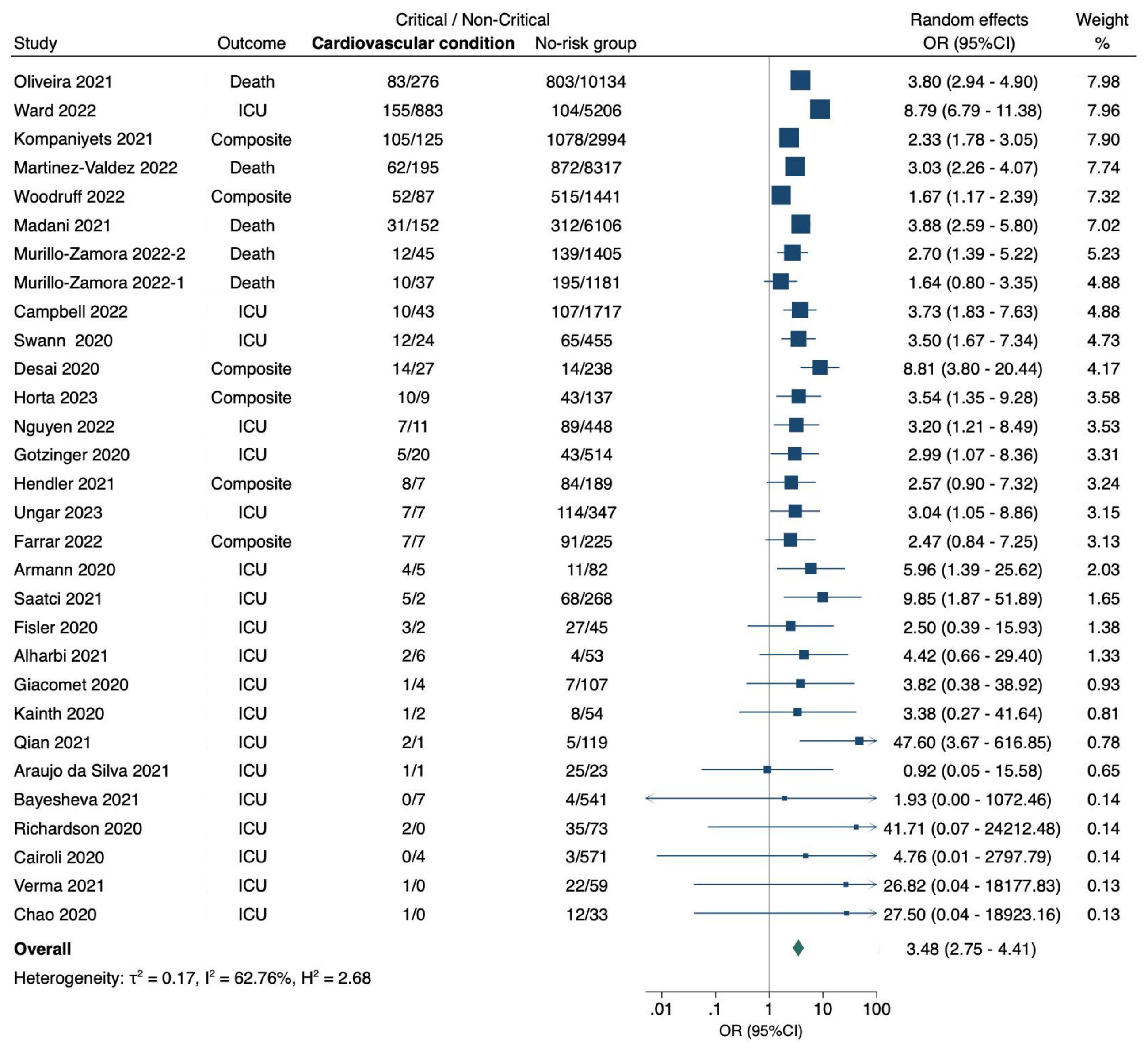

**OR (95% CI)** = odds ratio with 95% confidence interval, estimated using extracted data from published studies.

**ICU** = admission to intensive care unit; **Composite** = combined critical outcomes (ICU, IMV, CVS, or death).

#### **Supplementary Figure 7**. Congenital heart disease

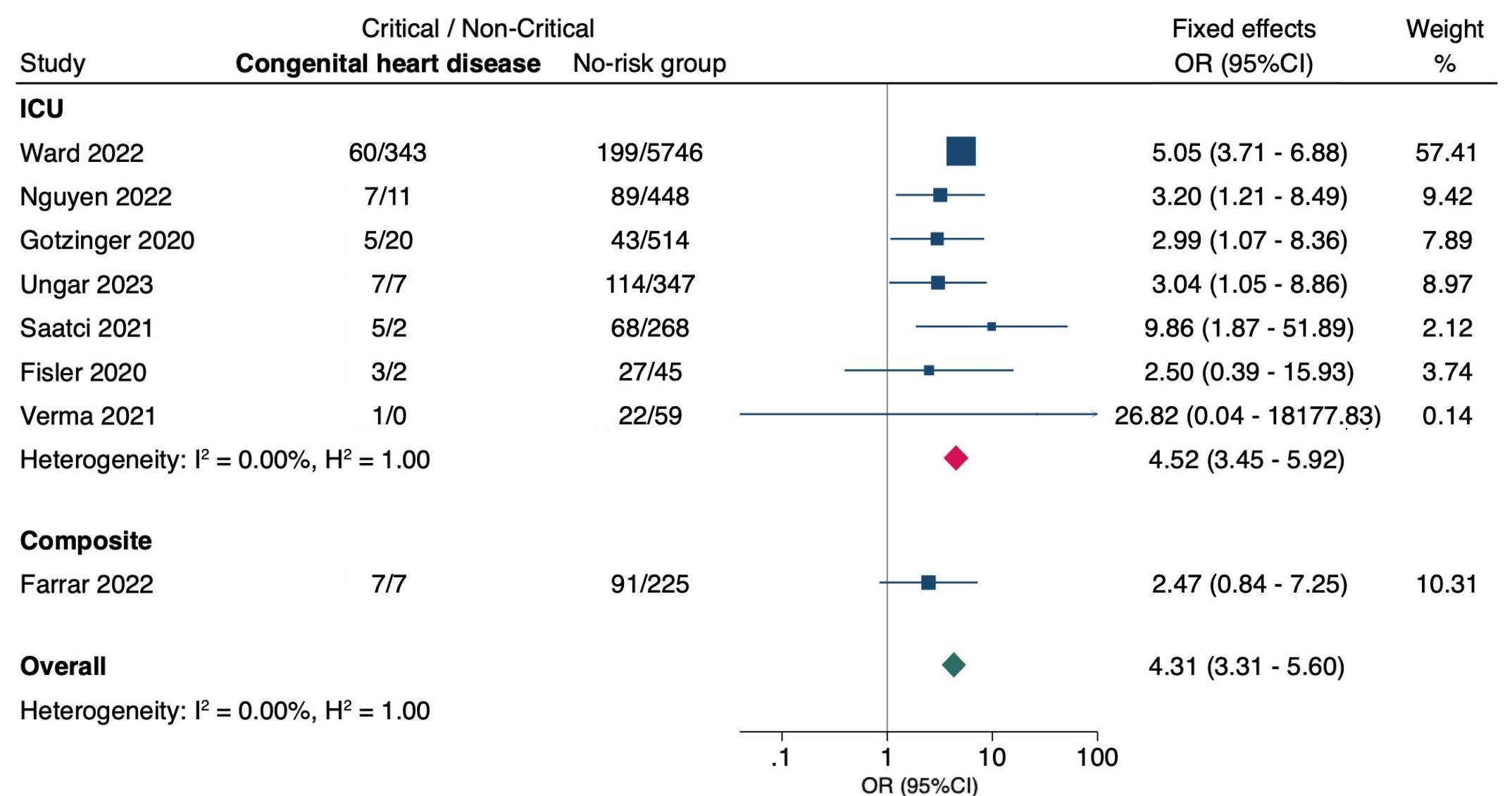

Fixed effects model was used due to convergence error with Random effects model.

**OR (95% CI)** = odds ratio with 95% confidence interval, estimated using extracted data from published studies.

**ICU** = admission to intensive care unit; **Composite** = combined critical outcomes (ICU, IMV, CVS, or death).

#### **Supplementary Figure 8**. Hypertension

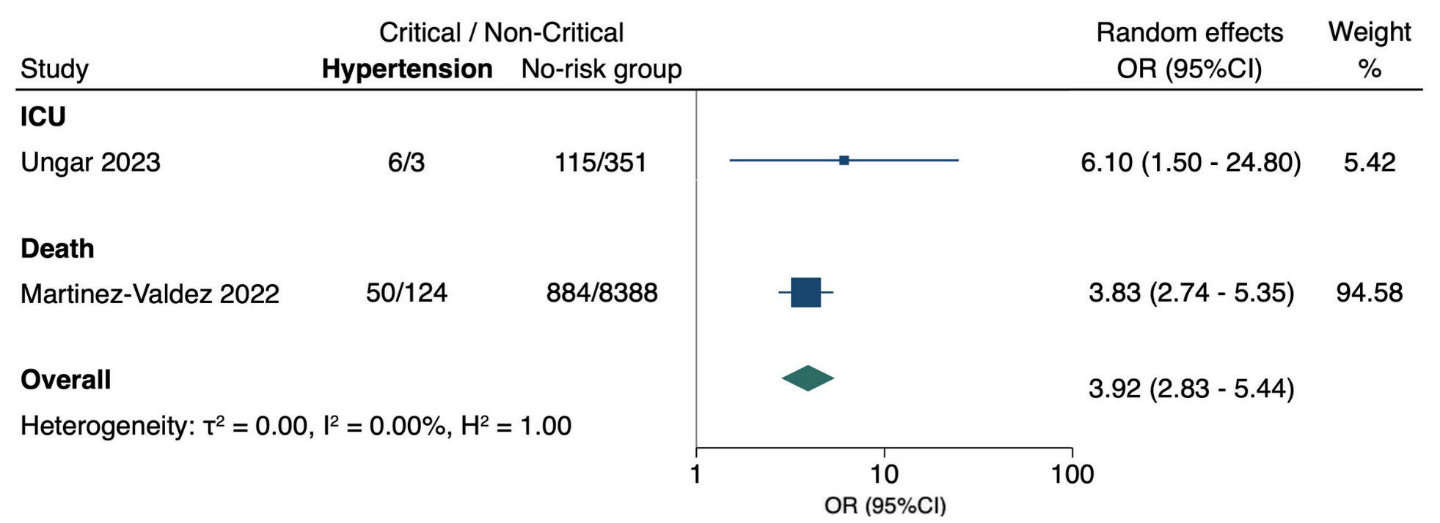

**OR (95% CI)** = odds ratio with 95% confidence interval, estimated using extracted data from published studies.

**ICU** = admission to intensive care unit.

#### **Supplementary Figure 9**. Pulmonary conditions

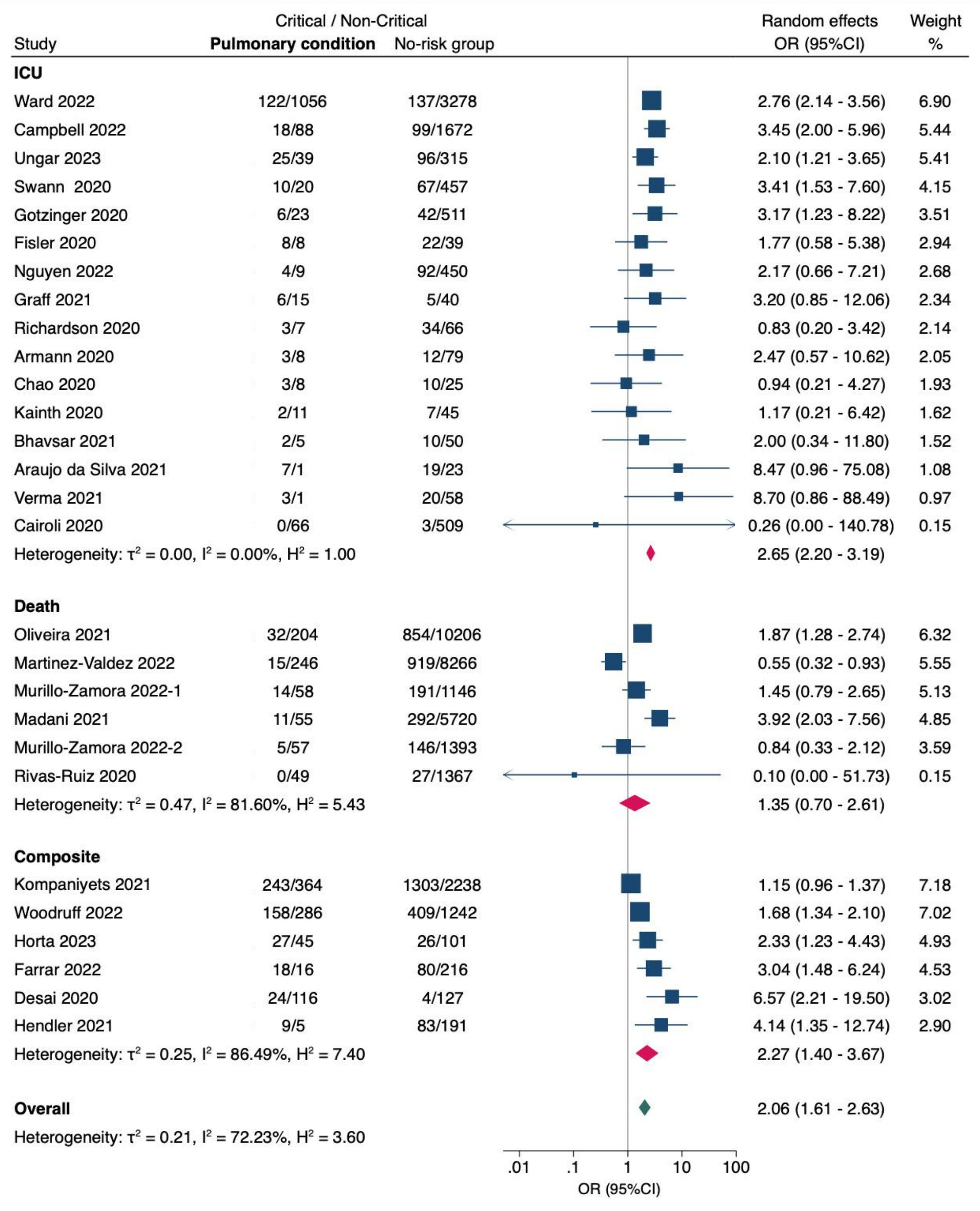

**OR (95% CI)** = odds ratio with 95% confidence interval, estimated using extracted data from published studies.

**ICU** = admission to intensive care unit; **Composite** = combined critical outcomes (ICU, IMV, CVS, or death).

#### **Supplementary Figure 10**. Asthma

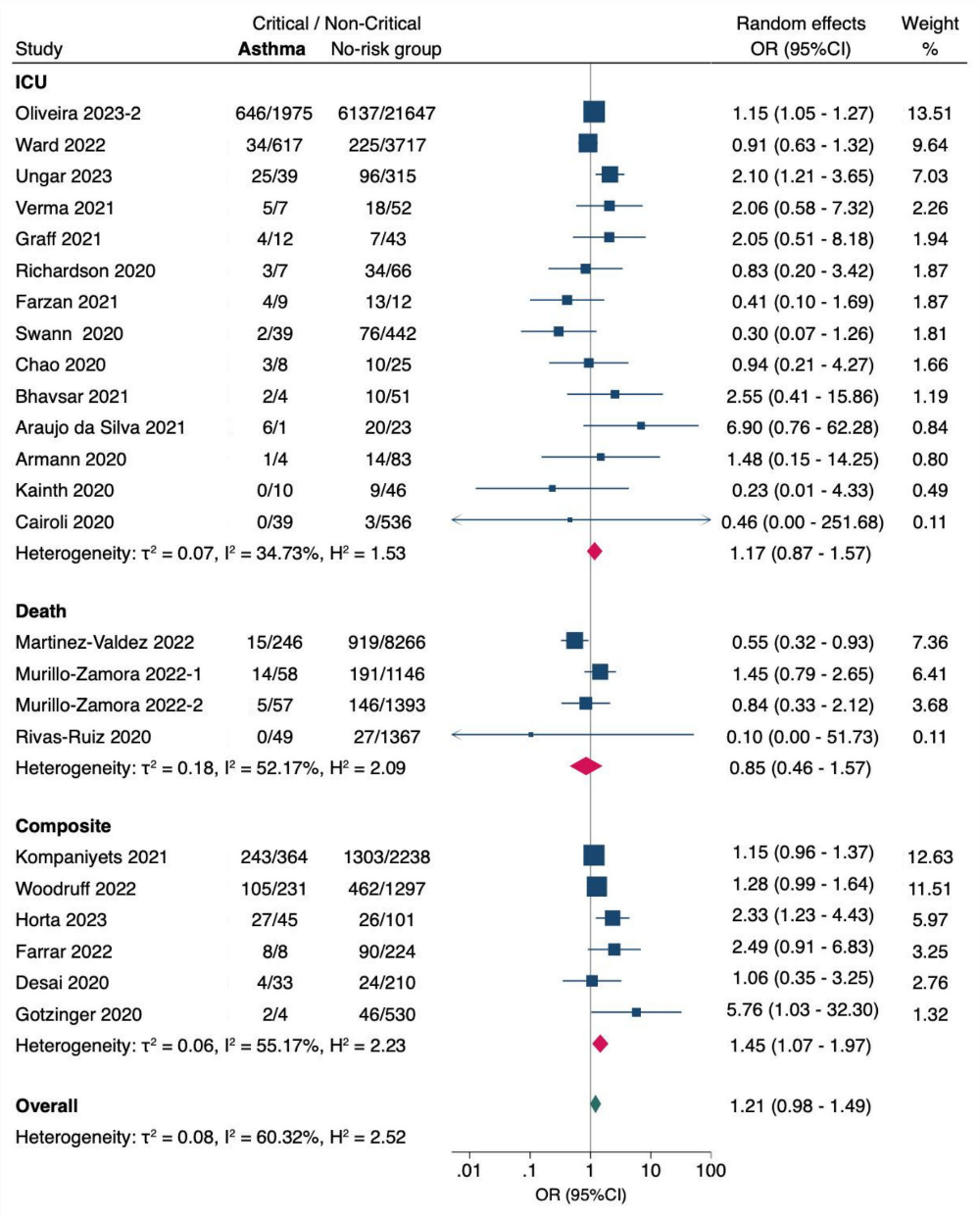

**OR (95% CI)** = odds ratio with 95% confidence interval, estimated using extracted data from published studies.

**ICU** = admission to intensive care unit; **Composite** = combined critical outcomes (ICU, IMV, CVS, or death).

#### **Supplementary Figure 11**. Pulmonary conditions excluding asthma

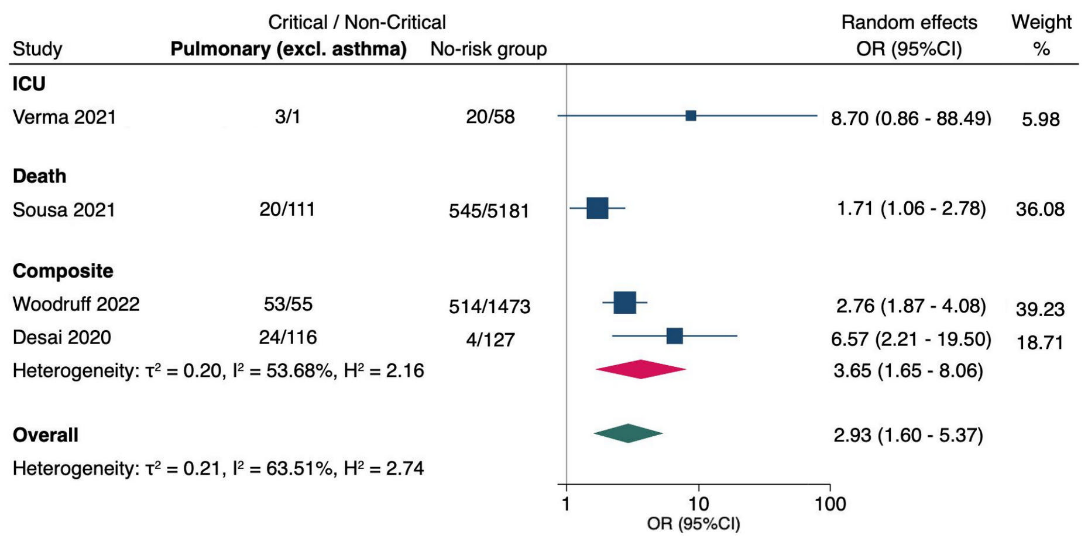

**OR (95% CI)** = odds ratio with 95% confidence interval, estimated using extracted data from published studies.

**ICU** = admission to intensive care unit; **Composite** = combined critical outcomes (ICU, IMV, CVS, or death).

#### **Supplementary Figure 12**. Neurologic conditions

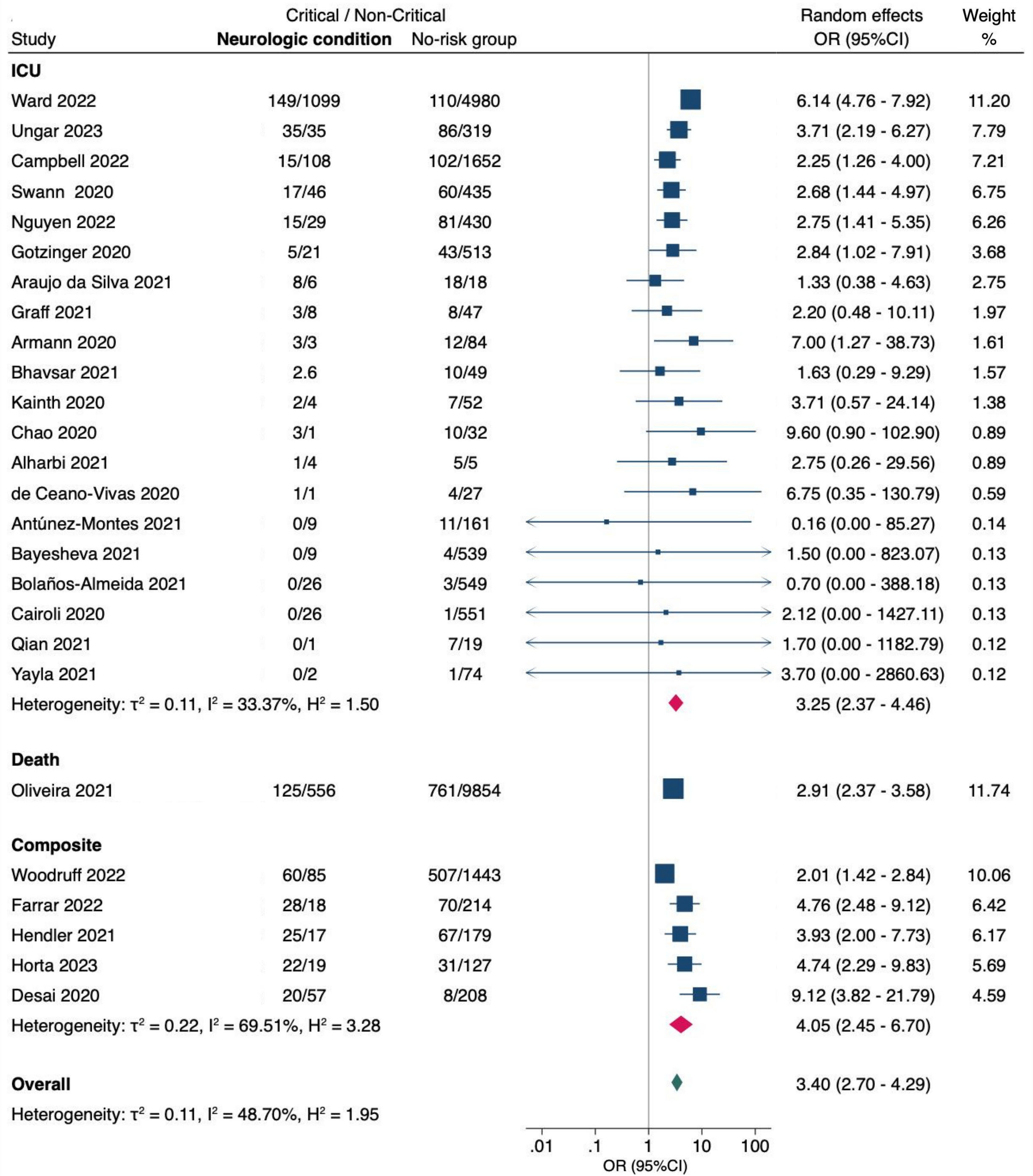

**OR (95% CI)** = odds ratio with 95% confidence interval, estimated using extracted data from published studies.

**ICU** = admission to intensive care unit; **Composite** = combined critical outcomes (ICU, IMV, CVS, or death).

#### **Supplementary Figure 13**. Seizure disorders

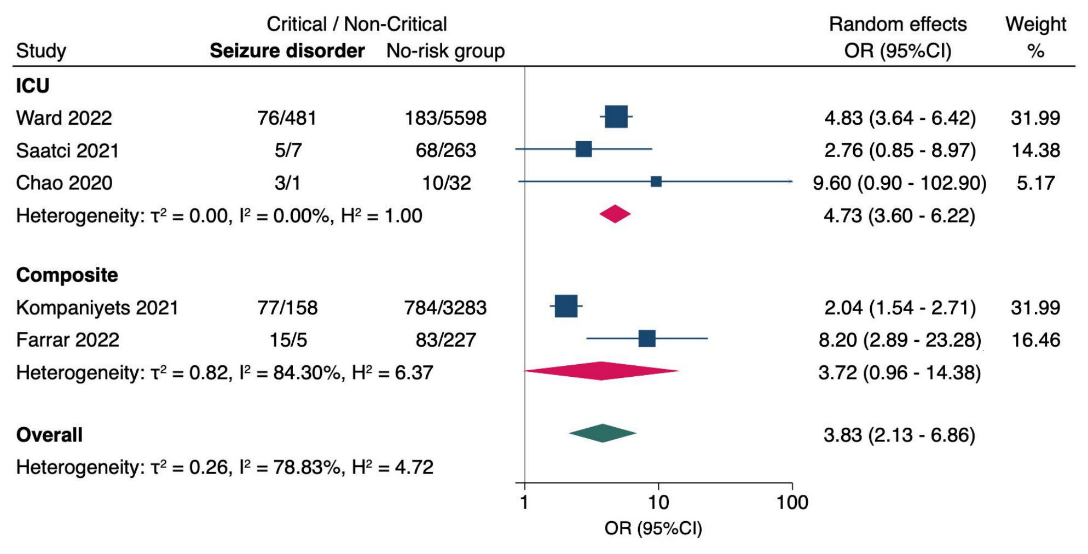

**OR (95% CI)** = odds ratio with 95% confidence interval, estimated using extracted data from published studies.

**ICU** = admission to intensive care unit; **Composite** = combined critical outcomes (ICU, IMV, CVS, or death).

#### **Supplementary Figure 14**. Neurodevelopmental disorders

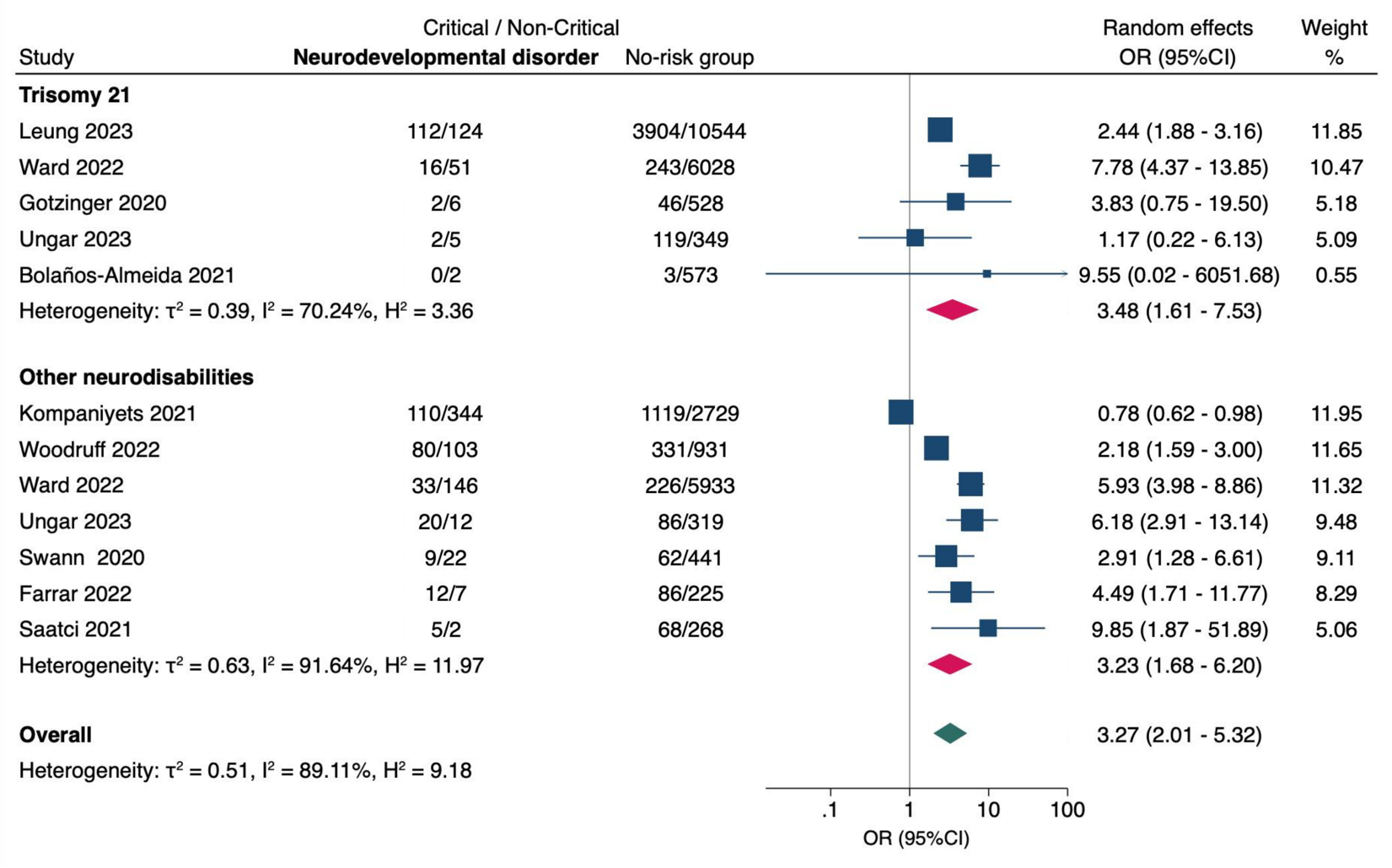

**OR (95% CI)** = odds ratio with 95% confidence interval, estimated using extracted data from published studies.

#### **Supplementary Figure 15**. Diabetes

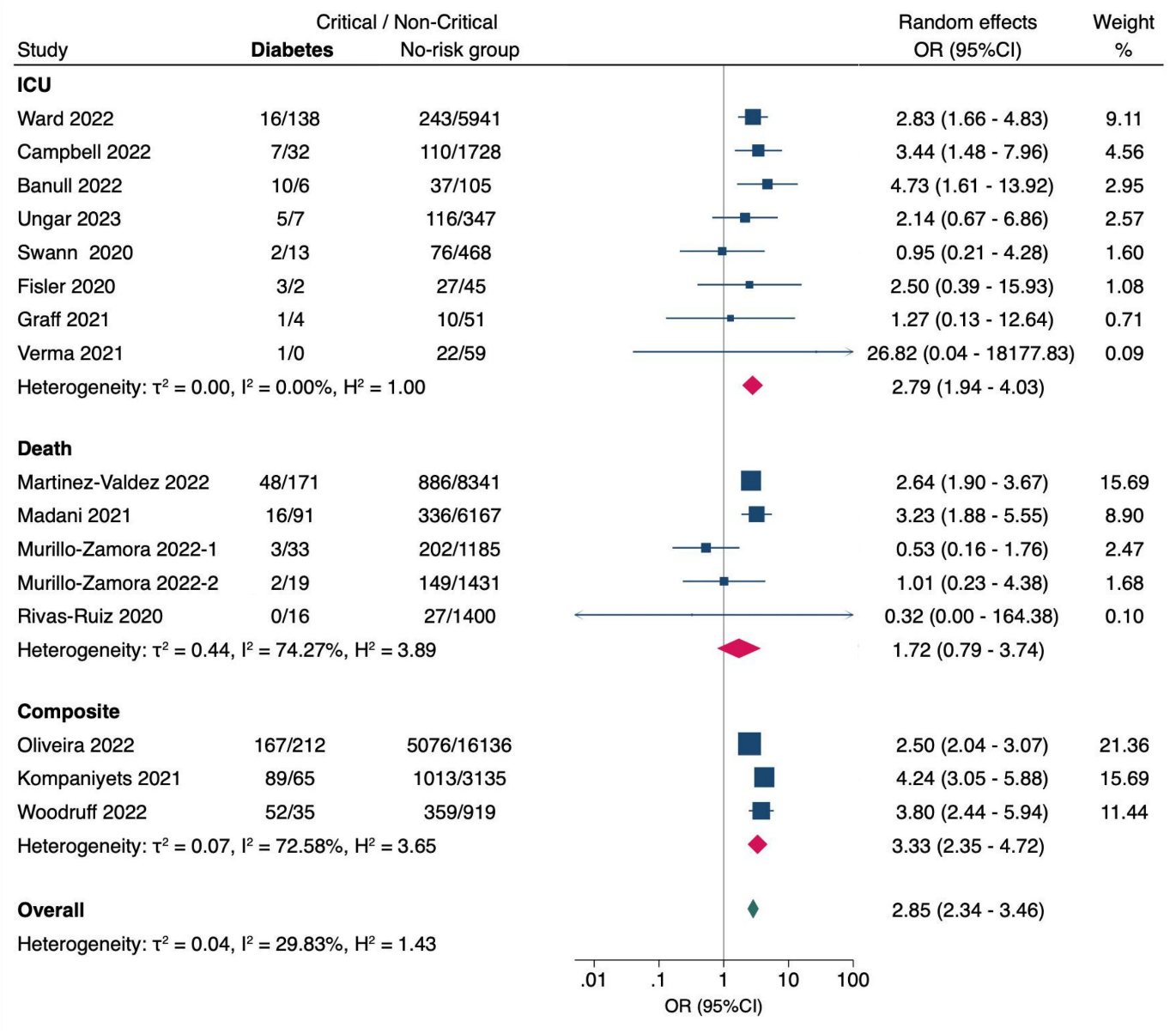

**OR (95% CI)** = odds ratio with 95% confidence interval, estimated using extracted data from published studies.

**ICU** = admission to intensive care unit; **Composite** = combined critical outcomes (ICU, IMV, CVS, or death).

#### **Supplementary Figure 16**. Obesity

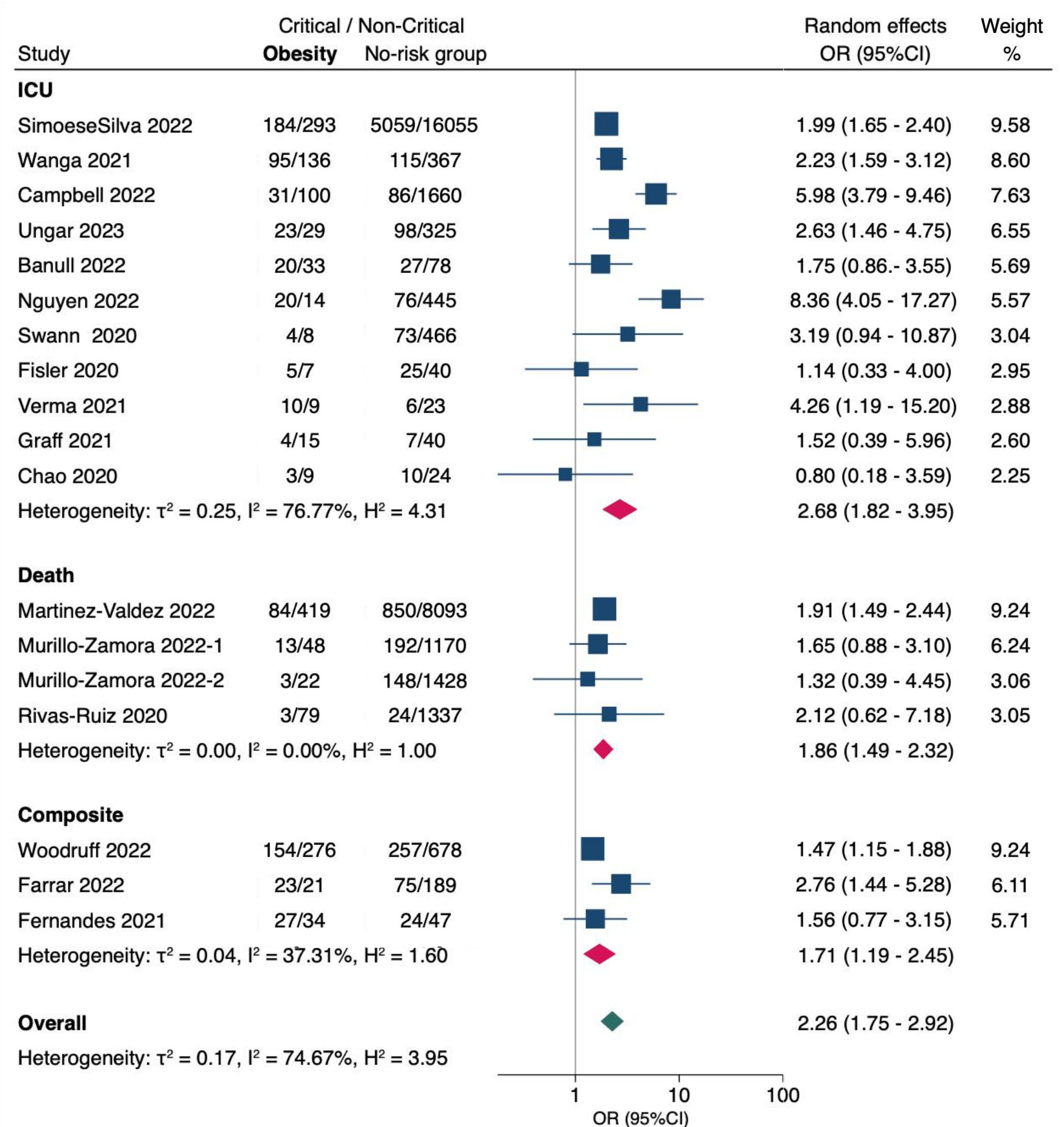

**OR (95% CI)** = odds ratio with 95% confidence interval, estimated using extracted data from published studies.

**ICU** = admission to intensive care unit; **Composite** = combined critical outcomes (ICU, IMV, CVS, or death).

#### **Supplementary Figure 17**. Immunocompromise

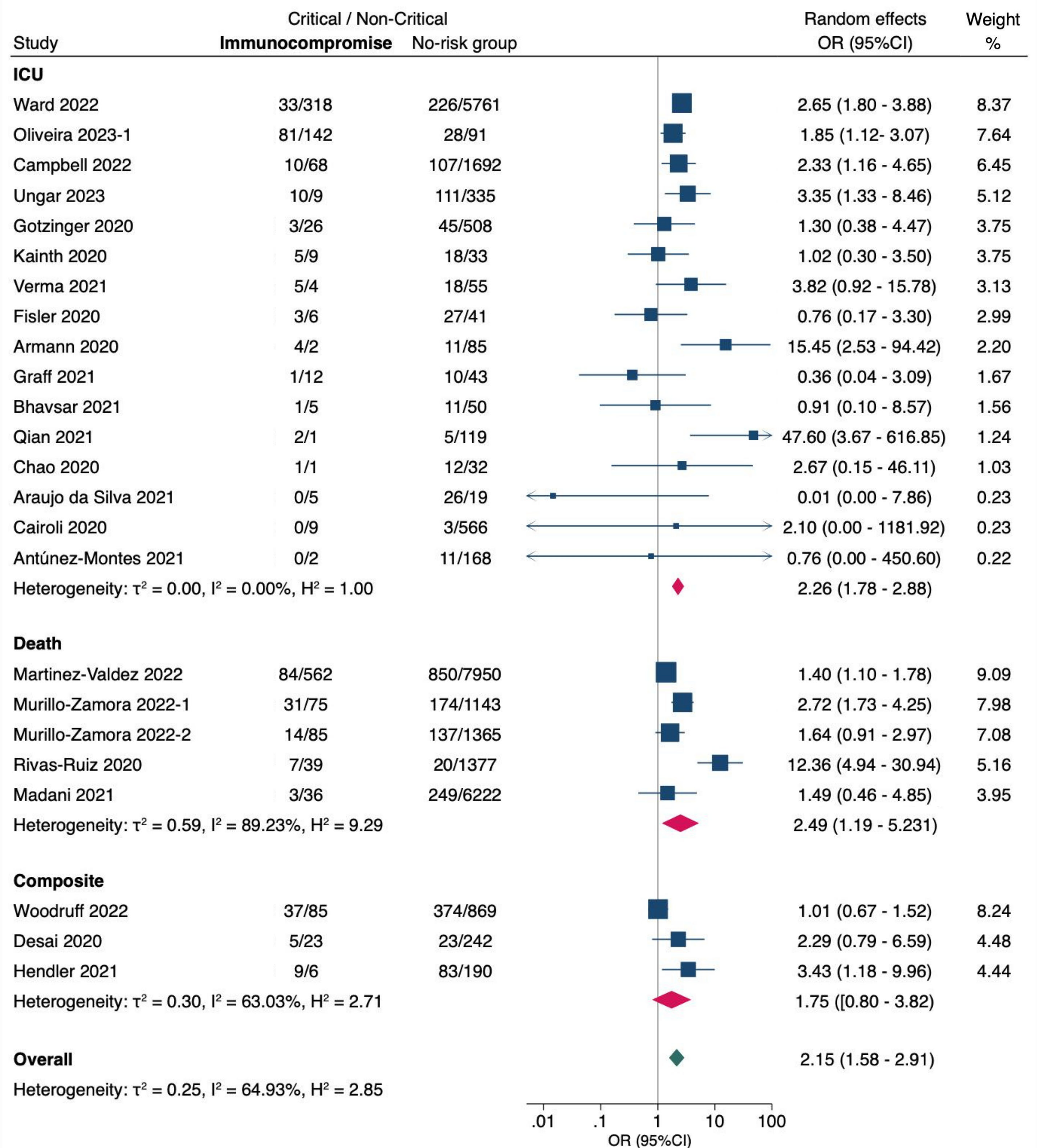

**OR (95% CI)** = odds ratio with 95% confidence interval, estimated using extracted data from published studies.

**ICU** = admission to intensive care unit; **Composite** = combined critical outcomes (ICU, IMV, CVS, or death).

#### **Supplementary Figure 18**. Oncologic diagnosis

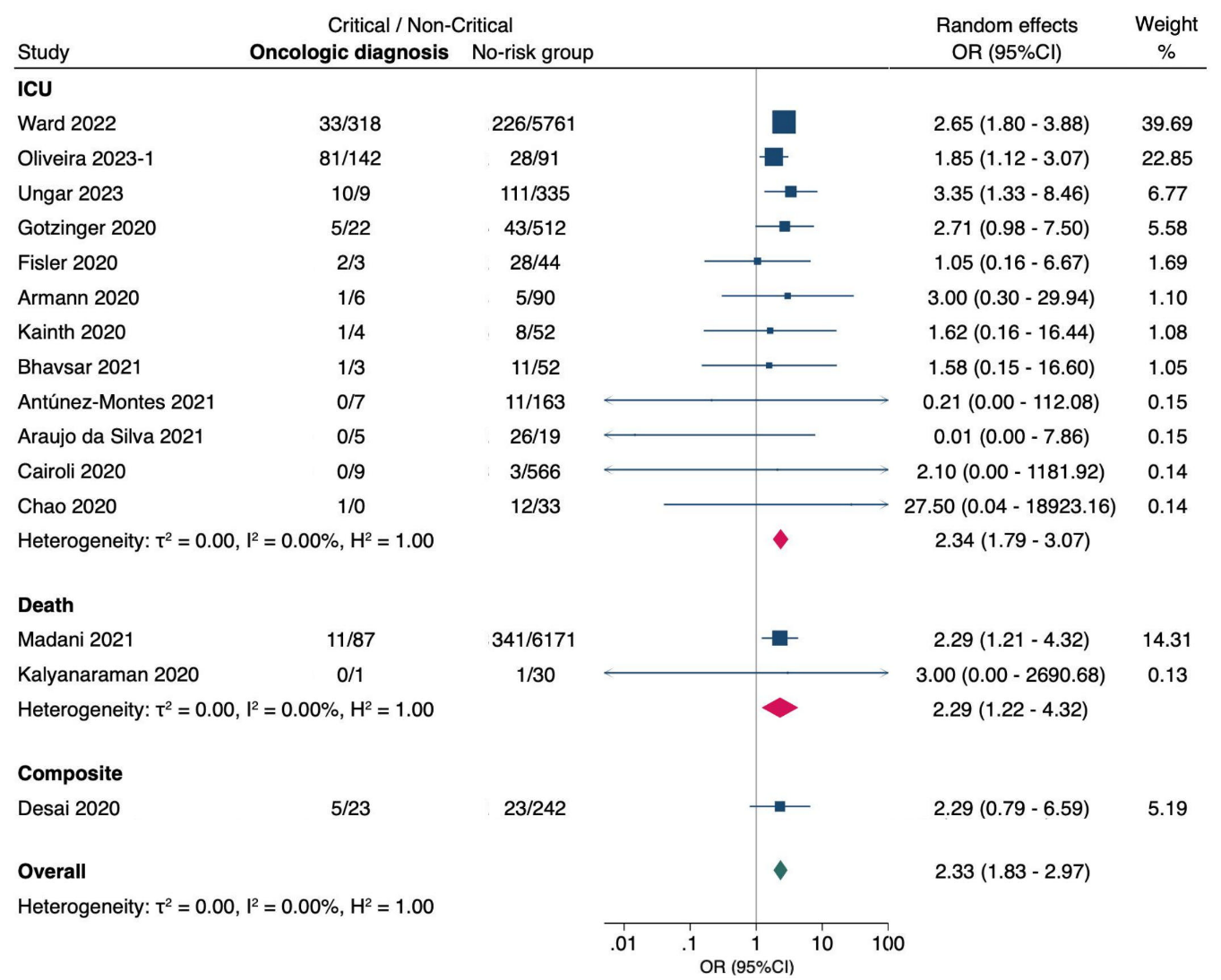

**OR (95% CI)** = odds ratio with 95% confidence interval, estimated using extracted data from published studies.

**ICU** = admission to intensive care unit; **Composite** = combined critical outcomes (ICU, IMV, CVS, or death).

#### **Supplementary Figure 19**. Rheumatologic conditions

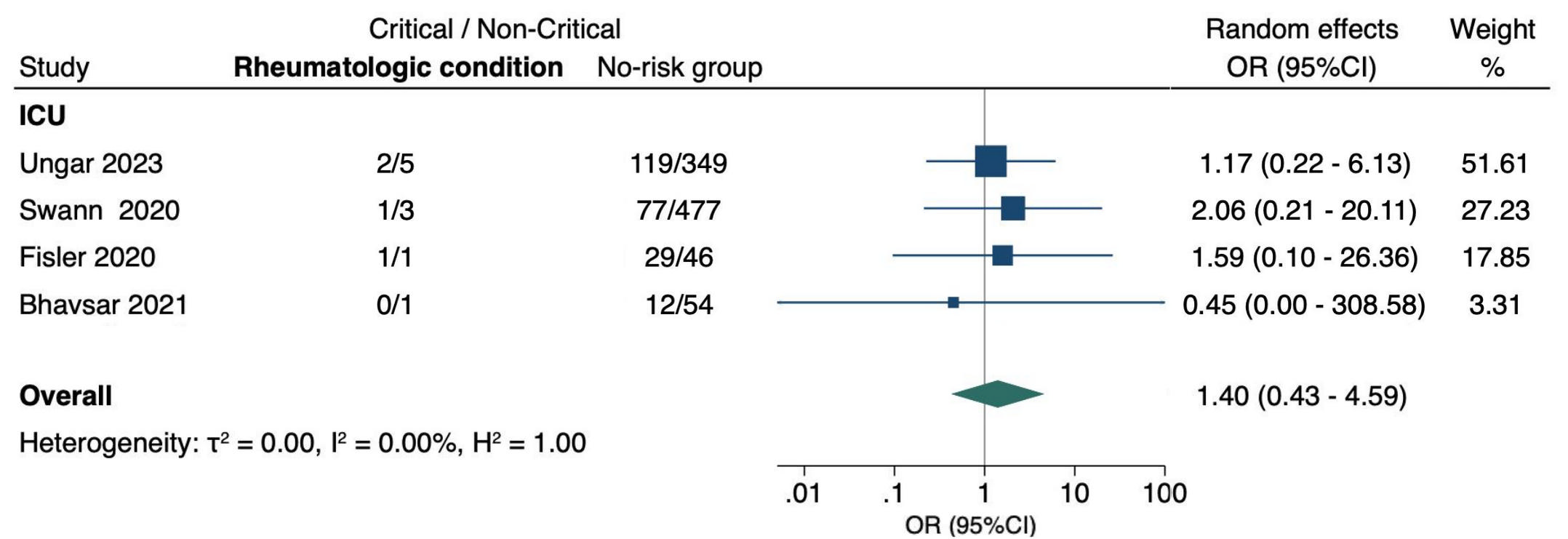

**OR (95% CI)** = odds ratio with 95% confidence interval, estimated using extracted data from published studies.

**ICU** = admission to intensive care unit.

#### **Supplementary Figure 20**. Chronic gastrointestinal conditions

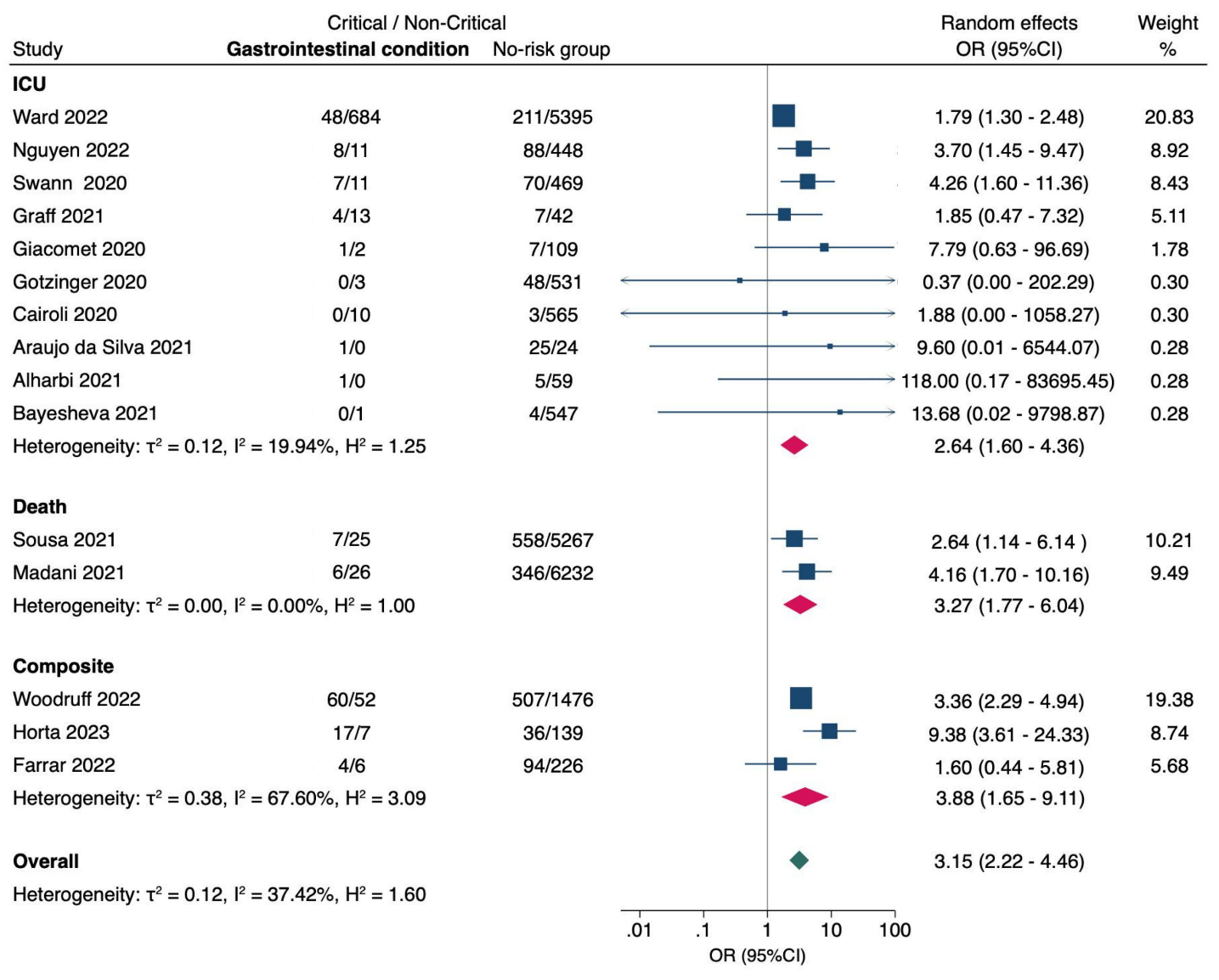

**OR (95% CI)** = odds ratio with 95% confidence interval, estimated using extracted data from published studies.

**ICU** = admission to intensive care unit; **Composite** = combined critical outcomes (ICU, IMV, CVS, or death).

**Supplementary Figure 21**. Hepatic conditions

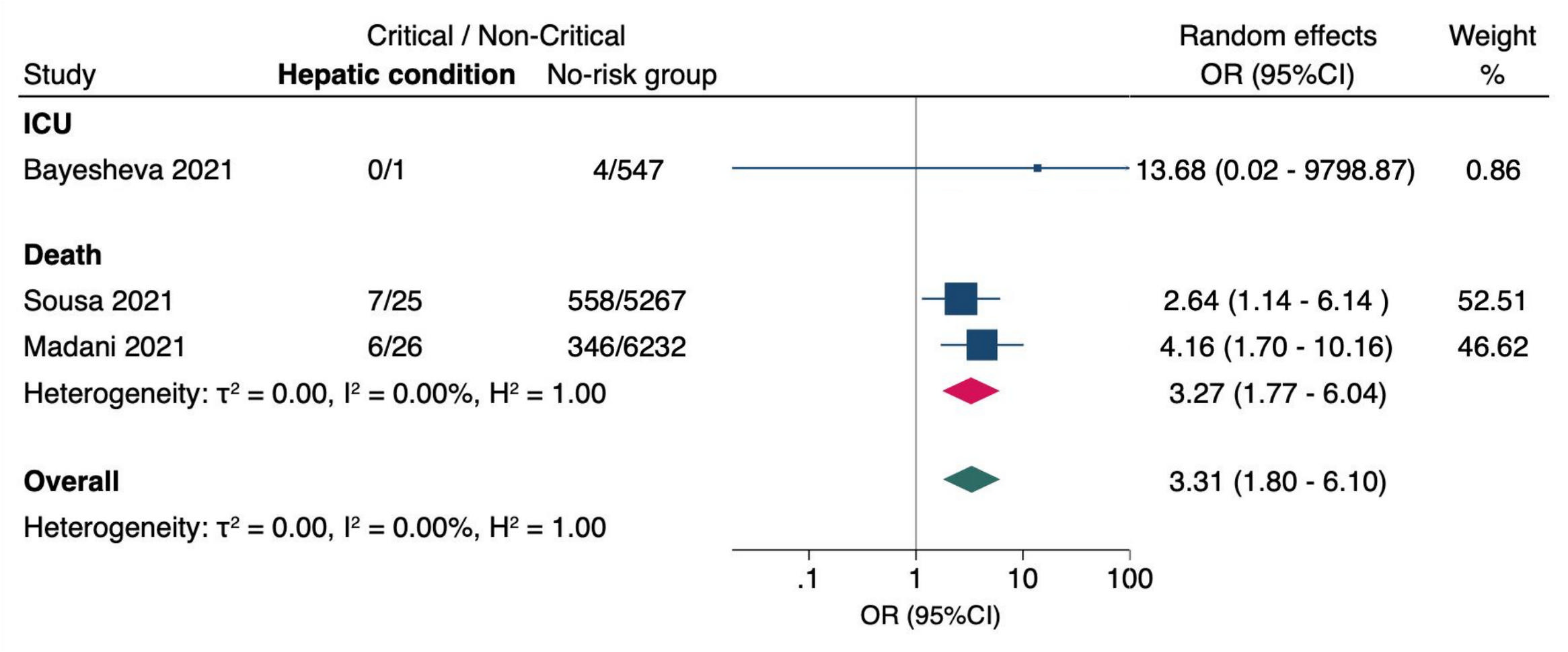

**OR (95% CI)** = odds ratio with 95% confidence interval, estimated using extracted data from published studies.

**ICU** = admission to intensive care unit.

#### **Supplementary Figure 22**. Chronic kidney disease

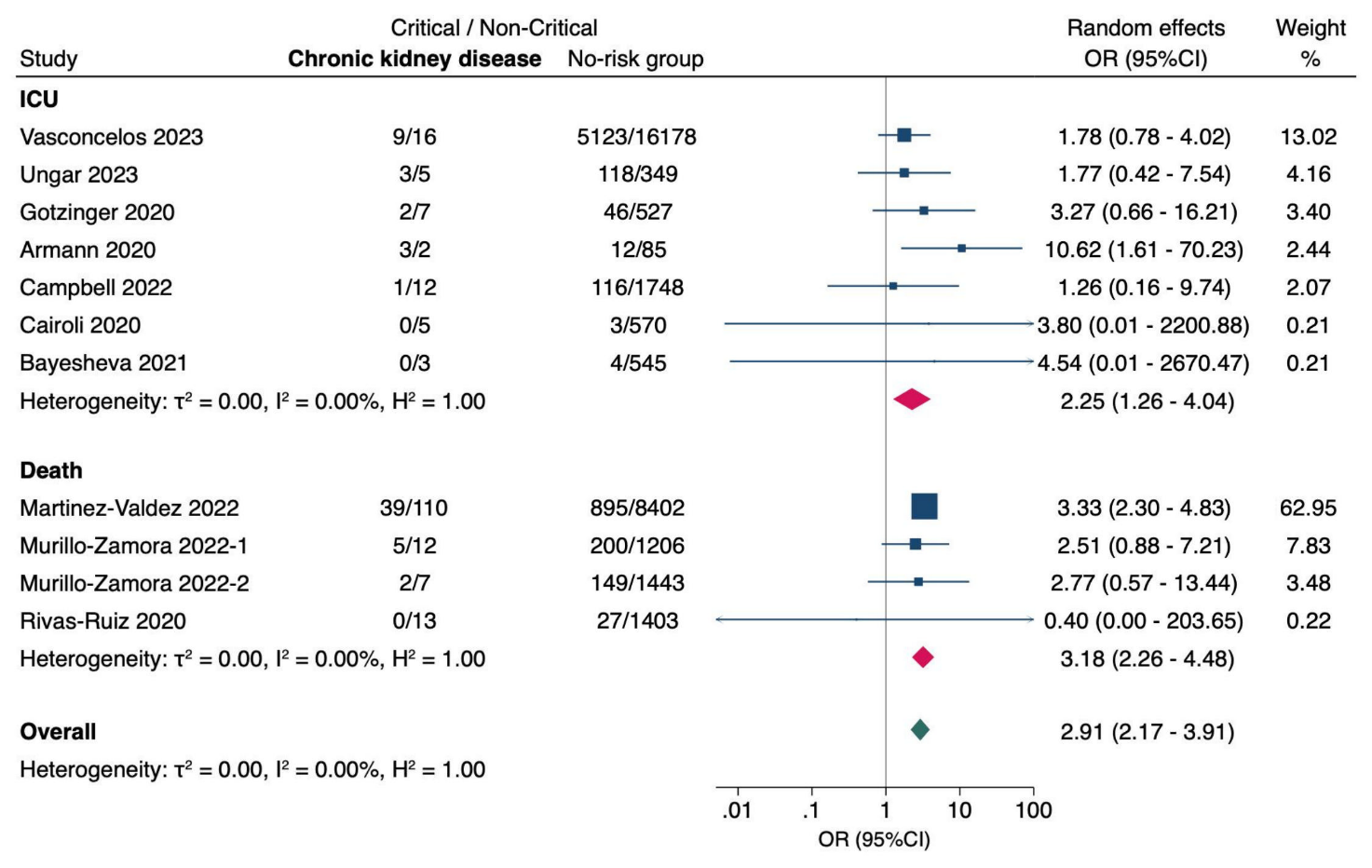

**OR (95% CI)** = odds ratio with 95% confidence interval, estimated using extracted data from published studies.

**ICU** = admission to intensive care unit; **Composite** = combined critical outcomes (ICU, IMV, CVS, or death).

**Supplementary Figure 23**. Sickle cell disease

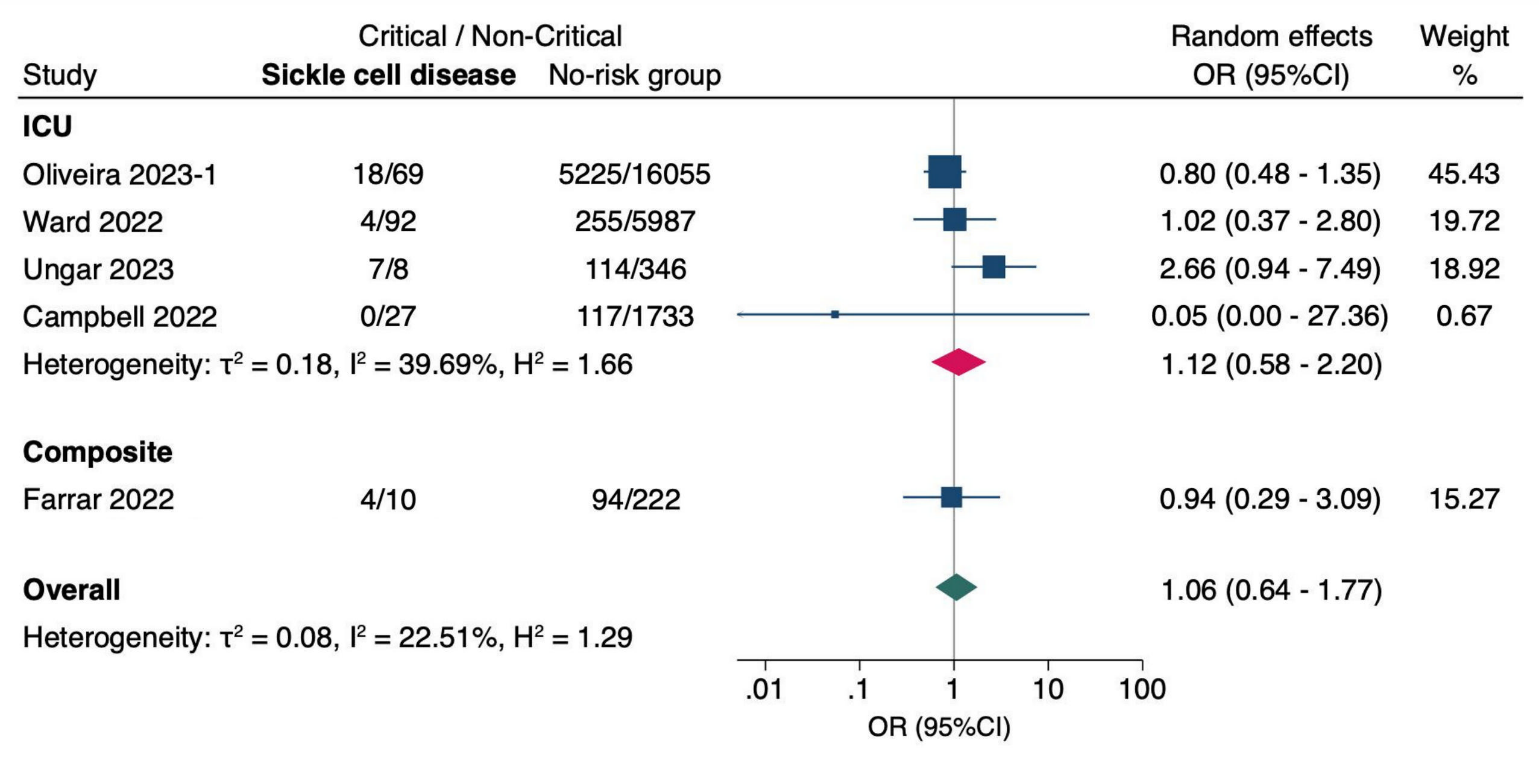

**OR (95% CI)** = odds ratio with 95% confidence interval, estimated using extracted data from published studies.

**ICU** = admission to intensive care unit; **Composite** = combined critical outcomes (ICU, IMV, CVS, or death).
